# A Systematic Review and Network Meta-Analysis for COVID-19 Treatments

**DOI:** 10.1101/2020.12.21.20248621

**Authors:** Chenyang Zhang, Huaqing Jin, Yifeng Wen, Guosheng Yin

## Abstract

**Background:** Numerous interventions for coronavirus disease 2019 (COVID-19) have been investigated by randomized controlled trials (RCTs). This systematic review and Bayesian network meta-analysis (NMA) aim to provide a comprehensive evaluation of efficacy of available treatments for COVID-19.

**Methods:** We searched for candidate COVID-19 studies in WHO COVID-19 Global Research Database, PubMed, PubMed Central, LitCovid, Proquest Central and Ovid up to December 19, 2020. RCTs for suspected or confirmed COVID-19 patients were included, regardless of publication status or demographic characteristics. Bayesian NMA with fixed effects was conducted to estimate the effect sizes using posterior means and 95% equal-tailed credible intervals (CrIs), while that with random effects was carried out as well for sensitivity analysis. Bayesian hierarchical models were used to estimate effect sizes of treatments grouped by their drug classifications.

**Results:** We identified 96 eligible RCTs with a total of 51187 patients. Compared with the standard of care (SOC), this NMA showed that dexamethasone led to lower risk of mortality with an odds ratio (OR) of 0.85 (95% CrI [0.76, 0.95]; moderate certainty) and lower risk of mechanical ventilation (MV) with an OR of 0.68 (95% CrI [0.56, 0.83]; low certainty). For hospital discharge, remdesivir (OR 1.37, 95% CrI [1.15, 1.64]; moderate certainty), dexamethasone (OR 1.20, 95% CrI [1.08, 1.34]; low certainty), interferon beta (OR 2.15, 95% CrI [1.26, 3.74]; moderate certainty), tocilizumab (OR 1.40, 95% CrI [1.05, 1.89]; moderate certainty) and baricitinib plus remdesivir (OR 1.75, 95% CrI [1.28, 2.39]; moderate certainty) could all increase the discharge rate respectively. Recombinant human granulocyte colony-stimulating factor indicated lower risk of MV (OR 0.20, 95% CrI [0.10, 0.40]; moderate certainty); and patients receiving convalescent plasma resulted in better viral clearance (OR 2.28, 95% CrI [1.57, 3.34]; low certainty). About two-thirds of the studies included in this NMA were rated as high risk of bias, and the certainty of evidence was either low or very low for most of the comparisons.

**Conclusion:** The Bayesian NMA identified superiority of several COVID-19 treatments over SOC in terms of mortality, requirement of MV, hospital discharge and viral clearance. These results provide a comprehensive comparison of current COVID-19 treatments and shed new light on further research and discovery of potential COVID-19 treatments.

## Introduction

The pandemic of novel coronavirus disease 2019 (COVID-19), first identified at the end of 2019 in Wuhan, China, has become a global threat to public health. By December 20, 2020, over 76.4 million confirmed cases including 1.69 million deaths have been reported.^1^ Faced with such a global crisis, identifying effective treatments for COVID-19 is of urgent need and paramount importance for clinical researchers. Development of novel drugs typically takes years of concerted efforts and thus most of current research in COVID-19 treatment has been focused on drug repositioning, i.e., investigating the treatment effect of drugs approved for other diseases on COVID-19 patients. Till December 2020, over 7000 clinical trials related to COVID-19 have been registered worldwide. Although numerous medications have been investigated by randomized controlled trials (RCTs), only dexamethasone^2, 3^ and remdesivir^4, 5^ were proven to be clinically effective, while several other antiviral medications were approved for emergency or compassionate use.

During the drug repurposing process, clinicians identify candidate drugs for a specific disease by estimating drug-disease or drug-drug similarities. Drugs with shared chemical structures and mechanisms of action are supposed to deliver similar therapeutic applications.^6^ Not only should research focus on individual treatment for COVID-19, but it is also of great interest to evaluate a class of treatments with shared clinical properties and biochemical structures. For example, systemic corticosteroids including methylprednisolone, dexamethasone and hydrocortisone were reported to be associated with reduced 28-day mortality for COVID-19 patients of critical illness.^7^

With global efforts on pursuing effective treatments during the pandemic, many short-term RCTs of small size were conducted and published at a high rate, and some trials were carried out in a rather rush manner which might cause deterioration of trial quality. Timely summaries and analyses on existing clinical trial results can help to better understand various treatments, early terminate investigation on proven ineffective treatments and provide necessary guidelines for further research and discovery of new treatment. However, with a large number of treatments evaluated against the standard of care (SOC) or other active comparators in RCTs, the conventional pairwise meta-analysis is limited in simultaneous comparisons among multiple trials and fails to capture indirect evidence for treatments that have not been tested in a head-to-head comparison. A network meta-analysis (NMA) which combines both direct and indirect information in a network would be more appropriate to accommodate such complex environments. Several NMA publications provided useful information on the comparative effectiveness of common repurposed drugs for patients with COVID-19.^8, 9^ Our work formulated Bayesian hierarchical models using fixed and random effects respectively, which could effectively borrow information across different trials in the NMA. Not only does our NMA evaluate treatments at the drug level, but it also provides an overall estimated effect at the class level which may contain several drugs of a similar type.

This NMA simultaneously evaluates the clinical benefits of individual treatments and classifications of multiple treatments for COVID-19. We construct a Bayesian hierarchical framework on the effect sizes of treatments and classes, and results were presented and interpreted on both the individual drug level and the class level.

## Methods

This systematic review and NMA were conducted and reported in accordance with the guidelines of Preferred Reporting Items for Systematic Reviews and Meta-Analyses (PRISMA) for NMAs^10^.

### Eligibility criteria

We only included RCTs for suspected or confirmed COVID-19 patients in the systematic review and meta-analysis, because non-randomized trials and observational studies were considered of low certainty of evidence^11^. We included trials published in English and Chinese regardless of the publication status (peer-reviewed or preprint), ways of randomization (double-blind, single-blind or open-label) or study location. Patients in all age groups and of all baseline severities of illness were considered eligible.

We examined treatments for COVID-19 irrespective of dose or duration of administration. Exclusion criteria for the interventions were given as follows: (i) herbal medicine; (ii) preventive interventions (e.g., vaccination and mask wearing); (iii) non-drug supportive care; (iv) exercise, psychological and educational treatments. No restrictions were placed on the control group and we included studies comparing drugs to other active comparator, placebo, SOC or no intervention.

### Outcome measures

The outcomes of interest in the NMA included overall mortality, requirement for mechanical ventilation (MV), discharge from hospital on day 14 or the day closest to that, and viral clearance on day 7 or the day closest to that. We evaluated only binary outcomes since most COVID-19 trials had a less than one-month follow-up,^8^ and for such short-term studies, continuous or survival outcomes might not provide a clinically meaningful and relevant summary for treatment effect^12^. In addition, the clinical definitions of several continuous outcomes, e.g., time to clinical improvement, deterioration and symptom resolution, were not consistent across trials. Different reporting patterns of point and interval estimates for continuous outcomes may also cause additional difficulties and biases in the NMA.

To solve inconsistent reporting for the requirement of MV across trials, we adopted a hierarchical data extraction approach.^8^ We used the number of patients requiring MV during the follow-up period if reported; otherwise, we collected the numbers of patients mechanically ventilated at all available time points and used the maximum value.

### Information sources

We performed exhaustive online search for eligible studies in six databases: WHO COVID-19 Global Research Database^13^, PubMed, PubMed Central, LitCovid, Proquest Central and Ovid. Supplementary Table S1 presents detailed searching strategies for each database.

We updated the literature search weekly to include newly published trials. The current version of manuscript included studies from January 1, 2020 to December 19, 2020. The update of our NMA results will continue on the monthly basis to provide timely assessment on all therapeutic treatments for COVID-19.

### Study selection

Two reviewers independently screened the titles and abstracts using the inclusion criteria. Studies identified as relevant with full text available were further assessed for eligibility. Discrepancies were resolved by the same two reviewers via discussion and, if necessary, a third senior investigator would make the final decision.

### Data collection process

Data extraction was conducted by two investigators independently. For each eligible study, we collected trial characteristics (e.g., the trial registration number, publication status, study status, randomization), interventions, characteristics of participants at baseline (e.g., age, sex, severity of illness, etc.) and outcomes of interest. For the four binary outcomes (mortality, MV, discharge and viral clearance), numbers of events and overall numbers of patients were collected. Two reviewers resolved discrepancies if any via discussion and a third party would adjudicate the conflict. For multiple reports on the same trial, we adopted the peer-reviewed publication if available, or the latest preprint.

### Risk of bias within individual studies

For each eligible trial, we used a revision^8^ of version 2 of the Cochrane risk of bias tool (RoB 2.0)^14^ to assess risk of bias in RCTs. Trials were evaluated on five bias domains including bias arising from the randomization process, bias due to deviations from intended interventions, bias due to missing outcome data, bias in measurement of the outcome and bias in selection of the reported result, and for each domain it was labelled as low, probably low, probably high or high risk of bias. A trial would be rated as low risk if all of five domains were labelled as probably low or low risk of bias; otherwise, it would be rated as high risk. Detailed risk-of-bias judgments were listed in Supplementary Materials. Two reviewers separately completed the RoB assessment and, in presence of any disagreement, a third party would make the final decision.

### Data synthesis and statistical analysis

We conducted a fixed-effects NMA under a Bayesian hierarchical framework. A random-effects NMA was adopted if there existed heterogeneity among trials. However, for most of the comparisons between two treatments, only a few studies were reported and the assessment of heterogeneity might not be reliable. Moreover, sample sizes of trials varied dramatically within some comparisons; for example, the relatively large trials, RECOVERY^2, 15, 16^ and SOLIDARITY^17^, would typically dominate the estimates. In the network, each node represents a treatment, regardless of dose or duration of administration. For the studies involving different doses or durations of the same drug^18^, we aggregated data of the same drug into one arm. Each multi-arm trial was treated as a single study in the network analysis, instead of splitting it into several two-arm sub-trials. Interventions composed of more than one drug were treated as separate nodes with each corresponding to one drug. In addition, interval estimates could be imprecise for treatments tested with small sample size^8, 19^. Therefore, for each clinical outcome, we excluded the treatments appearing in only one trial with fewer than 100 patients to alleviate potential risk caused by inadequate information. We plotted the network for each outcome of interest using the igraph^20^ package of R version 4.0.3 (RStudio, Boston, MA) as shown in Supplementary Materials.

The Bayesian NMA model was built upon assuming binomial distribution of observed data with a logit link on binomial probability and the relative effects were evaluated at the log odds scale while assuming consistency^21^. Based on the Anatomical Therapeutic Chemical Classification System with Defined Daily Doses (ATC/DDD)^22^ published by WHO, we classified the included drugs by the second level of their ATC/DDD codes. For example, both hydroxychloroquine (P01BA02) and chloroquine (P01BA01) belong to antiprotozoals (P01). For the investigational drugs without ATC/DDD codes, we determined their classifications according to the pharmacological mechanism and therapeutic use. Detailed information on the classifications of eligible drugs is presented in Supplementary Table S2. We considered a hierarchical structure for investigated interventions where the relative effects compared with the SOC were nested within drug classifications. The Bayesian multi-layer structure enables us to model the variability on different levels and estimate the treatment benefits of individual drugs and their classifications simultaneously. We assigned the same vague inverse-gamma prior distributions to the variance parameters and normal prior distributions to the effect size parameters. The Bayesian hierarchical structure for the NMA is shown in Supplementary Figure S1.

We fitted the Bayesian NMA model and generated posterior samples of parameters using the Markov chain Monte Carlo (MCMC) algorithm. The treatment effects of eligible drugs were evaluated in terms of odds ratios (ORs) which were estimated by the posterior means and corresponding equal-tailed 95% credible intervals (CrI). To obtain the direct and indirect estimates for treatment effects and assess local inconsistency in the network, we considered the node-splitting method^23^. The MCMC sampling was performed using the jagsUI^24, 25^ package of R, and further network analyses were performed using the gemtc^26^ package of R.

### Certainty of the evidence

The grading of recommendations assessment, development and evaluation (GRADE) approach for NMA^11^ was used to rate the certainty of evidence of NMA estimates. Two investigators rated the certainty of each treatment comparison independently and resolved discrepancies by discussions and, if necessary, consulted with a third party. We considered each of the following seven criteria: RoB^27^, inconsistency^28^, indirectness^29^, publication bias^30^, intransitivity^11^, incoherence^31^ and imprecision^32^, to evaluate the certainty of NMA estimates as high, moderate, low and very low. The certainty would be rated down by one scale if no less than half of the trials directly comparing two treatments were of high RoB. The inconsistency was assessed by the Higgins I^2^ statistics and the Cochran Q test^33^ and we identified inconsistency if I^2^>50% or p-value<0.05 from the Cochran Q test. We rated down by one scale for publication bias due to asymmetric funnel plots or industry sponsorship.^30^ Baseline patient characteristics, especially the severity of illness, were used for the evaluation of intransitivity. The minimally contextualised approach^34^ considering only the CrI and null effect was used for assessing imprecision. We set a threshold of 0.02 rather than the null value 0 in the log OR scale in order to demonstrate more convincing evidence in each paired comparison. Detailed ratings and rationales for downgrading were provided in the Supplementary Materials.

### Subgroup and sensitivity analysis

Planned subgroup analyses were conducted for peer-reviewed studies only and mild/moderate versus severe/critical COVID-19 patients at baseline. In addition to Bayesian fixed-effects NMA, we also performed Bayesian random-effects NMA and presented results in Supplementary Materials. In the primary analysis, we treated three published studies for the RECOVERY trial^2, 15, 16^ which respectively compared lopinavir/ritonavir, dexamethasone and hydroxychloroquine versus SOC, as a four-arm trial according to the RECOVERY protocol v6.0.^35^ The SOC group with the largest number of participants^2^ was used in the primary analysis. Similarly, although the SOLIDARTIY trial^17^ reported four pairwise comparisons between interventions and SOC, we treated it as a five-arm trial due to substantial overlap across SOC groups. Furthermore, we performed a sensitivity analysis by treating RECOVERY as three two-arm trials and SOLIDARITY as four two-arm trials.

## Results

According to the prespecified inclusion and exclusion criteria, we identified and screened 4563 titles and abstracts after de-duplication. Out of these, 146 studies were further reviewed for full text and 96 eligible studies reporting 93 unique randomized clinical trials were included in the systematic review,^2, 5, 15-18, 36-125^ where the RECOVERY trial was reported in four studies.^2, 15, 16, 76^ Figure 1 summarizes the process of our study selection.

**Figure 1.**
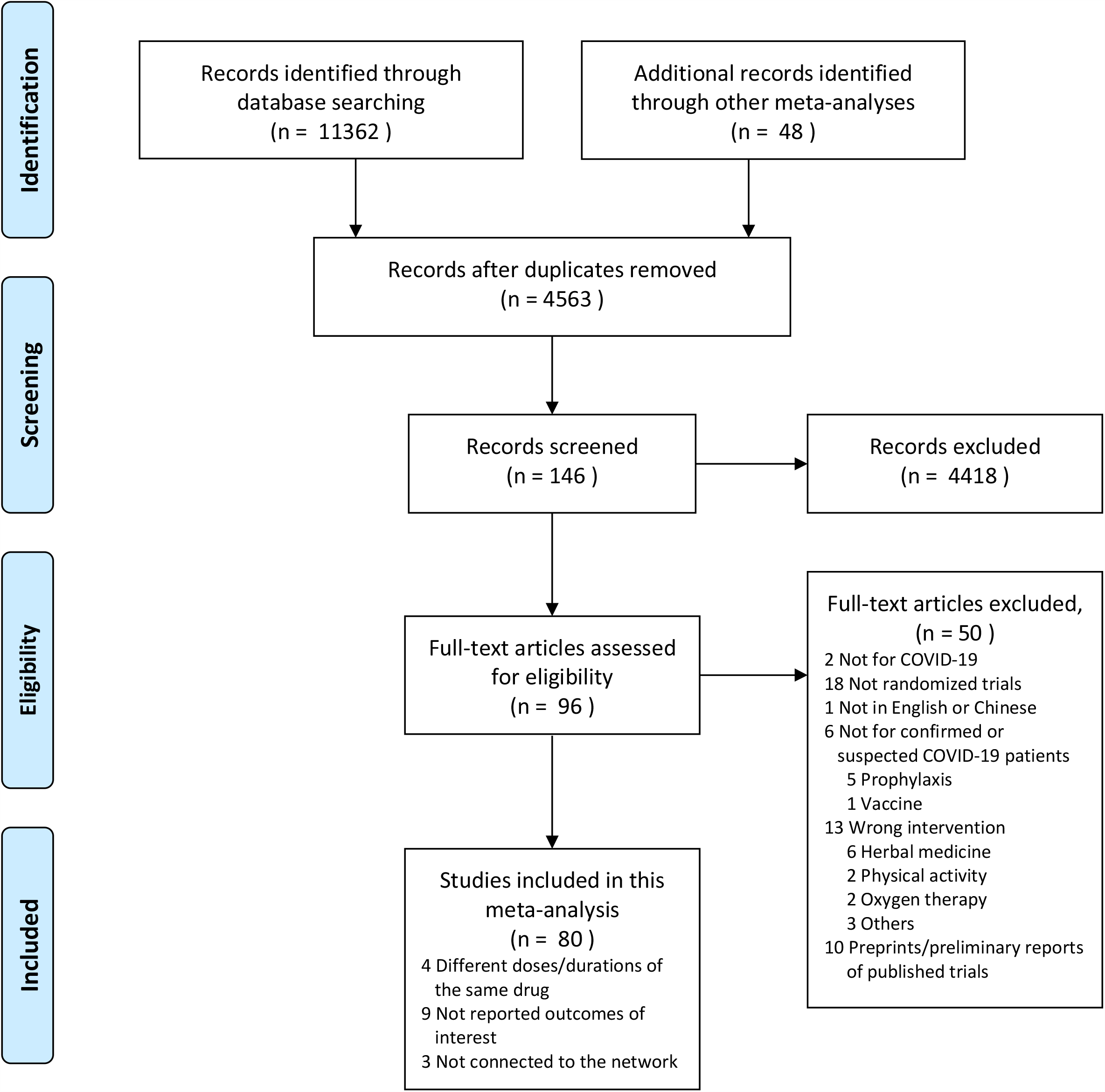
Flow diagram of the study selection

Out of these 96 studies included in the NMA, 68 were peer-reviewed and the other 28 were preprints. Over two-thirds of the studies (65/96) were open-label, 27 were double-blind and the remaining 4 were single-blind in randomization. Most of the studies reported completed clinical trials (84/96) rather than ongoing (6/96) or early terminated trials (6/96) and investigated hospitalized COVID-19 patients (88/96). Twenty-four studies were conducted in China, ten in Brazil, nine in Iran, eight in the USA and nine in multi-sites across countries. Among the 96 studies, ten were multi-arm and the rest were two-arm; 69 studies compared the investigated intervention with SOC, 19 with other active comparators and the other eight with both SOC and other interventions. About half of the studies (47/96) were of small sample size which enrolled less than 100 patients in the intention-to-treat population, seven studies recruited over 1000 patients, including four reports for the RECOVERY trial^2, 15, 16, 76^, one for the SOLIDARITY^17^ trial, one for remdesivir^5^ and one for baricitinib plus remdesivir^83^. Of 74 studies which recorded the baseline severity of illness, 28 involved patients mainly with severe or critical COVID-19 and 46 were mainly for mild/moderate disease. Detailed trial and patient characteristics are given in Supplementary Materials.

Sixteen randomized trials were not considered in the meta-analysis. Among them, four studies investigated different doses or durations of administration of the same intervention without comparisons with other interventions or SOC.^45, 46, 64, 71^ Eight trials did not specify the outcomes of interest.^55, 58, 68, 81, 93, 101, 122, 123^ Treatments in four trials were not connected in the network.^73, 77, 86, 98^

### Clinical outcomes Mortality

A total of 74 studies^2, 5, 15-18, 36-39, 41-44, 47-54, 56, 57, 59-63, 65-67, 69, 70, 72, 74-76, 78, 79, 82, 83, 85, 87, 88, 90-92, 94-97, 99, 100, 103-121, 125^ including 48622 patients reported all-cause mortality. After filtering out treatments with small sample size following the specified criteria, the network included azithromycin, hydroxychloroquine, hydroxychloroquine plus azithromycin, colchicine, arbidol (umifenovir), favipiravir, remdesivir, lopinavir/ritonavir, convalescent plasma, methylprednisolone, dexamethasone, hydrocortisone, immunoglobulin, interferon beta, recombinant human granulocyte colony-stimulating factor (GCSF), tocilizumab, vitamin D_3_, baricitinib plus remdesivir, sulodexide and SOC. Among the 50 trials which investigated the included treatments, the risk of bias was accessed as low for 18 trials. The other trials were judged at high risk of bias mainly because of bias due to deviations from the intended intervention (see Supplementary Materials). Compared with SOC, the Bayesian NMA with fixed-effects showed that only dexamethasone (OR 0.85, 95% CrI [0.76, 0.95]; moderate certainty) and lopinavir/ritonavir (OR 0.88, 95% CrI [0.79, 0.97]; moderate certainty) could reduce the mortality rate with statistical significance. Among all interventions, only hydroxychloroquine, tocilizumab and vitamin D3 reported higher risk of death versus control but not statistically significant, as shown in Figure 2. Colchicine (very low certainty), lopinavir/ritonavir (low certainty), hydrocortisone (moderate certainty), immunoglobulin (low certainty), baricitinib plus remdesivir (very low certainty) and sulodexide (very low certainty) might be of potential benefit compared to SOC with posterior probability favouring treatment larger than 0.9. The class of immunosuppressants plus antivirals for systemic use and antithrombotic agents might be of potential benefit due to their relatively large posterior probabilities favouring treatment (larger than 0.9) and the other classes showed no difference from SOC.

**Figure 2.**
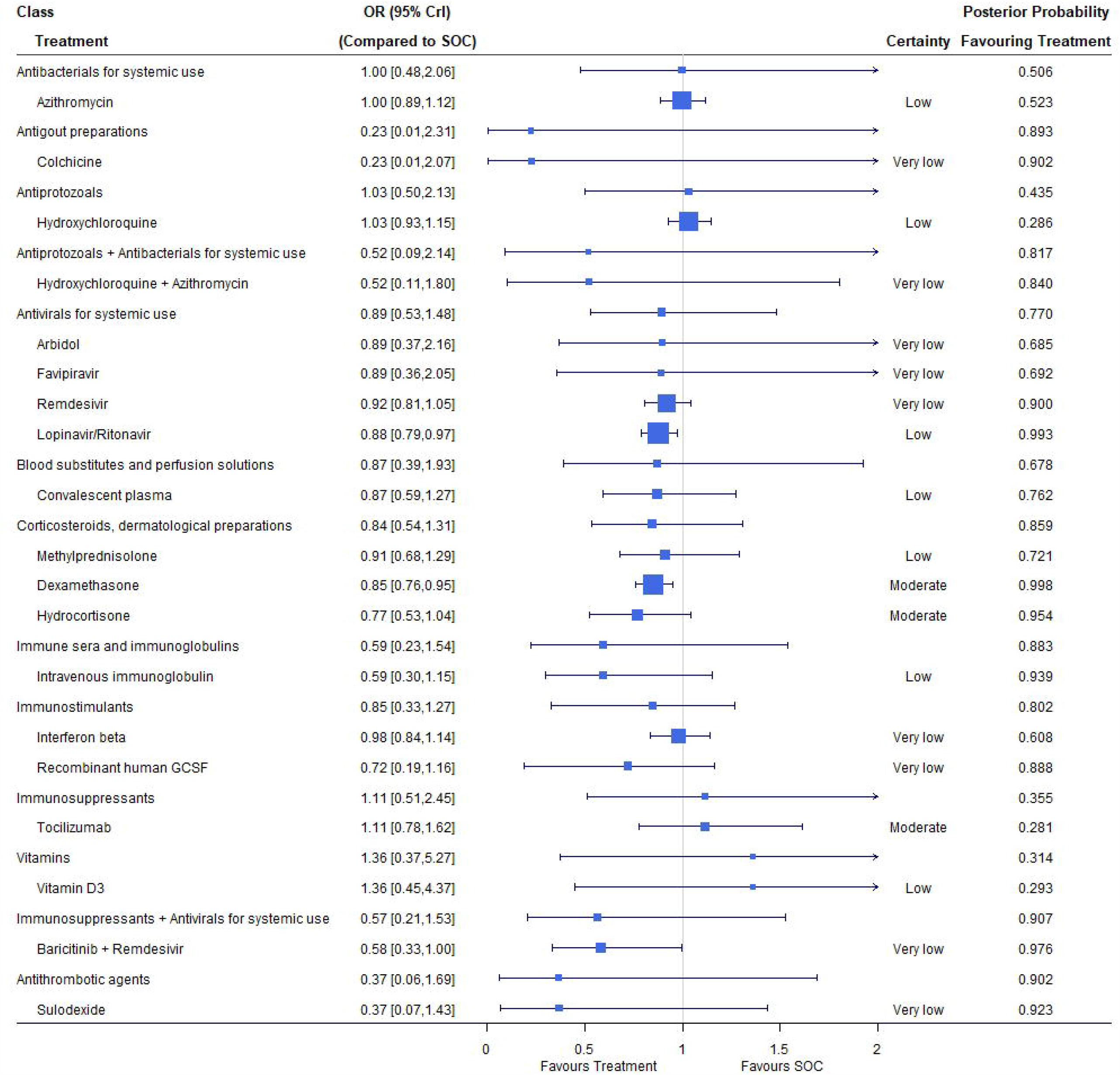
Mortality under treatments compared with the standard of care (SOC); OR is the odds ratio and CrI represents credible interval.

For the random-effects model, the estimated relative effects versus SOC were similar to those in the fixed-effects model but with wider credible intervals, e.g., dexamethasone with an OR of 0.82 (95% CrI [0.63, 1.04]). In the sensitivity analysis by treating three studies of RECOVERY^2, 15, 16^ as three two-arm trials and SOLIDARITY^17^ as four two-arm trials, all estimates were close to those in the primary analysis except for lopinavir/ritonavir (see Supplementary Materials), which reported an OR of 0.88 (95% CrI [0.79, 0.97]) in the primary analysis and 1.01 (95% CrI [0.90, 1.14]) in the sensitivity analysis. The gap on the 28-day mortality rate between the SOC arm with the largest number of patients^2^ (25.7%, 1110/4321) and the SOC arm of the lopinavir/ritonavir trial^15^ (22.4%, 767/3424) mainly contributed to the discrepancy in the estimates of OR for lopinavir/ritonavir versus SOC.

### Mechanical ventilation

Overall, 48 studies reported the number of patients required for MV during the study period.^2, 5, 15-17, 36-39, 41-44, 47-50, 56, 57, 60-63, 65, 69, 72, 76, 78, 82, 83, 87, 92, 95-97, 100, 103-105, 108-110, 112-114, 116, 117, 120^ with 41405 patients and 4455 events. We included azithromycin, hydroxychloroquine, hydroxychloroquine plus azithromycin, remdesivir, lopinavir/ritonavir, convalescent plasma, methylprednisolone, dexamethasone, hydrocortisone, immunoglobulin, interferon beta, recombinant human GCSF, tocilizumab, vitamin D_3_, baricitinib plus remdesivir, sulodexide and SOC as treatment nodes in the NMA. We evaluated the risk of bias for 34 RCTs and among them 12 were rated as low risk. Compared with SOC, dexamethasone (OR 0.68, 95% CrI [0.56, 0.83]; low certainty) and recombinant human GCSF (OR 0.20, 95% CrI [0.10, 0.40]; moderate certainty) had significantly lower MV rates (Figure 3). Tocilizumab (moderate certainty), vitamin D_3_ (moderate certainty) and baricitinib plus remdesivir (very low certainty) showed potential benefit over SOC in reducing MV events with posterior probability favouring treatment higher than 0.9. Patients randomized to azithromycin, hydroxychloroquine, hydroxychloroquine plus azithromycin and lopinavir/ritonavir had higher risk of MV than those receiving SOC and all were rated as low or very low certainty. The class of immunostimulants containing interferon beta and recombinant human GCSF was of potential benefit compared with SOC due to its relatively large posterior probability favouring treatment (0.94) and the other classes showed no difference from SOC. The Bayesian random-effects NMA reported similar point estimates with substantial wider interval estimates. Dexamethasone had an OR of 0.70 with 95% CrI [0.47, 1.08] covering the null effect, while recombinant human GSCF still yielded a significantly lower rate of MV compared with SOC (OR 0.27, 95% CrI [0.11, 0.67]) under the random-effects model. Whether RECOVERY and SOLIDARITY were treated as multi-arm trials or multiple two-arm trials had no influence on estimates of relative effects since the MV rates in the SOC groups of the three RECOVERY studies^2, 15, 16^ were similar and so was the SOLIDARTITY trial.^17^

**Figure 3.**
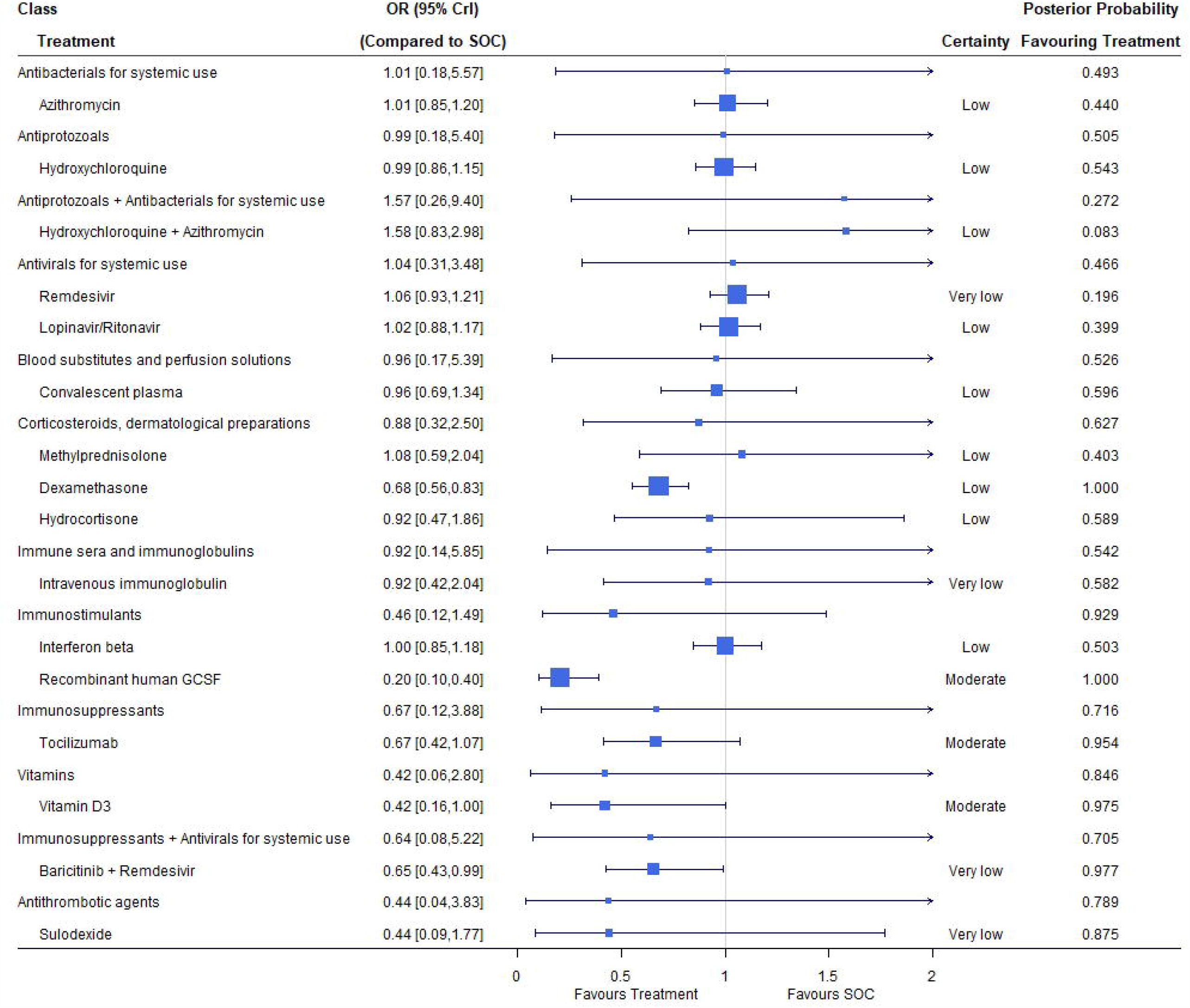
Requirement of mechanical ventilation under treatments compared with the standard of care (SOC); OR is the odds ratio and CrI represents credible interval.

### Discharge (closest to 14 days)

The hospital discharge rate was reported in 36 studies including 31436 patients and 19365 events.^2, 5, 15, 16, 36, 43, 47, 48, 50, 54, 60, 65, 66, 69, 75, 76, 79, 80, 83, 85, 88, 89, 91, 92, 96, 100, 103, 107-110, 112, 113, 116, 117, 120^ Treatment nodes included in the network were azithromycin, hydroxychloroquine, hydroxychloroquine plus azithromycin, favipiravir, remdesivir, lopinavir/ritonavir, convalescent plasma, dexamethasone, interferon beta, tocilizumab, baricitinib plus remdesivir and SOC. Out of the 26 RCTs included in the NMA, nine were assessed as low risk of bias. Patients who received remdesivir (OR 1.37, 95% CrI [1.15, 1.64]; moderate certainty), lopinavir/ritonavir (OR 1.30, 95% CrI [1.16, 1.47]; low certainty), dexamethasone (OR 1.20, 95% CrI [1.08, 1.34]; low certainty), interferon beta (OR 2.16, 95% CrI [1.26, 3.74]; moderate certainty), tocilizumab (OR 1.40, 95% CrI [1.05, 1.89]; moderate certainty) and baricitinib plus remdesivir (OR 1.75, 95% CrI [1.28, 2.39]; moderate certainty) had a higher discharge rate compared with those in the SOC arm. Hydroxychloroquine (OR 0.87, 95% CrI [0.78, 0.96]; moderate certainty) was even inferior to SOC and led to high risk of hospitalization at around 14 days (Figure 4). The class of immunostimulants containing only interferon beta showed potential benefit on hospital discharge with posterior probability favouring treatment equal to 0.95.

**Figure 4.**
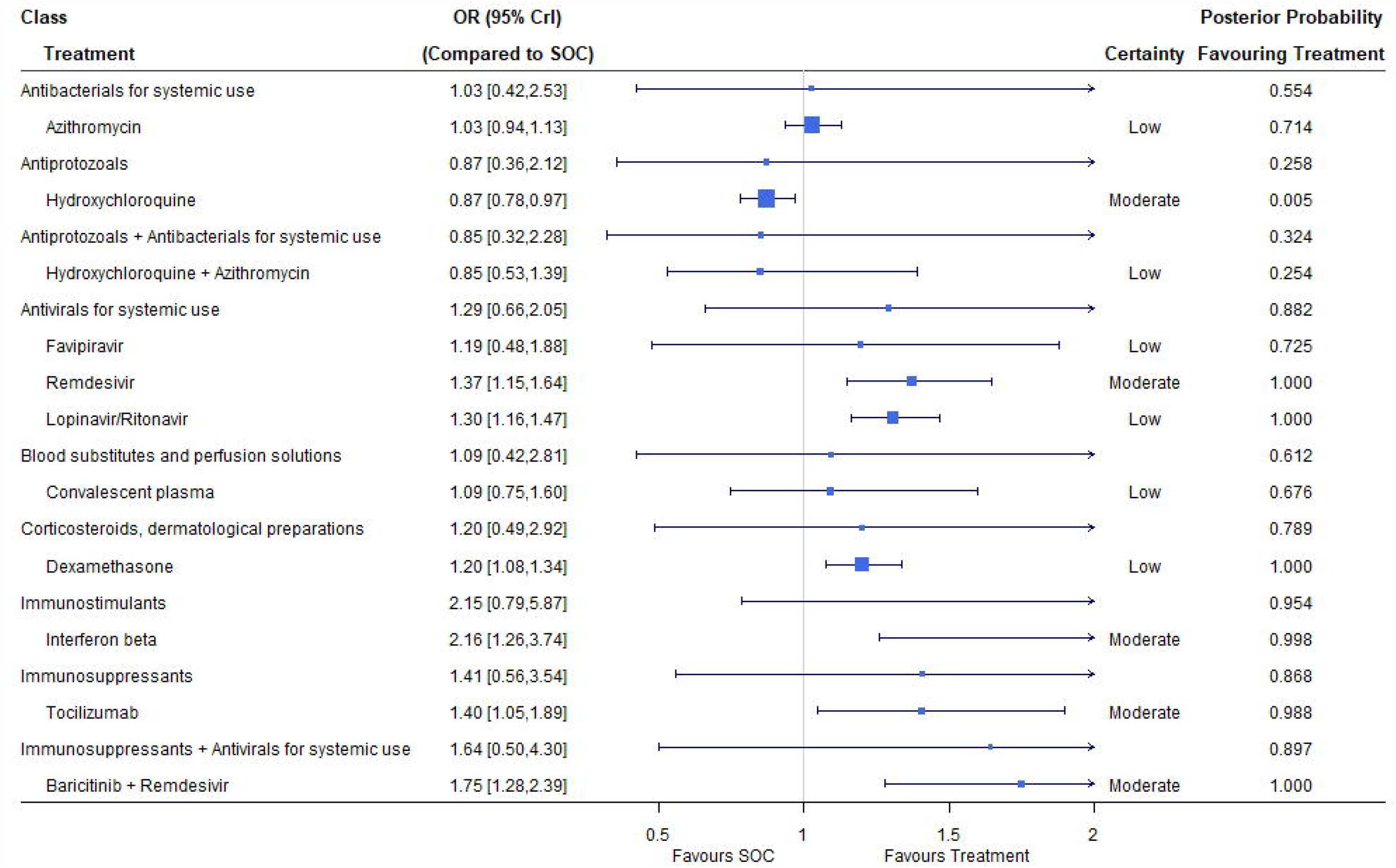
Discharge (closest to 14 days) under treatments compared with the standard of care (SOC); OR is the odds ratio and CrI represents credible interval.

For the random-effects model, with wider interval estimates, remdesivir (OR 1.36, 95% CrI [1.06, 1.75]), lopinavir/ritonavir (OR 1.38, 95% CrI [1.09, 1.82]) and interferon beta (OR 2.17, 95% CrI [1.23, 3.88]) still maintained their significant benefit over SOC in terms of discharge. However, a sensitivity analysis treating RECOVERY^2, 15, 16^ as three two-arm trials reported an insignificant OR of 1.01 (95% CrI [0.89, 1.14]) for lopinavir/ritonavir versus SOC. Similar to the case when evaluating mortality, such discrepancy was caused by the different event rates in the two SOC arms used.^2, 15^

### Viral clearance (closest to 7 days)

Twenty-one studies including 3183 patients and 1403 events reported viral clearance rates. ^39, 43, 52, 53, 65, 67, 79, 80, 82, 84, 88, 90, 92, 99, 102, 115, 117, 119-121, 124^ Treatment nodes in the network included hydroxychloroquine, nitazoxanide, hydroxychloroquine plus azithromycin, favipiravir, remdesivir, convalescent plasma, methylprednisolone and SOC. Out of these 15 trials included in the NMA, six were judged at low risk of bias. Under the fixed-effects NMA, the OR of nitazoxanide versus SOC was 1.72 (95% CrI [1.13, 2.80]; low certainty) and that of convalescent plasma was 2.28 (95% CrI [1.57, 3.34]; low certainty), which indicated that administration of nitazoxanide and convalescent plasma improved virologic cure (Figure 5). Hydroxychloroquine and favipiravir might be of potential benefit compared with SOC due to posterior probability favouring treatment larger than 0.9, although both had very low certainty. The class of blood substitutes and perfusion solutions contained only one treatment convalescent plasma, which probably improved viral clearance with an OR of 2.28 (95% CrI [0.48, >10]; posterior probability favouring treatment 0.92).

**Figure 5.**
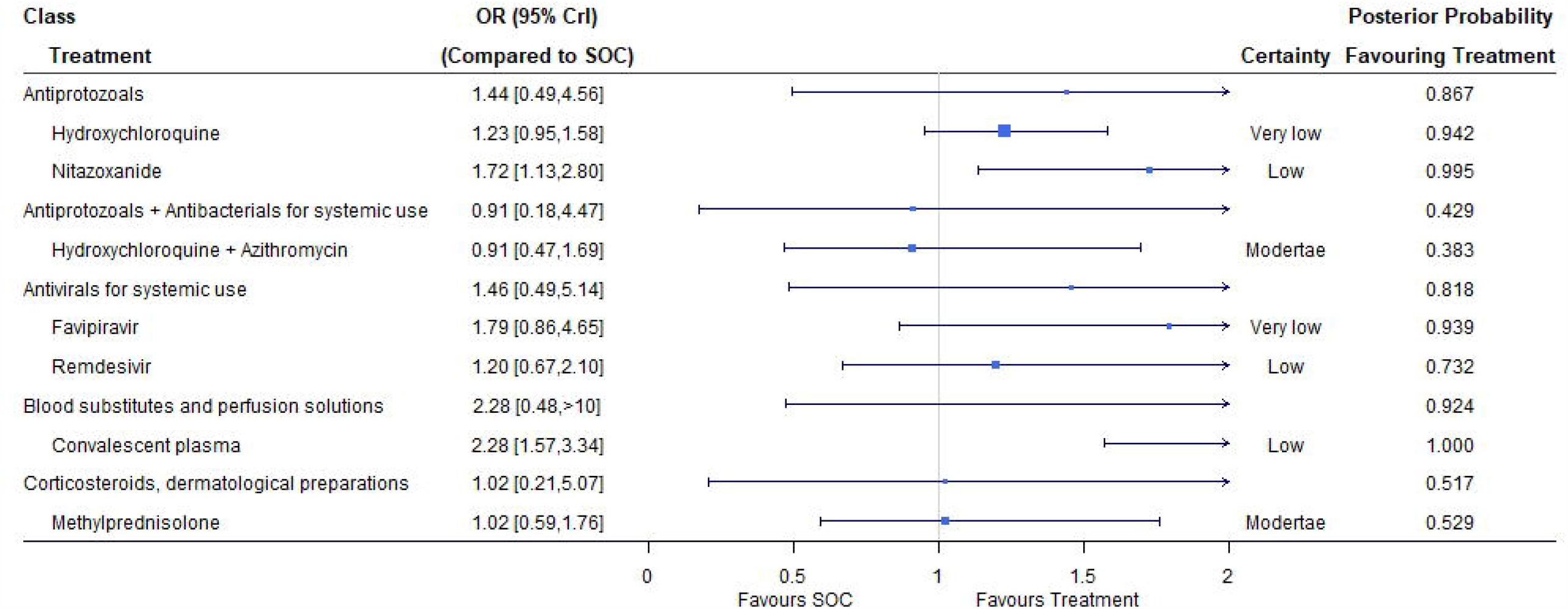
Viral clearance (closest to 7 days) of treatments compared with the standard of care (SOC); OR is the odds ratio and CrI represents credible interval.

For the random-effects model, nitazoxanide (OR 1.34, 95% CrI [0.53, 3.97]) did not show superiority over SOC, while convalescent plasma (OR 2.87, 95% CrI [1.23, 7.31]) was still effective in virus elimination. The RECOVERY trial did not report viral clearance and thus no sensitivity analysis was carried out.

## Discussion

In this systematic review and NMA, we provided a detailed summary of trial characteristics of published RCTs for COVID-19 patients up to December 7, 2020 and reported effectiveness of treatments at both the drug and class levels compared with SOC in terms of mortality, MV, discharge and viral clearance. Dexamethasone was shown to reduce the risk of mortality (moderate certainty), MV (low certainty) and hospitalization (low certainty). Recombinant GCSF resulted in fewer events of MV with moderate certainty of evidence. Patients who received nitazoxanide and convalescent plasma (both low certainty) had a higher viral elimination rate in comparison with SOC. Remdesivir, lopinavir/ritonavir, interferon beta, tocilizumab and baricitinib plus remdesivir demonstrated their effectiveness on clinical improvement with significantly higher 14-day discharge rates compared with SOC. However, most of the comparisons were rated as low or very low certainty of evidence.

At the class level of treatments, immunosuppressants plus antivirals for systemic use and antithrombotic agents might reduce mortality, immunostimulants showed potential clinical benefit over SOC in reducing MV and hospitalization, and blood substitutes and perfusion solutions probably increased the rate of viral clearance on day 7, with posterior probability favouring treatment larger than 0.9. For other classes and outcomes, we observed no difference from SOC.

### Strength and limitations

Not only was this NMA timely conducted, but it also included a wide range of RCTs, which contained not only common drugs but also interferons, blood products, mineral and vitamin supplementations without restrictions on publication status. Risk of bias for individual studies and rating of certainty for NMA estimates were carefully conducted and detailed information and reasons for corresponding assessments are given in Supplementary Materials. Treatment effects for multiple treatments were evaluated in a network at both the individual drug and class levels.

This study has several limitations. The primary one is the low or very low certainty of evidence for most NMA estimates. For each outcome of interest, about two-thirds of trials were graded as high risk and the major reason was lack of blinding in the trials, leading to potential bias in the NMA. At the early stage of COVID-19 pandemic, with limited clinical resources and urgent need to obtain trial results, many RCTs were conducted with simplified procedures, e.g., no placebo prepared, including the large RECOVERY and SOLIDARITY trials. As time goes on, such situation has changed and recently many double-blind RCTs have been conducted and published. Moreover, networks of treatments were sparse because most of the included studies evaluated interventions versus SOC and there were few direct comparisons between interventions. As we considered COVID-19 RCTs regardless of baseline patient characteristics, intransitivity existed in many indirect comparisons. For example, hydroxychloroquine trials usually investigated patients with mild or moderate COVID-19, while patients treated by convalescent plasma were mainly of severe or critical illness. Detailed subgroup analysis might help to solve such problem.

Another limitation of this study arises from the evaluation of NMA estimates at the class level. Many investigated classes in the NMA contained only one treatment, leading to large variation and thus insignificant interval estimates. To confirm the superiority of a class of drugs, one should present evidence of more strengths. More treatments can be included in the NMA if the exclusion criteria of treatment nodes were relaxed, while such operation would introduce additional bias due to trials with small sample size.

With a large number of completed COVID-19 trials, studies reporting positive results or with large sample size were more likely to be published, leading to possible publication bias.^30^ We considered studies irrespective of publication status to alleviate publication bias and obtained as much information as possible. While extra attention should be paid on the evidence implied by only preprints. For example, the significant benefit of nitazoxanide on viral clearance was shown in only one preprint,^102^ and clinical results without peer-reviews should not be trusted equally as those published.

Different approaches to dealing with the RECOVERY trial led to discrepancies between the results of the primary and sensitivity analyses, especially for lopinavir/ritonavir. The RECOVERY trial was designed as a multi-arm trial^35^ while the numbers of patients randomized to SOC and event rates of outcomes of interest were different across different reports.^2, 15, 16^ Although lopinavir/ritonavir showed potential clinical effects on the reduction of mortality and increase of the discharge rate, credibility of this finding warrants extra caution.

## Conclusion

This systematic review and meta-analysis showed that dexamethasone could reduce the mortality, requirements of MV and increase the discharge rate. Administration of remdesivir led to a higher discharge rate, but showed no difference from SOC for mortality, MV and viral clearance. Patients receiving recombinant GCSF had lower risk of MV. Nitazoxanide and convalescent plasma could improve the viral elimination, although the evidence of nitazoxanide was supported solely by one preprint.

Lopinavir/ritonavir showed clinical benefits on the increase of the discharge rate and reduction of mortality, while inconsistency between the primary and sensitivity analyses caused extra uncertainty. At the treatment class level, immunosuppressants plus antivirals for systemic use and antithrombotic agents could probably reduce the risk of death, immunostimulants tended to reduce MV and increase discharge, and blood substitutes and perfusion solutions might improve viral clearance.

## Data Availability

The data that support the findings of this study are available from the corresponding author, G Yin, upon reasonable request.

