## Supplementary Materials for "A Systematic Review and Network Meta-Analysis for COVID-19 Treatments"

### Supplementary Methods

#### Model structure of network meta-analysis

For the $i$-th trial, suppose the sample size is $n_{i,x}$ and the number of events is $y_{i,x}$ for treatment $x$. We assume a binomial model,

$$y_{i,x} \sim \text{Binomial}(p_{i,x}, n_{i,x}),$$

where $p_{i,x}$ is the probability of the event of interest.

Taking arm $b_i$ as the baseline treatment for trial $i$, under the logit link function, our fixed-effects model is formulated as

$$\text{logit}(p_{i,x}) = \mu_i + d_{b_i,x},$$

$$d_{b_i,x} = d_{0,x} - d_{0,b_i},$$

where $\mu_i$ is the effect size of baseline treatment $b_i$, $d_{b_i,x}$ is the relative effect of treatment $x$ compared with $b_i$ in the $i$-th trial, $d_{0,x}$ is the relative treatment effect of $x$ compared with the standard of care (SOC) labelled by 0. Under a Bayesian hierarchical model, we assume the relative treatment effect as

$$d_{0,x} \sim N(\theta_{c_x}, \sigma^2),$$

$$d_{0,b_i} = 0 \text{ if } b_i = 0,$$

$$d_{0,b_i} \sim N\left(\theta_{c_{b_i}}, \sigma^2\right) \text{ if } b_i \neq 0,$$

where $c_x$ is the class that treatment $x$ belongs to, and $\theta_{c_x}$ is the relative treatment effect of class $c_x$ compared with SOC.

For the random-effects model, we specify a Bayesian hierarchical structure as follows,

$$\text{logit}(p_{i,x}) = \mu_i + \delta_{i,b_i,x},$$

$$\delta_{i,b_i,x} \sim N(d_{b_i,x}, \tau^2),$$

$$d_{b_i,x} = d_{0,x} - d_{0,b_i},$$

$$d_{0,x} \sim N(\theta_{c_x}, \sigma^2),$$

$$d_{0,b_i} = 0 \text{ if } b_i = 0,$$

$$d_{0,b_i} \sim N\left(\theta_{c_{b_i}}, \sigma^2\right) \text{ if } b_i \neq 0,$$

Our prior distributions are given as follows:

- $\mu_i \sim N(0, 10^2), i = 1, \dots, n$, where $n$ is the number of trials.
- $\theta_c \sim N(0, 10^2), c = 1, \dots, m$, where $m$ is the number of classes.
- $\sigma^2 \sim \text{InverseGamma}(0.01, 0.01)$.
- $\tau^2 \sim \text{InverseGamma}(0.01, 0.01)$

#### Supplementary Tables

##### Table S1. Detailed search strategy

| Database | Search strategy |
|---|---|
| WHO COVID-19 Global Research Database | (tw:(randomized)) OR (tw:(randomised)) |
| PubMed | ((randomized[Title/Abstract]) OR (randomised[Title/Abstract])) AND (coronavirus OR "corona virus" OR coronavirinae OR coronaviridae OR betacoronavirus OR covid19 OR "covid 19" OR nCoV OR "CoV 2" OR CoV2 OR sarscov2 OR 2019nCoV OR "novel CoV") |
| PubMed Central | "severe acute respiratory syndrome coronavirus 2"[Supplementary Concept] OR "severe acute respiratory syndrome coronavirus 2"[All Fields] OR "ncov"[All Fields] OR "2019-nCoV"[All Fields] OR "COVID-19"[All Fields] OR "SARS-CoV-2"[All Fields] OR ((("coronavirus"[MeSH Terms] OR "coronavirus"[All Fields]) OR "cov"[All Fields]) AND 2020/01[PubDate] : 3000[PubDate]) AND ("randomized"[Abstract] OR "randomised"[Abstract]) |
| LitCovid | 'randomized' OR 'randomised' |
| PROQUEST | ab((randomized) OR (randomised)) AND (coronavirus OR "corona virus" OR coronavirinae OR coronaviridae OR betacoronavirus OR covid19 OR "covid 19" OR nCoV OR "CoV 2" OR CoV2 OR sarscov2 OR 2019nCoV OR "novel CoV") |
| Ovid | (coronavir* or corona virus* or betacoronavir* or covid19 or covid 19 or nCoV or novel CoV or CoV 2 or CoV2 or sarscov2 or 2019nCoV or wuhan virus*).mp. and (randomized or randomised).ab. |

#### Table S2. Treatments included and their classifications based on Anatomical Therapeutic Chemical Classification System with Defined Daily Doses.

| Drug Type (Second level) | Drug Name | ATC/DDD Code | First level |
|---|---|---|---|
| ANTHELMINTICS + ANTIBACTERIALS FOR SYSTEMIC USE | Ivermectin + Doxycycline | P02CF01 + J01AA02 | ANTIPARASITIC PRODUCTS, INSECTICIDES AND REPELLENTS + ANTIINFECTIVES FOR SYSTEMIC USE |
| ANTHELMINTICS | Ivermectin | P02CF01 | ANTIPARASITIC PRODUCTS, INSECTICIDES AND REPELLENTS |
| ANTIBACTERIALS FOR SYSTEMIC USE | Azithromycin | J01FA10 | ANTIINFECTIVES FOR SYSTEMIC USE |
| ANTIEMETICS AND ANTINAUSEANTS | Aprepitant | A04AD12 | ALIMENTARY TRACT AND METABOLISM |
| ANTIGOUT PREPARATIONS | Colchicine | M04AC01 | MUSCULO-SKELETAL SYSTEM |
| ANTIGOUT PREPARATIONS | Febuxostat | M04AA03 | MUSCULO-SKELETAL SYSTEM |
| ANTINEOPLASTIC AGENTS | Ruxolitinib | L01XE18 | ANTINEOPLASTIC AND IMMUNOMODULATING AGENTS |
| ANTIPROTOZOALS | Hydroxychloroquine | P01BA02 | ANTIPARASITIC PRODUCTS, INSECTICIDES AND REPELLENTS |
| ANTIPROTOZOALS | Chloroquine | P01BA01 | ANTIPARASITIC PRODUCTS, INSECTICIDES AND REPELLENTS |
| ANTIPROTOZOALS | Nitazoxanide | P01AX11 | ANTIPARASITIC PRODUCTS, INSECTICIDES AND REPELLENTS |
| ANTIPROTOZOALS + ANTIBACTERIALS FOR SYSTEMIC USE | Hydroxychloroquine + Azithromycin | P01BA02 + J01FA10 | ANTIPARASITIC PRODUCTS, INSECTICIDES AND REPELLENTS + ANTIINFECTIVES FOR SYSTEMIC USE |
| ANTIPROTOZOALS + ANTIVIRALS FOR SYSTEMIC USE | Hydroxychloroquine + Darunavir | P01BA02 + J05AE10 | ANTIPARASITIC PRODUCTS, INSECTICIDES AND REPELLENTS + ANTIINFECTIVES FOR SYSTEMIC USE |
| ANTIPROTOZOALS + MINERAL SUPPLEMENTS | Hydroxychloroquine + Zinc | P01BA02 + A12CB01 | ANTIPARASITIC PRODUCTS, INSECTICIDES AND REPELLENTS + ALIMENTARY TRACT AND METABOLISM |
| ANTIVIRALS FOR SYSTEMIC USE | Arbidol | J05AX13 | ANTIINFECTIVES FOR SYSTEMIC USE |
| ANTIVIRALS FOR SYSTEMIC USE | Triazavirin | NA | ANTIINFECTIVES FOR SYSTEMIC USE |
| ANTIVIRALS FOR SYSTEMIC USE | Favipiravir | J05AX27 | ANTIINFECTIVES FOR SYSTEMIC USE |
| ANTIVIRALS FOR SYSTEMIC USE | Baloxavir marboxil | J05AX25 | ANTIINFECTIVES FOR SYSTEMIC USE |
| ANTIVIRALS FOR SYSTEMIC USE | Sofosbuvir + Daclatasvir + Ribavirin | J05AP08 + J05AP07 + J05AP01 | ANTIINFECTIVES FOR SYSTEMIC USE |
| ANTIVIRALS FOR SYSTEMIC USE | Remdesivir | NA | ANTIINFECTIVES FOR SYSTEMIC USE |
| ANTIVIRALS FOR SYSTEMIC USE | Sofosbuvir + Daclatasvir | J05AP08 + J05AP07 | ANTIINFECTIVES FOR SYSTEMIC USE |
| ANTIVIRALS FOR SYSTEMIC USE | Azvudine | NA | ANTIINFECTIVES FOR SYSTEMIC USE |
| ANTIVIRALS FOR SYSTEMIC USE | Lopinavir/ritonavir | J05AR10 | ANTIINFECTIVES FOR SYSTEMIC USE |
| ANTIVIRALS FOR SYSTEMIC USE | Favipiravir + Interferon beta | J05AX27 | ANTIINFECTIVES FOR SYSTEMIC USE |

| Drug Type (Second level) | Drug Name | ATC/DDD Code | First level |
|---|---|---|---|
| ANTIVIRALS FOR SYSTEMIC USE + ANTIVIRALS FOR SYSTEMIC USE + IMMUNOSTIMULANTS | Ribavirin + Lopinavir/Ritonavir + Interferon alpha | J05AP01 + J05AR10 + L03AB04 | ANTIINFECTIVES FOR SYSTEMIC USE + ANTIINFECTIVES FOR SYSTEMIC USE + ANTINEOPLASTIC AND IMMUNOMODULATING AGENTS |
| ANTIVIRALS FOR SYSTEMIC USE + ANTIVIRALS FOR SYSTEMIC USE + IMMUNOSTIMULANTS | Interferon beta + Lopinavir/ritonavir + Ribavirin | J05AP01 + J05AR10 + L03AB07 | ANTIINFECTIVES FOR SYSTEMIC USE + ANTIINFECTIVES FOR SYSTEMIC USE + ANTINEOPLASTIC AND IMMUNOMODULATING AGENTS |
| ANTIVIRALS FOR SYSTEMIC USE + IMMUNOSTIMULANTS | Ribavirin + Interferon alpha | J05AP01 + L03AB04 | ANTIINFECTIVES FOR SYSTEMIC USE + ANTINEOPLASTIC AND IMMUNOMODULATING AGENTS |
| ANTIVIRALS FOR SYSTEMIC USE + IMMUNOSTIMULANTS | Lopinavir/Ritonavir + Interferon Alpha | J05AR10 + L03AB04 | ANTIINFECTIVES FOR SYSTEMIC USE + ANTINEOPLASTIC AND IMMUNOMODULATING AGENTS |
| ANTIVIRALS FOR SYSTEMIC USE + IMMUNOSTIMULANTS | Lopinavir/ritonavir + Novaferon | J05AR10 + NA | ANTIINFECTIVES FOR SYSTEMIC USE + ANTINEOPLASTIC AND IMMUNOMODULATING AGENTS |
| BLOOD SUBSTITUTES AND PERFUSION SOLUTIONS | Convalescent plasma | B05AX03 (Blood plasma) | BLOOD AND BLOOD FORMING ORGANS |
| BLOOD SUBSTITUTES AND PERFUSION SOLUTIONS | Mesenchymal stem cells | B05AX04 | BLOOD AND BLOOD FORMING ORGANS |
| BLOOD SUBSTITUTES AND PERFUSION SOLUTIONS | Fresh frozen plasma | B05AX03 (Blood plasma) | BLOOD AND BLOOD FORMING ORGANS |
| CALCIUM CHANNEL BLOCKERS | Auxora | NA | CARDIOVASCULAR SYSTEM |
| CORTICOSTEROIDS, DERMATOLOGICAL PREPARATIONS | Methylprednisolone | D07AA01 | DERMATOLOGICALS |
| CORTICOSTEROIDS, DERMATOLOGICAL PREPARATIONS | Dexamethasone | D07AB19 | DERMATOLOGICALS |
| CORTICOSTEROIDS, DERMATOLOGICAL PREPARATIONS | Hydrocortisone | D07AA02 | DERMATOLOGICALS |
| COUGH AND COLD PREPARATIONS | N-acetylcysteine | R05CB01 | RESPIRATORY SYSTEM |
| COUGH AND COLD PREPARATIONS | Bromhexine | R05CB02 | RESPIRATORY SYSTEM |
| IMMUNE SERA AND IMMUNOGLOBULINS | Intravenous Immunoglobulin | J06B | ANTIINFECTIVES FOR SYSTEMIC USE |
| IMMUNOSTIMULANTS | IFX | NA | ANTINEOPLASTIC AND IMMUNOMODULATING AGENTS |
| IMMUNOSTIMULANTS | Interleukin 7 | L03AC | ANTINEOPLASTIC AND IMMUNOMODULATING AGENTS |
| IMMUNOSTIMULANTS | Interferon beta | L03AB07, L03AB08 | ANTINEOPLASTIC AND IMMUNOMODULATING AGENTS |
| IMMUNOSTIMULANTS | Novaferon | NA | ANTINEOPLASTIC AND IMMUNOMODULATING AGENTS |

| **Drug Type (Second level)** | **Drug Name** | **ATC/DDD Code** | **First level** |
|---|---|---|---|
| IMMUNOSTIMULANTS | Interferon alpha | L03AB04, L03AB05 | ANTINEOPLASTIC AND IMMUNOMODULATING AGENTS |
| IMMUNOSTIMULANTS | Interferon alpha + Interferon gamma | L03AB04 + L03AB03 | ANTINEOPLASTIC AND IMMUNOMODULATING AGENTS |
| IMMUNOSTIMULANTS | Interferon kappa + TFF2 | NA | ANTINEOPLASTIC AND IMMUNOMODULATING AGENTS |
| IMMUNOSTIMULANTS | Interferon gamma | L03AB03 | ANTINEOPLASTIC AND IMMUNOMODULATING AGENTS |
| IMMUNOSTIMULANTS | Recombinant human GCSF | L03AA | ANTINEOPLASTIC AND IMMUNOMODULATING AGENTS |
| IMMUNOSTIMULANTS | Intravenous CIGB-325 | NA | ANTINEOPLASTIC AND IMMUNOMODULATING AGENTS |
| IMMUNOSUPPRESSANTS | Tocilizumab | L04AC07 | ANTINEOPLASTIC AND IMMUNOMODULATING AGENTS |
| IMMUNOSUPPRESSANTS | Leflunomide | L04AA13 | ANTINEOPLASTIC AND IMMUNOMODULATING AGENTS |
| IMMUNOSUPPRESSANTS + IMMUNOSTIMULANTS | Leflunomide + Interferon Alpha | L04AA13 + L03AB04 | ANTINEOPLASTIC AND IMMUNOMODULATING AGENTS + ANTINEOPLASTIC AND IMMUNOMODULATING AGENTS |
| OTHER ALIMENTARY TRACT AND METABOLISM PRODUCTS | Alpha-Lipoic acid | A16AX01 | ALIMENTARY TRACT AND METABOLISM |
| PSYCHOANALEPTICS | Fluvoxamine | N06AB08 | NERVOUS SYSTEM |
| VITAMINS | Calcifediol | A11CC06 | ALIMENTARY TRACT AND METABOLISM |
| VITAMINS | Vitamin D3 | A11CC | ALIMENTARY TRACT AND METABOLISM |
| IMMUNOSUPPRESSANTS + ANTIVIRALS FOR SYSTEMIC USE | Baricitinib + Remdesivir | L04AA37 + NA | ANTINEOPLASTIC AND IMMUNOMODULATING AGENTS + ANTIINFECTIVES FOR SYSTEMIC USE |
| ANTITHROMBOTIC AGENTS | Sulodexide | B01AB11 | BLOOD AND BLOOD FORMING ORGANS |

#### Table S3. Detailed trial characteristics

| Registration number | Publication status (Published 1 or preprinted 0) | Study status (Completed; Interim analysis; Terminated early) | Randomization (1=double-blinded; 2=single-blined;3=open-label) | isMultiarm | Control type (1: compared with SOC; 2: compared with other interventions; 3: compared to both other interventions and SOC) | Design (1=parallel group 2=cluster randomized) | Funding resources(1=Industry 2=Government 3=Institutional 4=Not-for-profit foundation 0=None) | Intervention (Details) | Geographies | No. of patients |
|---|---|---|---|---|---|---|---|---|---|---|
| NCT04384380 | 1 | Completed | 3 | 0 | 1 | 1 | 2 | Hydroxychloroquine vs SOC | China | 33 |
| NCT04261517 | 1 | Completed | 3 | 0 | 1 | 1 | 2 | Hydroxychloroquine vs SOC | China | 30 |
| jRCTs041190120 | 1 | Completed | 3 | 0 | 2 | 1 | 1 | Early favipiravir vs late favipiravir | Japan | 88 |
| IRCT20100228003449N28 | 1 | Completed | 3 | 0 | 1 | 1 | 0 | Interferon beta-1a vs SOC | Iran | 81 |
| NCT04383535 | 1 | Completed | 1 | 0 | 1 | 1 | 3 | Convalescent plasma vs SOC | Europe | 333 |
| ChiCTR2000029853 | 1 | Completed | 3 | 0 | 1 | 1 | NA | Azvudine vs SOC | China | 20 |
| ChiCTR2000029851 | 0 | Completed | 2 | 0 | 1 | 1 | NA | Alpha-Lipoic acid vs SOC | China | 17 |
| ChiCTR2000029308 | 1 | Completed | 3 | 0 | 1 | 1 | 2 | lopinavir–ritonavir vs SOC | China | 199 |
| ChiCTR2000030262 | 1 | Completed | 3 | 0 | 1 | 1 | 2 | Interferon kappa + TFF2 vs SOC | China | 80 |
| NCT04333420 | 0 | Completed | 3 | 0 | 1 | 1 | 1 | IFX-1 vs SOC | Netherland | 30 |
| NCT04468646 | 0 | Completed | 3 | 0 | 1 | 1 | 0 | Aprepitant vs SOC | Pakistan | 18 |
| NCT04345614 | 1 | Completed | 3 | 0 | 1 | 1 | 1 | Auxora vs SOC | USA | 30 |
| NCT04434248 | 1 | Interim analysis | 3 | 0 | 1 | 1 | 2 | Favipiravir vs SOC | Russia | 60 |
| NCT04321278 | 1 | Completed | 3 | 0 | 1 | 1 | 3 | Azithromycin vs SOC | Brazil | 397 |
| ChiCTR2000029544 | 1 | Completed | 3 | 1 | 3 | 1 | NA | Baloxavir marboxil vs favipiravir vs SOC | China | 29 |
| CTRI202004024775 | 1 | Completed | 3 | 0 | 1 | 1 | 2 | Convalescent plasma vs SOC | India | 464 |
| NCT04381936_Dex | 1 | Interim analysis | 3 | 0 | 1 | 1 | 2 | Dexamethasone vs SOC | UK | 6424 |
| RBR8969zg | 1 | Completed | 1 | 0 | 1 | 1 | 2 | N-acetylcysteine vs SOC | Brazil | 135 |

| Registration number | Publication status (Published 1 or preprinted 0) | Study status (Completed; Interim analysis; Terminated early) | Randomization (1=double-blinded; 2=single-blined;3=open-label) | isMultiarm | Control type (1: compared with SOC; 2: compared with other interventions; 3: compared to both other interventions and SOC) | Design (1=parallel group 2=cluster randomized) | Funding resources(1=Industry 2=Government 3=Institutional 4=Not-for-profit foundation 0=None) | Intervention (Details) | Geographies | No. of patients |
|---|---|---|---|---|---|---|---|---|---|---|
| RPCEC00000307 | 0 | Interim analysis | 3 | 0 | 2 | 1 | NA | Interferon alpha-2b + interferon-gamma vs Interferon alpha-2b | Cuba | 63 |
| NCT04366908 | 1 | Completed | 3 | 0 | 1 | 1 | NAA | Calcifediol vs SOC | Spain | 76 |
| NCT04326790 | 1 | Completed | 3 | 0 | 1 | 1 | 3 | Colchicine vs SOC | Greece | 105 |
| ChiCTR2000029757 | 1 | Completed | 3 | 0 | 1 | 1 | 2 | Convalescent plasma vs SOC | China | 103 |
| NCT04327401 | 1 | Completed | 3 | 0 | 1 | 1 | NA | Dexamethasone vs SOC | Brazil | 299 |
| NCT042323527 | 1 | Completed | 1 | 0 | 2 | 1 | 2 | High-dose chloroquine vs low-dose chloroquine | Brazil | 81 |
| NCT02517489 | 1 | Terminated early | 1 | 0 | 1 | 1 | 2 | Hydrocortisone vs SOC | France | 149 |
| NCT02735707 | 1 | Terminated early | 3 | 0 | 1 | 1 | 2 | Hydrocortisone vs SOC | Multisite | 384 |
| NCT04381936_Hydro | 1 | Completed | 3 | 0 | 1 | 1 | 2 | Hydroxychloroquine vs SOC | UK | 4716 |
| NCT04332991 | 1 | Completed | 1 | 0 | 1 | 1 | 2 | Hydroxychloroquine vs SOC | USA | 479 |
| ChiCTR2000030007 | 1 | Completed | 3 | 0 | 1 | 1 | 1, 2 | Recombinant human granulocyte colony-stimulating factor vs SOC | China | 200 |
| NCT04292730 | 1 | Completed | 3 | 0 | 1 | 1 | 1 | Remdesivir vs SOC | Multisite | 584 |
| NCT04346355 | 1 | Completed | 3 | 0 | 1 | 1 | 2 | Tocilizumab vs SOC | Italy | 126 |
| NCT04331808 | 1 | Completed | 3 | 0 | 1 | 1 | 2 | Tocilizumab vs SOC | France | 130 |
| NCT04449718 | 0 | Completed | 1 | 0 | 1 | 1 | 2 | Vitamin D3 vs SOC | Brazil | 240 |
| ChiCTR2000030054 | 0 | Terminated early | 3 | 1 | 3 | 1 | 2 | Hydroxychloroquine vs Chloroquine vs SOC | China | 48 |
| NCT04252885 | 1 | Completed | 2 | 1 | 3 | 1 | 2 | Lopinavir/ritonavir vs Arbidol vs SOC | China | 86 |

| Registration number | Publication status (Published 1 or preprinted 0) | Study status (Completed; Interim analysis; Terminated early) | Randomization (1=double-blinded; 2=single-blined;3=open-label) | isMultiarm | Control type (1: compared with SOC; 2: compared with other interventions; 3: compared to both other interventions and SOC) | Design (1=parallel group 2=cluster randomized) | Funding resources(1=Industry 2=Government 3=Institutional 4=Not-for-profit foundation 0=None) | Intervention (Details) | Geographies | No. of patients |
|---|---|---|---|---|---|---|---|---|---|---|
| ChiCTR20000300001 | 1 | Terminated early | 1 | 0 | 1 | 1 | NA | Triazavirin vs SOC | China | 52 |
| ChiCTR2000029559 | 0 | Completed | 3 | 0 | 1 | 1 | 2 | Hydroxychloroquine vs SOC | China | 62 |
| NCT04356937 | 1 | Completed | 3 | 0 | 1 | 1 | 1 | Tocilizumab vs SOC | USA | 243 |
| IRCT20151227025726N20 | 1 | Completed | 3 | 0 | 1 | 1 | NA | Intravenous Immunoglobulin vs SOC | Iran | 84 |
| IRCT20200328046886N1 | 1 | Completed | 3 | 0 | 1 | 1 | 3 | Sofosbuvir + Daclatasvir + Ribavirin vs SOC | Iran | 48 |
| ChiCTR2000030254 | 0 | Completed | 3 | 0 | 2 | 1 | 2 | Favipiravir vs Arbidol | China | 236 |
| IRCT2019072704434N1 | 1 | Completed | 3 | 0 | 2 | 1 | 3 | Febuxostat vs Hydroxychloroquine | Iran | 54 |
| NCT04342663 | 1 | Completed | 1 | 0 | 1 | 1 | 3 | Fluvoxamine vs SOC | USA | 152 |
| EudraCT202000193437 | 0 | Completed | 3 | 0 | 1 | 1 | 0 | Methylprednisolone vs SOC | Spain | 85 |
| NCT04304053 | 1 | Completed | 3 | 1 | 3 | 2 | 1,2,3 | Hydroxychloroquine vs Hydroxychloroquine + Darunavir vs SOC | Spain | 307 |
| NCT04308668 | 1 | Completed | 1 | 0 | 1 | 1 | Private donors | Hydroxychloroquine vs SOC | Multisite | 423 |
| ChiCTR2000029868 | 1 | Completed | 3 | 0 | 1 | 1 | 2 | Hydroxychloroquine vs SOC | China | 150 |
| NCT04353336 | 1 | Completed | 3 | 0 | 1 | 1 | NA | Hydroxychloroquine vs SOC | Egypt | 175 |
| NCT04329832 | 1 | Terminated early | 3 | 0 | 2 | 1 | 2,3 | Hydroxychloroquine vs Azithromycin | USA | 85 |
| NCT04322123 | 1 | Completed | 3 | 1 | 3 | 1 | 3 | Hydroxychloroquine + Azithromycin vs Hydroxychloroquine vs SOC | Brazil | 665 |
| IRCT20100228003449N27 | 1 | Completed | 3 | 0 | 1 | 1 | 0 | Interferon β-1b vs SOC | Iran | 66 |

| Registration number | Publication status (Published 1 or preprinted 0) | Study status (Completed ; Interim analysis; Terminated early) | Randomization (1=double-blinded; 2=single-blined;3=open-label) | isMultiarm | Control type (1: compared with SOC; 2: compared with other interventions; 3: compared to both other interventions and SOC) | Design (1=parallel group 2=cluster randomized) | Funding resources(1=Industry 2=Government 3=Institutional 4=Not-for-profit foundation 0=None) | Intervention (Details) | Geographies | No. of patients |
|---|---|---|---|---|---|---|---|---|---|---|
| NCT04381936_Lopi | 1 | Completed | 3 | 0 | 1 | 1 | 2 | Lopinavir/ritonavir vs SOC | UK | 5040 |
| NCT04343729 | 1 | Completed | 1 | 0 | 1 | 1 | 2, 3 | Methylprednisolone vs SOC | Brazil | 393 |
| ChiCTR2000029387 | 1 | Completed | 3 | 1 | 2 | 1 | 0 | Ribavirin + Interferon-Alpha vs Lopinavir/Ritonavir + Interferon-Alpha vs Ribavirin + Lopinavir/Ritonavir + Interferon-Alpha | China | 101 |
| mTcMDP | 0 | Completed | 3 | 0 | 1 | 1 | NA | 99mTc-MDP vs SOC | China | 21 |
| NCT04331899 | 0 | Completed | 2 | 0 | 1 | 1 | 1, 3 | Interferon lambda vs SOC | USA | 120 |
| NCT04354259 | 0 | Completed | 1 | 0 | 1 | 1 | 3 | Interferon lambda vs SOC | Canada | 60 |
| Oman | 1 | Completed | 3 | 0 | 2 | 1 | 0 | Favipiravir + Interferon beta vs Hydroxychloroquine | Oman | 89 |
| NCT04356534 | 0 | Completed | 3 | 0 | 1 | 1 | 2, 3 | Convalescent plasma vs SOC | Bahrain | 40 |
| NCT04349592 | 1 | Completed | 1 | 1 | 3 | 1 | 2 | Hydroxychloroquine vs Hydroxychloroquine + Azithromycin vs SOC | Qatar | 456 |
| NCT04280705 | 1 | Completed | 1 | 0 | 1 | 1 | 2 | Remdesivir vs SOC | Multisite | 1062 |
| NCT04257656 | 1 | Completed | 1 | 0 | 1 | 1 | 2 | Remdesivir vs SOC | China | 236 |
| NCT04315948 | 1 | Interim analysis | 3 | 1 | 3 | 1 | 3 | Hydroxychloroquine vs Interferon alpha-2b vs Lopinavir/ritonavir vs Remdesivir vs SOC | Multisite | 11266 |
| JAK | 1 | Completed | 2 | 0 | 1 | 1 | 2, 3 | Ruxolitinib vs SOC | China | 41 |

| Registration number | Publication status (Published 1 or preprinted 0) | Study status (Completed; Interim analysis; Terminated early) | Randomization (1=double-blinded; 2=single-blined;3=open-label) | isMultiarm | Control type (1: compared with SOC; 2: compared with other interventions; 3: compared to both other interventions and SOC) | Design (1=parallel group 2=cluster randomized) | Funding resources(1=Industry 2=Government 3=Institutional 4=Not-for-profit foundation 0=None) | Intervention (Details) | Geographies | No. of patients |
|---|---|---|---|---|---|---|---|---|---|---|
| ChiCTR2000029496 | 1 | Completed | 3 | 1 | 2 | 1 | 2 | Novaferon vs Lopinavir/ritonavir vs Lopinavir/ritonavir + Novaferon | China | 89 |
| IRCT20200128046294N2 | 1 | Completed | 3 | 0 | 1 | 1 | 3 | Sofosbuvir + Daclatasvir vs SOC | Iran | 66 |
| RBR949z6v | 1 | Completed | 3 | 0 | 2 | 1 | 0 | Prophylactic anticoagulation vs Therapeutic enoxaparin | Brazil | 20 |
| ChiCTR2000030058 | 1 | Completed | 3 | 0 | 2 | 1 | 2 | Leflunomide + IFN alpha-2a vs IFN alpha-2a | China | 48 |
| NCT04276688 | 1 | Completed | 3 | 0 | 2 | 1 | 4 | Interferon beta-1b + Lopinavir–ritonavir + Ribavirin vs Lopinavir–ritonavir | China | 127 |
| NCT04447534 | 1 | Completed | 1 | 0 | 2 | 2 | NA | Zinc + HCQ vs HCQ | Egypt | 191 |
| NCT04385095 | 1 | Completed | 1 | 0 | 1 | 1 | 1 | Interferon beta vs SOC | UK | 98 |
| NCT04346446 | 0 | Completed | 3 | 0 | 2 | 1 | 0 | Convalescent plasma vs fresh frozen plasma | India | 29 |
| NCT04591600 | 0 | Completed | 3 | 0 | 1 | 1 | NA | Ivermectin + Doxycycline vs SOC | Iraq | 140 |
| NCT04372186 | 0 | Completed | 1 | 0 | 1 | 1 | 1 | Tocilizumab vs SOC | Multisite | 377 |
| IRCT20200501047259N1 | 1 | Completed | 1 | 0 | 1 | 1 | 3 | Intravenous immunoglobulin vs SOC | Iran | 59 |
| NCT04552483 | 0 | Completed | 1 | 0 | 1 | 1 | 2 | Nitazoxanide vs SOC | Brazil | 392 |
| NCT04325893 | 0 | Completed | 1 | 0 | 1 | 1 | 2 | Hydroxychloroquine vs SOC | France | 250 |
| NCT04288102 | 0 | Completed | 1 | 0 | 1 | 1 | 2 | Mesenchymal stem cells vs SOC | China | 100 |

| Registration number | Publication status (Published 1 or preprinted 0) | Study status (Completed; Interim analysis; Terminated early) | Randomization (1=double-blinded; 2=single-blined;3=open-label) | isMultiarm | Control type (1: compared with SOC; 2: compared with other interventions; 3: compared to both other interventions and SOC) | Design (1=parallel group 2=cluster randomized) | Funding resources(1=Industry 2=Government 3=Institutional 4=Not-for-profit foundation 0=None) | Intervention (Details) | Geographies | No. of patients |
|---|---|---|---|---|---|---|---|---|---|---|
| Umi | 1 | Completed | 3 | 0 | 1 | 1 | 0 | umifenovir vs SOC | Kyrgyzstan | 30 |
| IRCT202003117046797N4 | 1 | Completed | 3 | 0 | 1 | 1 | 3 | Bromhexine vs SOC | Iran | 78 |
| NCT04369742 | 1 | Completed | 1 | 0 | 1 | 1 | 2, 3 | Hydroxychloroquine vs SOC | USA | 128 |
| NCT04375098 | 0 | Completed | 3 | 0 | 2 | 1 | 4 | Early plasma vs deferred plasma | Chile | 58 |
| RPCEC00000317-En | 0 | Completed | 3 | 0 | 1 | 1 | NA | Intravenous CIGB-325 vs SOC | Cuba | 20 |
| NCT04273763 | 1 | Completed | 3 | 0 | 1 | 1 | 2 | Bromhexine vs SOC | China | 18 |
| NCT04320615 | 0 | Completed | 1 | 0 | 1 | 1 | 2 | Tocilizumab vs SOC | Multisite | 438 |
| NCT04345523 | 0 | Terminated early | 3 | 0 | 1 | 1 | 3 | Convalescent Plasma vs SOC | Spain | 81 |
| RBR-8jyhx | 0 | Interim analysis | 1 | 0 | 1 | 1 | 2 | Colchicine vs SOC | Brazil | 35 |
| NCT0449199 | 0 | Completed | 3 | 0 | 1 | 2 | 0 | Hydroxychloroquine vs SOC | Pakistan | 500 |
| NCT04411667 | 0 | Completed | 3 | 0 | 1 | 1 | 1 | Intravenous immunoglobulin vs SOC | USA | 33 |
| Linazi | 1 | Completed | 3 | 0 | 2 | 1 | 0 | LINCOCIN® vs AZITRO® | Turkey | 24 |
| NCT04292899 | 1 | Completed | 3 | 0 | 2 | 1 | 1 | 5-day Remdesivirv 10-day Remdesivir | Multisite | 397 |
| Iver | 1 | Completed | 1 | 1 | 3 | 1 | 1 | Ivermectin vs Ivermectin + Doxycycline vs SOC | Bangladesh | 72 |
| NCT04381936_Azi | 0 | Interim analysis | 3 | 0 | 1 | 1 | 2,3 | Azithromycin vs SOC | UK | 7764 |
| NCT04401579 | 1 | Completed | 1 | 0 | 2 | 1 | 2 | Baricitinib + Remdesivir vs Remdesivir | Multisite | 1033 |
| IRCT20180725040596N2 | 1 | Completed | 3 | 0 | 2 | 1 | 3 | Hydroxychloroquine + Lopinavir/Ritonavir | Iran | 100 |

| Registration number | Publication status (Published 1 or preprinted 0) | Study status (Completed; Interim analysis; Terminated early) | Randomization (1=double-blinded; 2=single-blined;3=open-label) | isMultiarm | Control type (1: compared with SOC; 2: compared with other interventions; 3: compared to both other interventions and SOC) | Design (1=parallel group 2=cluster randomized) | Funding resources(1=Industry 2=Government 3=Institutional 4=Not-for-profit foundation 0=None) | Intervention (Details) | Geographies | No. of patients |
|---|---|---|---|---|---|---|---|---|---|---|
| | | | | | | | | vs Hydroxychloroquine + Arbidol | | |
| ISRCTN59048638 | 0 | Completed | 1 | 0 | 1 | 1 | 1 | Sulodexide vs SOC | Mexico | 243 |

#### Table S4. Detailed patient characteristics for included studies.

| Registration number | Age | Male % | Mechanical ventilation at baseline (%) | Patient Type (Mild , Moderate, Severe, Critical) | Severity | Current or unspecified smokers (%) | Former smokers (%) | Pregnant (%) | Inpatient (%) | Confirmed COVID-19 (%) |
|---|---|---|---|---|---|---|---|---|---|---|
| NCT04384380 | Mean (SD) 32.9 (10.7) | 57.6 | NA | Mild (87.9); Moderate (12.1) | 0.00 | NA | NA | 0.0 | 100 | 100 |
| NCT04261517 | Mean (SD) 48.6 (4.1) | 70.0 | NA | Moderate (100) | 0.00 | NA | NA | 0.0 | 100 | 100 |
| jRCTs041190120 | Median (IQR) 50 (38 to 64.5) | 61.4 | NA | Mild (100) | 0.00 | NA | NA | 0.0 | 100 | 100 |
| IRCT20100228003449N28 | Mean (SD) 57.7 (15.1) | 54.3 | NA | Severe (100) | 1.00 | NA | NA | 0.0 | 100 | 100 |
| NCT04383535 | Median (IQR) 62.5 (53 to 72.5); 62 (49 to 71) | 67.6 | 0.0 | Severe (100) | 1.00 | NA | NA | 0.0 | 100 | 100 |
| ChiCTR2000029853 | Median (IQR) 52 (17 to 76) | 60.0 | 0.0 | Mild (15); Moderate (85) | 0.00 | 25.0 | NA | 0.0 | 100 | 100 |
| ChiCTR2000029851 | Median (IQR) 63 (59 to 66) | 76.4 | 94.1 | Critical | 1.00 | NA | NA | 0.0 | 100 | 100 |
| ChiCTR2000029308 | Median (IQR) 58 (49 to 68) | 60.3 | 16.1 | Severe | 1.00 | NA | NA | 0.0 | 100 | 100 |
| ChiCTR2000030262 | Mean (SD) 35.35 (11.2) | 63.8 | NA | Moderate | 0.00 | NA | NA | 0.0 | 100 | 100 |
| NCT04333420 | Mean (SD) 60 (9) | 73.0 | NA | Severe | 1.00 | NA | NA | 0.0 | 100 | 100 |
| NCT04468646 | Mean (SD) 53.5 (12.7) | 61.0 | NA | Severe/Critical | 1.00 | NA | NA | 0.0 | 100 | 100 |
| NCT04345614 | Mean (SD) 59.3 (12.7) | 46.7 | NA | Severe/Critical | 1.00 | NA | NA | NA | 100 | 100 |
| NCT04434248 | NA | NA | NA | Moderate | 0.00 | NA | NA | 0.0 | 100 | 100 |

| Registration number | Age | Male % | Mechanical ventilation at baseline (%) | Patient Type (Mild , Moderate, Severe, Critical) | Severity | Current or unspecified smokers (%) | Former smokers (%) | Pregnant (%) | Inpatient (%) | Confirmed COVID-19 (%) |
|---|---|---|---|---|---|---|---|---|---|---|
| NCT04321278 | Median (IQR) 59.4 (49.3 to 70.0); 60.2 (52.0 to 70.1) | 66.0 | 49.4 | Severe | 1.00 | 9.1 | NA | NA | 100 | 100 |
| ChiCTR2000029544 | Mean (SD) 52.5 (12.5) | 72.4 | NA | NA | 0.50 | NA | NA | NA | 100 | 100 |
| CTRI202004024775 | Median (IQR) 52 (42 to 60); 52 (41, 60) | 76.3 | NA | Moderate | 0.00 | 8.0 | | 0.0 | 100 | 100 |
| NCT04381936_Dex | Mean (SD) 66.2 (15.7) | 63.6 | 76.1 | NA | 0.50 | NA | NA | 0.0 | 100 | 100 |
| RBR8969zg | Median (IQR) 59 (47 to 70); 58 (48 to 70) | 59.3 | NA | Severe | 1.00 | NA | NA | 0.0 | 100 | 100 |
| RPCEC00000307 | Median (IQR) 38 (19 to 82) | 54.0 | NA | Mild/Moderate | 0.00 | 12.7 | NA | 0.0 | 100 | 100 |
| NCT04366908 | Mean (SD) 53.0 (10.2) | 59.2 | NA | NA | 0.50 | NA | NA | 0.0 | 100 | 100 |
| NCT04326790 | Median (IQR) 65 (54 to 80); 63 (55 to 70) | 58.1 | 0.0 | Mild/Moderate | 0.00 | 4.4 | 22.2 | 0.0 | 100 | 100 |
| ChiCTR2000029757 | Median (IQR) 70 (62 to 80); 69 (63 to 76) | 58.3 | 96.1 | Severe | 1.00 | NA | NA | 0.0 | 100 | 100 |
| NCT04327401 | Mean (SD) 61.4 (14.6) | 62.5 | 100.0 | Severe | 1.00 | 4.3 | NA | 0.0 | 100 | 100 |
| NCT042323527 | Mean (SD) 51.1 (13.9) | 75.3 | NA | Severe | 1.00 | 8.3 | 22.9 | 2.5 | 100 | 100 |
| NCT02517489 | Median (IQR) 63.1 (51.5 to 70.8); 66.3 (53.5 to 72.7) | 69.8 | 81.2 | Severe | 1.00 | 23.9 | | NA | 100 | 100 |
| NCT02735707 | Mean (SD) 59.9 (12.8) | 71.1 | 85.2 | Severe | 1.00 | NA | NA | NA | 100 | 100 |

| Registration number | Age | Male % | Mechanical ventilation at baseline (%) | Patient Type (Mild , Moderate, Severe, Critical) | Severity | Current or unspecified smokers (%) | Former smokers (%) | Pregnant (%) | Inpatient (%) | Confirmed COVID-19 (%) |
|---|---|---|---|---|---|---|---|---|---|---|
| NCT04381936_Hydro | Mean (SD) 65.3 (15.3) | 62.2 | 76.4 | NA | 0.50 | NA | NA | 0.0 | 100 | 91 |
| NCT04332991 | Median (IQR) 58 (45 to 69); 57 (43 to 68) | 55.7 | 64.9 | Mild/Moderate (81.8) Severe (18.2) | 0.18 | NA | NA | NA | 100 | 100 |
| ChiCTR2000030007 | Median (IQR) 45 (40 to 55) | 56.0 | 87.0 | Mild/Moderate (81.8) Severe (18.2) | 0.00 | NA | NA | 0.0 | 100 | 100 |
| NCT04292730 | Median (IQR) 56 (45 to 66); 58 (48 to 66); 57 (45 to 66) | 60.8 | 15.9 | Moderate | 0.00 | NA | NA | NA | 100 | 100 |
| NCT04346355 | Median (IQR) 60.0 (53.0 to 72.0) | 61.1 | 0.0 | Mild/Moderate (81.8) Severe (18.2) | 0.00 | NA | NA | NA | 100 | 100 |
| NCT04331808 | Median (IQR) 64.0 (57.1 to 74.3) vs 63.3 (57.1 to 72.3) | 67.7 | NA | Moderate/Severe | 0.50 | 2.3 | 6.3 | NA | 100 | 100 |
| NCT04449718 | Mean (SD) 56.3 (14.6) | 56.3 | NA | Severe | 1.00 | NA | NA | 0.0 | 100 | 100 |
| ChiCTR2000030054 | Mean (SD) 46.9 (14.3) | 45.8 | NA | Mild/Moderate (81.8) Severe (18.2) | 0.00 | NA | NA | NA | 100 | 100 |
| NCT04252885 | Mean (Range) 50.7 (17 to 19); 50.5 (20 to 74); 44.3 (27 to 62) | 46.5 | 15.4 | Mild/Moderate (81.8) Severe (18.2) | 0.00 | NA | NA | 0.0 | 100 | 100 |
| ChiCTR20000300001 | 58 [48, 65] | 50.0 | NA | NA | 0.50 | 11.5 | NA | 0.0 | 100 | 100 |

| Registration number | Age | Male % | Mechanical ventilation at baseline (%) | Patient Type (Mild , Moderate, Severe, Critical) | Severity | Current or unspecified smokers (%) | Former smokers (%) | Pregnant (%) | Inpatient (%) | Confirmed COVID-19 (%) |
|---|---|---|---|---|---|---|---|---|---|---|
| ChiCTR2000029559 | Mean (SD) 44.7 (15.3) | 46.8 | NA | Mild/Moderate (81.8) Severe (18.2) | 0.00 | NA | NA | 0.0 | 100 | 100 |
| NCT04356937 | Median (IQR) 59.8 (45.3 to 69.4) | 58.0 | 0.0 | Moderate | 0.00 | 3.0 | 30.0 | NA | 100 | 100 |
| IRCT20151227025726N20 | Mean (SD) 53.6 (13.5) | 77.4 | NA | Severe | 1.00 | 1.0 | NA | 0.0 | 100 | 100 |
| IRCT20200328046886N1 | Median (IQR) 45 (38 to 69); 60 (47.5 to 68.5) | 37.5 | NA | Moderate | 0.00 | NA | NA | 0.0 | 100 | 100 |
| ChiCTR2000030254 | NA | 46.6 | NA | Moderate (88.6); Severe/Critical (11.4) | 0.11 | NA | NA | 0.0 | 100 | 100 |
| IRCT2019072704434N1 | Mean (SD) 57.7 (9.3) | 59.3 | NA | Mild/Moderate (81.8) Severe (18.2) | 0.00 | 1.9 | NA | NA | 100 | NA |
| NCT04342663 | Median (IQR) 46 (35 to 58); 45 (36 to 54) | 36.8 | NA | Mild | 0.00 | NA | NA | NA | 0 | 100 |
| EudraCT202000193437 | Mean (SD) 69 (12) | 58.0 | NA | Moderate/Severe | 0.50 | NA | NA | 0.0 | 100 | 100 |
| NCT04304053 | Mean (SD) 42.0 (12.8) | 71.0 | 0.0 | Mild/Moderate (81.8) Severe (18.2) | 0.00 | NA | NA | 0.0 | 0 | 100 |
| NCT04308668 | Median (IQR) 41 (33 to 49); 39 (31 to 50) | 43.7 | NA | Mild | 0.00 | 4.0 | NA | 0.0 | 0 | 100 |

| Registration number | Age | Male % | Mechanical ventilation at baseline (%) | Patient Type (Mild , Moderate, Severe, Critical) | Severity | Current or unspecified smokers (%) | Former smokers (%) | Pregnant (%) | Inpatient (%) | Confirmed COVID-19 (%) |
|---|---|---|---|---|---|---|---|---|---|---|
| ChiCTR2000029868 | Mean (SD) 46.1 (14.7) | 55.0 | NA | Mild(15); Moderate(84); Severe (1) | 0.00 | NA | NA | 0.0 | 100 | 100 |
| NCT04353336 | Mean (SD) 40.72 (19.32) | 58.8 | NA | NA | 0.50 | 31.4 | NA | 0.0 | 100 | 100 |
| NCT04329832 | Median (IQR) 55 (42 to 65) | 61.0 | 16.0 | Moderate(69); Severe(31) | 0.31 | NA | NA | NA | 100 | 100 |
| NCT04322123 | Mean (SD) 50.3 (14.6) | 58.3 | 0.0 | Mild/Moderate (81.8) Severe (18.2) | 0.00 | 6.6 | NA | NA | 100 | 76 |
| IRCT20100228003449N27 | Median (IQR) 60 (47 to 73); 61 (50 to 71) | 59.1 | 1.5 | Severe | 1.00 | NA | NA | 0.0 | 100 | 100 |
| NCT04381936_Lopi | Mean (SD) 66.3 (15.9) | 61.1 | NA | NA | 0.50 | NA | NA | 0.1 | 100 | 100 |
| NCT04343729 | Mean (SD) 55 (15) | 64.6 | 81.6 | NA | 0.50 | NA | NA | 0.0 | 100 | NA |
| ChiCTR2000029387 | Mean (SD) 42.5 (11.5) | 46.0 | 0.0 | Mild/Moderate (81.8) Severe (18.2) | 0.00 | NA | NA | 0.0 | 100 | 100 |
| mTcMDP | Median (IQR) 61 (47 to 67) | 42.9 | NA | Mild | 0.00 | NA | NA | NA | 100 | 100 |
| NCT04331899 | Median (IQR) 36 (18 to 71) | 58.3 | NA | Mild/Moderate (81.8) Severe (18.2) | 0.00 | NA | NA | 0.0 | 0 | 100 |
| NCT04354259 | Median (IQR) 48 (30 to 53); 39 (33 to 55) | 41.7 | NA | Moderate | 0.00 | NA | NA | 0.0 | 0 | 100 |

| Registration number | Age | Male % | Mechanical ventilation at baseline (%) | Patient Type (Mild , Moderate, Severe, Critical) | Severity | Current or unspecified smokers (%) | Former smokers (%) | Pregnant (%) | Inpatient (%) | Confirmed COVID-19 (%) |
|---|---|---|---|---|---|---|---|---|---|---|
| Oman | Mean (SD) 55 (14) | 58.0 | NA | Moderate/Severe | 0.50 | NA | NA | 0.0 | 100 | 100 |
| NCT04356534 | Mean (SD) 51.7 (13.6) | 80.0 | NA | Severe | 1.00 | 0.0 | NA | NA | 100 | 100 |
| NCT04349592 | Median (IQR) 42 (38 to 48); 40 (31 to 47); 41 (31 to 47) | 98.5 | NA | Mild/Moderate (81.8) Severe (18.2) | 0.00 | NA | NA | 0.0 | 0 | 100 |
| NCT04280705 | Mean (SD) 58.9 (15.0) | 64.4 | 45.0 | Mild/Moderate (15); Severe (85) | 0.85 | NA | NA | NA | 100 | 100 |
| NCT04257656 | Median (IQR) 66 (57 to 73); 64 (53 to 70) | 59.3 | 16.1 | Severe | 1.00 | NA | NA | 0.0 | 100 | 100 |
| NCT04315948 | NA | 62.0 | 8.0 | NA | 0.50 | 7.0 | NA | NA | 100 | 100 |
| JAK | Median (IQR) 63 (58 to 68) | 58.5 | 12.2 | Severe | 1.00 | 9.8 | NA | 0.0 | 100 | 100 |
| ChiCTR2000029496 | Median (IQR) 46.5 (40.0 to 63.8); 50.0 (37.8 to 62.8); 37.0 (26.0 to 54.0) | 47.2 | NA | Mild/Moderate (95); Severe (5) | 0.05 | NA | NA | NA | 100 | 100 |
| IRCT20200128046294N2 | Median (IQR) 58 (38 to 65); 62 (49 to 70) | 51.5 | NA | Moderate/Severe | 0.50 | NA | NA | NA | 100 | 100 |
| RBR949z6v | Mean (SD) 56.5 (13.1) | 80.0 | 100.0 | Severe | 1.00 | NA | NA | 0.0 | 100 | 100 |

| Registration number | Age | Male % | Mechanical ventilation at baseline (%) | Patient Type (Mild , Moderate, Severe, Critical) | Severity | Current or unspecified smokers (%) | Former smokers (%) | Pregnant (%) | Inpatient (%) | Confirmed COVID-19 (%) |
|---|---|---|---|---|---|---|---|---|---|---|
| ChiCTR2000030058 | Median (IQR) 56.0 (43.0 to 67.3); 55.5 (47.8 to 66.5) | 45.8 | NA | Mild/moderate (39); severe (9) | 0.19 | NA | NA | 0.0 | 100 | 100 |
| NCT04276688 | Median (IQR) 51.0 (31.0 to 61.3); 52.0 (33.5 to 62.5) | 53.5 | NA | Mild/Moderate | 0.00 | 5.5 | NA | 0.0 | 100 | 100 |
| NCT04447534 | Mean (SD) 43.56 (13.88) | 60.7 | NA | Mild/Moderate (70); Severe (30) | 0.30 | 42.4 | NA | 0.0 | 100 | 100 |
| NCT04385095 | Mean (SD) 57.1 (13.2) | 59.2 | 2.0 | NA | 0.50 | 2.0 | 27.6 | 0.0 | 100 | 100 |
| NCT04346446 | Mean (SD) 48.2 (9.8) | 75.9 | NA | Severe | 1.00 | NA | NA | 0.0 | 100 | 100 |
| NCT04591600 | Mean (SD) 48.7 (8.6) | 52.0 | NA | 22/70 severe | 0.31 | NA | NA | 0.0 | 100 | 100 |
| NCT04372186 | Mean (SD) 55.9 (14.4) | 59.2 | 26.5 | Moderate | 0.00 | 5.8 | 17.0 | NA | 100 | 100 |
| IRCT20200501047259N1 | Median (IQR) 56 (46 to 62) | 69.5 | NA | Severe | 1.00 | NA | NA | NA | 100 | 100 |
| NCT04552483 | NA | 46.9 | NA | Mild | 0.00 | NA | NA | NA | NA | 100 |
| NCT04325893 | Median (IQR) 77 (58 to 86) | 48.4 | 60.4 | Mild | 0.00 | 2.4 | NA | 0.0 | 100 | 100 |
| NCT04288102 | Mean (SD) 60.45 (8.66) | 56.0 | 1.0 | Severe | 1.00 | NA | NA | 0.0 | 100 | 100 |
| Umi | Mean (SD) 36.5 (12.1) | 60.0 | NA | Mild | 0.00 | NA | NA | 0.0 | 100 | 100 |
| IRCT202003117046797N4 | Mean (SD) 59.8 (14.9) | 56.4 | NA | NA | 0.50 | NA | NA | 0.0 | 100 | 100 |

| Registration number | Age | Male % | Mechanical ventilation at baseline (%) | Patient Type (Mild , Moderate, Severe, Critical) | Severity | Current or unspecified smokers (%) | Former smokers (%) | Pregnant (%) | Inpatient (%) | Confirmed COVID-19 (%) |
|---|---|---|---|---|---|---|---|---|---|---|
| NCT04369742 | Mean (SD) 66.2 (16.2) | 59.4 | 0.8 | Mild/Moderate(35); Severe (65) | 0.65 | 6.2 | 28.1 | 0.0 | 100 | 100 |
| NCT04375098 | Mean (Range) 65.8 (27 to 92) | 50.0 | NA | NA | 0.50 | NA | NA | 0.0 | 100 | 100 |
| RPCEC00000317-En | Mean (SD) 45.35 (12.0) | 70.0 | NA | Mild/Moderate (90); Severe (10) | 0.10 | NA | NA | NA | 100 | 100 |
| NCT04273763 | Median (IQR) 53 (50 to 62); 48 (32 to 51) | 77.8 | NA | NA | 0.50 | NA | NA | 0.0 | 100 | 100 |
| NCT04320615 | Mean (SD) 60.8 (14.3) | 69.9 | 37.7 | Severe | 1.00 | NA | NA | NA | 100 | 100 |
| NCT04345523 | Median 59 | 54.3 | 0.0 | NA | 0.50 | NA | NA | 0.0 | 100 | 100 |
| RBR-8jyhx | Median (IQR) 53.5 (35.5 to 65.5); 48.0 (41.5 to 64.0) | 40.0 | NA | Moderate/Severe | 0.50 | 20.0 | NA | 0.0 | 100 | 100 |
| NCT0449199 | Mean (SD) 35.96 (11.2) | 93.2 | NA | Mild | 0.00 | NA | NA | NA | 100 | 100 |
| NCT04411667 | Mean 54 | 60.6 | 0.0 | NA | 0.50 | 6.1 | 9.1 | NA | 100 | 100 |
| Linazi | Mean (SD) 58.75 (15.55) | 62.5 | 0.0 | NA | 0.50 | NA | NA | 0.0 | 100 | 100 |
| NCT04292899 | Median (IQR) 61 (50 to 69); 62 (50 to 71) | 63.7 | 30.7 | Severe | 1.00 | NA | NA | NA | 100 | 100 |
| Iver | Mean 42 | NA | 0.0 | Mild/Moderate | 0.00 | NA | NA | NA | 100 | 100 |
| NCT04381936_Azi | Mean (SD) 65.4 (15.6); 65.2 (15.7) | 62.1 | 0.1 | NA | 0.50 | NA | NA | 0.3 | 100 | 89 |

| Registration number | Age | Male % | Mechanical ventilation at baseline (%) | Patient Type (Mild , Moderate, Severe, Critical) | Severity | Current or unspecified smokers (%) | Former smokers (%) | Pregnant (%) | Inpatient (%) | Confirmed COVID-19 (%) |
|---|---|---|---|---|---|---|---|---|---|---|
| NCT04401579 | Mean (SD) 55.4 (15.7) | 63.1 | 31.7 | Moderate (68.3); Severe (31.7) | 0.32 | NA | NA | NA | 100 | 100 |
| IRCT20180725040596N2 | Mean (SD) 56.4 (16.3) | 60.0 | 5.0 | Mild (19); Moderate (58); Severe (23) | 0.23 | 15.0 | NA | 0.0 | 100 | 100 |
| ISRCTN59048638 | Mean (SD) 55.3 (10.3); 54.0 (10.9) | 0.5 | 0.0 | Mild | 0.00 | NA | NA | MA | 0 | 100 |

#### Table S5. Evaluation of risk of bias (mortality).

| Registration Number | Randomization | Deviations from the intended intervention | Missing outcome data | Measurement of outcome | Selection of the reported results | Final |
|---|---|---|---|---|---|---|
| NCT04384380 | Low | Probably high | Low | Low | Low | High |
| NCT04261517 | Probably low | Probably high | Low | Low | Low | High |
| NCT04325893 | Low | Low | Low | Low | Low | Low |
| IRCT20100228003449N28 | Low | Probably high | Probably high | Low | Low | High |
| NCT04383535 | Low | Low | Low | Low | Low | Low |
| ChiCTR2000029308 | Low | Probably high | Low | Low | Low | High |
| NCT04321278 | Low | Probably high | Probably low | Low | Low | High |
| RBR-8jyhx | Probably low | Low | Low | Low | Low | Low |
| ChiCTR2000029544 | Low | Probably high | Low | Low | Low | High |
| NCT04345523 | Probably low | Probably high | Low | Low | Low | High |
| CTRI202004024775 | Low | Probably high | Low | Low | Probably low | High |
| NCT04326790 | Low | Probably high | Low | Low | Low | High |
| ChiCTR2000029757 | Probably low | Probably high | Low | Low | Low | High |
| NCT04327401 | Low | Probably high | Low | Low | Low | High |
| NCT02517489 | Low | Low | Low | Low | Low | Low |
| NCT02735707 | Low | Probably high | Low | Low | Low | Low |

| Registration Number | Randomization | Deviations from the intended intervention | Missing outcome data | Measurement of outcome | Selection of the reported results | Final |
|---|---|---|---|---|---|---|
| NCT04332991 | Low | Low | Low | Low | Low | Low |
| ChiCTR2000030007 | Low | Probably high | Low | Low | Low | High |
| NCT04292730 | Low | Probably high | Low | Low | Probably low | High |
| NCT04346355 | Low | Probably high | Low | Low | Low | High |
| NCT04331808 | Low | Probably high | Low | Low | Low | High |
| NCT04449718 | Probably low | Low | Low | Low | Low | Low |
| ChiCTR2000030054 | Probably low | Probably high | High | Low | Low | High |
| NCT04252885 | Low | Probably high | Low | Low | Probably low | High |
| NCT04356937 | Low | Low | Low | Low | Probably low | Low |
| IRCT20151227025726N20 | Probably low | Probably high | Low | Low | Low | High |
| ChiCTR2000030254 | Probably low | Probably high | Low | Low | Low | High |
| EudraCT202000193437 | High | Probably high | Low | Low | Low | High |
| NCT04304053 | Low | Probably high | Low | Low | Probably low | High |
| NCT04308668 | Low | Low | Probably low | Low | Low | Low |
| ChiCTR2000029868 | Low | Probably high | Low | Low | Low | High |
| NCT04353336 | Probably low | Probably high | Probably high | Low | Low | High |

| Registration Number | Randomization | Deviations from the intended intervention | Missing outcome data | Measurement of outcome | Selection of the reported results | Final |
|---|---|---|---|---|---|---|
| NCT04329832 | Low | Probably high | Low | Low | Probably low | High |
| NCT04322123 | Low | Probably high | Low | Low | Low | High |
| IRCT20100228003449N27 | Probably low | Probably high | Low | Low | Low | High |
| NCT04411667 | Probably low | Probably high | Low | Low | Probably low | High |
| NCT04343729 | Low | Low | Low | Low | Low | Low |
| NCT04356534 | Probably low | Probably high | Low | Low | Low | High |
| NCT04349592 | Low | Low | Low | Low | Low | Low |
| NCT04381936 | Low | Probably high | Low | Low | Low | High |
| NCT04280705 | Low | Low | Low | Low | Low | Low |
| NCT04257656 | Low | Low | Low | Low | Low | Low |
| NCT04315948 | Low | Probably high | Low | Low | Low | High |
| NCT04385095 | Low | Low | High | Low | Probably low | High |
| IRCT20200501047259N1 | Low | Low | Low | Low | Low | Low |
| NCT04320615 | Probably low | Low | Low | Low | Low | Low |
| NCT04372186 | Probably low | Low | Low | Low | Low | Low |
| NCT04369742 | Probably low | Low | Low | Low | Low | Low |
| NCT04381936_Azi | Low | Probably high | Low | Low | Low | High |

| **Registration Number** | **Randomization** | **Deviations from the intended intervention** | **Missing outcome data** | **Measurement of outcome** | **Selection of the reported results** | **Final** |
|---|---|---|---|---|---|---|
| NCT04401579 | Probably low | Low | Low | Low | Low | Low |
| ISRCTN59048638 | Probably low | Low | Probably high | Low | Low | High |

#### Table S6. Evaluation of risk of bias (mechanical ventilation).

| Registration Number | Randomization | Deviations from the intended intervention | Missing outcome data | Measurement of outcome | Selection of the reported results | Final |
|---|---|---|---|---|---|---|
| NCT04325893 | Low | Low | Low | Low | Low | Low |
| IRCT20100228003449N28 | Low | Probably high | Probably high | Low | Low | High |
| NCT04383535 | Low | Low | Low | Low | Probably low | Low |
| ChiCTR2000029308 | Low | Probably high | Low | Low | Probably low | High |
| NCT04321278 | Low | Probably high | Probably low | Low | Probably low | High |
| NCT04345523 | Probably low | Probably high | Low | Low | Probably low | High |
| CTRI202004024775 | Low | Probably high | Low | Low | Low | High |
| NCT04326790 | Low | Probably high | Low | Low | Low | High |
| NCT04327401 | Low | Probably high | Low | Low | Probably low | High |
| NCT02517489 | Low | Low | Low | Low | Probably low | Low |
| NCT04332991 | Low | Low | Low | Low | Probably low | Low |
| ChiCTR2000030007 | Low | Probably high | Low | Low | Low | High |
| NCT04292730 | Low | Probably high | Low | Low | Probably low | High |
| NCT04449718 | Probably low | Low | Low | Low | Low | Low |
| NCT04356937 | Low | Low | Low | Low | Low | Low |
| IRCT20151227025726N20 | Probably low | Probably high | Low | Low | Low | High |
| EudraCT202000193437 | High | Probably high | Low | Low | Low | High |
| NCT04304053 | Low | Probably high | Probably low | Low | Low | High |
| NCT04353336 | Probably low | Probably high | Probably low | Low | Low | High |
| NCT04329832 | Low | Probably high | Low | Low | Low | High |
| NCT04322123 | Low | Probably high | Low | Low | Low | High |

| Registration Number | Randomization | Deviations from the intended intervention | Missing outcome data | Measurement of outcome | Selection of the reported results | Final |
|---|---|---|---|---|---|---|
| IRCT20100228003449N27 | Probably low | Probably high | Low | Low | Low | High |
| NCT04411667 | Probably low | Probably high | Low | Low | Probably low | High |
| NCT04343729 | Low | Low | Low | Low | Probably low | Low |
| NCT04356534 | Probably low | Probably high | Low | Low | Low | High |
| NCT04381936 | Low | Probably high | Low | Low | Low | High |
| NCT04280705 | Low | Low | Low | Low | Probably low | Low |
| NCT04257656 | Low | Low | Low | Low | Probably low | Low |
| NCT04315948 | Low | Probably high | Low | Low | Low | High |
| NCT04385095 | Low | Low | High | Low | Low | High |
| NCT04320615 | Probably low | Low | Low | Low | Low | Low |
| NCT04369742 | Probably low | Low | Low | Low | Low | Low |
| NCT04381936_Azi | Low | Probably high | Low | Low | Low | High |
| NCT04401579 | Probably low | Low | Probably low | Low | Low | Low |
| ISRCTN59048638 | Probably low | Low | Probably high | Low | Low | High |

#### Table S7. Evaluation of risk of bias (discharge).

| Registration Number | Randomization | Deviations from the intended intervention | Missing outcome data | Measurement of outcome | Selection of the reported results | Final |
|---|---|---|---|---|---|---|
| NCT04434248 | Probably low | Probably high | Low | Low | Probably Low | High |
| NCT04321278 | Low | Probably high | Probably low | Low | Probably Low | High |
| NCT04327401 | Low | Probably high | Low | Low | Probably Low | High |
| NCT04332991 | Low | Low | Low | Low | Low | Low |
| NCT04292730 | Low | Probably high | Low | Low | Probably Low | High |
| NCT04346355 | Low | Probably high | Low | Low | Probably Low | High |
| NCT04331808 | Low | Probably high | Low | Low | Low | High |
| ChiCTR2000030054 | Probably low | Probably high | High | Low | Low | High |
| NCT04356937 | Low | Low | Low | Low | Probably Low | Low |
| NCT04329832 | Low | Probably high | Low | Low | Low | High |
| NCT04322123 | Low | Probably high | Low | Low | Low | High |
| NCT04381936 | Low | Probably high | Low | Low | Low | High |
| NCT04280705 | Low | Low | Low | Low | Low | Low |
| NCT04257656 | Low | Low | Low | Low | Probably Low | Low |
| ChiCTR2000029757 | Probably low | Probably high | Low | Low | Low | High |
| NCT04383535 | Low | Low | Low | Low | Low | Low |
| IRCT20100228003449N27 | Probably low | Probably high | Low | Low | Probably Low | High |
| NCT04320615 | Probably low | Low | Low | Low | Probably Low | Low |
| NCT04325893 | Low | Low | Low | Low | Probably Low | Low |
| NCT04345523 | Probably low | Probably high | Low | Low | Probably Low | High |
| NCT04369742 | Probably low | Low | Low | Low | Probably Low | Low |
| NCT04385095 | Low | Low | High | Low | Probably Low | High |
| ChiCTR2000029308 | Low | Probably high | Low | Low | Low | High |

| Registration Number | Randomization | Deviations from the intended intervention | Missing outcome data | Measurement of outcome | Selection of the reported results | Final |
|---|---|---|---|---|---|---|
| IRCT20100228003449N28 | Low | Probably high | Probably high | Low | Low | High |
| ChiCTR2000029544 | Low | Probably high | Low | Low | Low | High |
| NCT04381936_Azi | Low | Probably high | Low | Low | Low | High |
| NCT04401579 | Probably low | Low | Probably low | Low | Probably Low | Low |

#### Table S8. Evaluation of risk of bias (viral clearance).

| Registration Number | Randomization | Deviations from the intended intervention | Missing outcome data | Measurement of outcome | Selection of the reported results | Final |
|---|---|---|---|---|---|---|
| NCT04384380 | Low | Probably high | Low | Low | Low | High |
| NCT04261517 | Probably low | Probably high | Low | Low | Low | High |
| NCT04325893 | Low | Low | Low | Low | Low | Low |
| NCT04434248 | Probably low | Probably high | Low | Low | Low | High |
| NCT0449199 | Low | Probably high | Low | Low | Low | High |
| ChiCTR2000029544 | Low | Probably high | Low | Low | Low | High |
| NCT04345523 | Probably low | Probably high | Low | Low | Low | High |
| CTRI202004024775 | Low | Probably high | Low | Low | Low | High |
| NCT04552483 | Low | Low | Probably low | Low | Low | Low |
| ChiCTR2000029757 | Probably low | Probably high | Low | Low | Low | High |
| ChiCTR2000029868 | Low | Probably high | Low | Low | Low | High |
| NCT04343729 | Low | Low | Low | Low | Probably Low | Low |
| NCT04349592 | Low | Low | Low | Low | Low | Low |
| NCT04257656 | Low | Low | Low | Low | Low | Low |
| NCT04369742 | Probably low | Low | Low | Low | Low | Low |

#### Table S9. Network meta-analysis results of the primary analysis (log odds ratio, log OR) and evaluation of certainty of evidence (mortality).

| | | Direct | | | | | Indirect | | | | | Network | | | | |
|---|---|---|---|---|---|---|---|---|---|---|---|---|---|---|---|---|
| **Treatment 1** | **Treatment 2** | **EST*** | **LCrI*** | **UCrI*** | **Evidence** | **Reason** | **EST** | **LCrI** | **UCrI** | **Evidence** | **Reason** | **EST** | **LCrI** | **UCrI** | **Evidence** | **Reason** |
| azithromycin | soc | 0.00 | -0.12 | 0.11 | Moderate | RoB | -0.33 | -2.17 | 0.32 | Moderate | NA | 0.00 | -0.12 | 0.11 | Low | Imprecision |
| colchicine | soc | -1.48 | -4.68 | 0.73 | Moderate | RoB | NA | NA | NA | NA | NA | -1.48 | -4.68 | 0.73 | Very low | Severe imprecision |
| hydroxychloroquine | soc | 0.04 | -0.07 | 0.15 | Moderate | RoB | 0.33 | -0.29 | 2.00 | Moderate | NA | 0.03 | -0.08 | 0.14 | Low | Imprecision |
| hydroxychloroquine + azithromycin | soc | -0.65 | -2.22 | 0.59 | Moderate | RoB | NA | NA | NA | NA | NA | -0.65 | -2.22 | 0.59 | Very low | Severe imprecision |
| arbidol | soc | -0.12 | -1.04 | 0.72 | Moderate | RoB | -0.11 | -1.28 | 0.99 | Moderate | NA | -0.11 | -0.99 | 0.77 | Very low | Severe imprecision |
| favipiravir | soc | -0.11 | -1.06 | 0.74 | Moderate | RoB | -0.12 | -1.36 | 1.01 | Moderate | NA | -0.12 | -1.03 | 0.72 | Very low | Severe imprecision |
| remdesivir | soc | -0.08 | -0.21 | 0.04 | Low | RoB, Publication bias | NA | NA | NA | NA | NA | -0.08 | -0.21 | 0.04 | Very low | Imprecision |
| lopinavir/ritonavir | soc | -0.13 | -0.24 | -0.03 | Moderate | RoB | NA | NA | NA | NA | NA | -0.13 | -0.24 | -0.03 | Low | Imprecision |
| convalescent plasma | soc | -0.14 | -0.52 | 0.24 | Moderate | RoB | NA | NA | NA | NA | NA | -0.14 | -0.52 | 0.24 | Low | Imprecision |
| methylprednisolone | soc | -0.09 | -0.38 | 0.26 | Moderate | RoB | NA | NA | NA | NA | NA | -0.09 | -0.38 | 0.26 | Low | Imprecision |
| dexamethasone | soc | -0.16 | -0.28 | -0.05 | Moderate | RoB | NA | NA | NA | NA | NA | -0.16 | -0.28 | -0.05 | Moderate | NA |
| hydrocortisone | soc | -0.26 | -0.64 | 0.04 | High | NA | NA | NA | NA | NA | NA | -0.26 | -0.64 | 0.04 | Moderate | Imprecision |
| intravenous immunoglobulin | soc | -0.52 | -1.19 | 0.14 | Moderate | RoB | NA | NA | NA | NA | NA | -0.52 | -1.19 | 0.14 | Low | Imprecision |
| interferon beta | soc | -0.02 | -0.18 | 0.13 | Low | RoB, Inconsistency | NA | NA | NA | NA | NA | -0.02 | -0.18 | 0.13 | Very low | Imprecision |
| recombinant human gcsf | soc | -0.33 | -1.65 | 0.15 | Moderate | RoB | NA | NA | NA | NA | NA | -0.33 | -1.65 | 0.15 | Very low | Severe imprecision |
| tocilizumab | soc | 0.11 | -0.25 | 0.48 | High | NA | NA | NA | NA | NA | NA | 0.11 | -0.25 | 0.48 | Moderate | Imprecision |
| vitamin d3 | soc | 0.31 | -0.79 | 1.47 | Moderate | RoB | NA | NA | NA | NA | NA | 0.31 | -0.79 | 1.47 | Low | Imprecision |
| baricitinib + remdesivir | soc | NA | NA | NA | NA | NA | -0.54 | -1.10 | 0.00 | Low | NA | -0.54 | -1.10 | 0.00 | Very low | Imprecision |
| sulodexide | soc | -1.00 | -2.61 | 0.36 | Moderate | RoB | NA | NA | NA | NA | NA | -1.00 | -2.61 | 0.36 | Very low | Severe imprecision |
| colchicine | azithromycin | NA | NA | NA | NA | NA | -1.48 | -4.68 | 0.73 | Low | Intransitivity | -1.48 | -4.68 | 0.73 | Very low | Severe imprecision |

| | | Direct | | | | | Indirect | | | | | Network | | | | |
|---|---|---|---|---|---|---|---|---|---|---|---|---|---|---|---|---|
| Treatment 1 | Treatment 2 | EST[*] | LCrI[*] | UCrI[*] | Evidence | Reason | EST | LCrI | UCrI | Evidence | Reason | EST | LCrI | UCrI | Evidence | Reason |
| hydroxychloroquine | azithromycin | 0.37 | -0.30 | 2.23 | Moderate | RoB | 0.03 | -0.13 | 0.19 | Low | Intransitivity | 0.03 | -0.12 | 0.19 | Low | Imprecision |
| hydroxychloroquine + azithromycin | azithromycin | NA | NA | NA | NA | NA | -0.65 | -2.22 | 0.60 | Low | Intransitivity | -0.65 | -2.22 | 0.60 | Very low | Severe imprecision |
| arbidol | azithromycin | NA | NA | NA | NA | NA | -0.11 | -1.00 | 0.78 | Low | Intransitivity | -0.11 | -1.00 | 0.78 | Very low | Severe imprecision |
| favipiravir | azithromycin | NA | NA | NA | NA | NA | -0.11 | -1.03 | 0.73 | Moderate | NA | -0.11 | -1.03 | 0.73 | Very low | Severe imprecision |
| remdesivir | azithromycin | NA | NA | NA | NA | NA | -0.08 | -0.25 | 0.09 | Low | NA | -0.08 | -0.25 | 0.09 | Very low | Imprecision |
| lopinavir/ritonavir | azithromycin | NA | NA | NA | NA | NA | -0.13 | -0.28 | 0.03 | Moderate | NA | -0.13 | -0.28 | 0.03 | Low | Imprecision |
| convalescent plasma | azithromycin | NA | NA | NA | NA | NA | -0.14 | -0.53 | 0.26 | Moderate | NA | -0.14 | -0.53 | 0.26 | Low | Imprecision |
| methylprednisolone | azithromycin | NA | NA | NA | NA | NA | -0.09 | -0.40 | 0.28 | Moderate | NA | -0.09 | -0.40 | 0.28 | Low | Imprecision |
| dexamethasone | azithromycin | NA | NA | NA | NA | NA | -0.16 | -0.32 | 0.00 | Moderate | NA | -0.16 | -0.32 | 0.00 | Low | Imprecision |
| hydrocortisone | azithromycin | NA | NA | NA | NA | NA | -0.26 | -0.66 | 0.07 | Moderate | NA | -0.26 | -0.66 | 0.07 | Low | Imprecision |
| intravenous immunoglobulin | azithromycin | NA | NA | NA | NA | NA | -0.52 | -1.20 | 0.15 | Moderate | NA | -0.52 | -1.20 | 0.15 | Low | Imprecision |
| interferon beta | azithromycin | NA | NA | NA | NA | NA | -0.02 | -0.21 | 0.17 | Low | NA | -0.02 | -0.21 | 0.17 | Very low | Severe imprecision |
| recombinant human gcsf | azithromycin | NA | NA | NA | NA | NA | -0.33 | -1.65 | 0.17 | Low | Intransitivity | -0.33 | -1.65 | 0.17 | Very low | Severe imprecision |
| tocilizumab | azithromycin | NA | NA | NA | NA | NA | 0.11 | -0.27 | 0.50 | Low | Intransitivity | 0.11 | -0.27 | 0.50 | Very low | Imprecision |
| vitamin d3 | azithromycin | NA | NA | NA | NA | NA | 0.31 | -0.79 | 1.48 | Moderate | NA | 0.31 | -0.79 | 1.48 | Very low | Severe imprecision |
| baricitinib + remdesivir | azithromycin | NA | NA | NA | NA | NA | -0.54 | -1.11 | 0.01 | Low | | -0.54 | -1.11 | 0.01 | Very low | Imprecision |
| sulodexide | azithromycin | NA | NA | NA | NA | NA | -0.99 | -2.61 | 0.37 | Low | Intransitivity | -0.99 | -2.61 | 0.37 | Very low | Severe imprecision |
| hydroxychloroquine | colchicine | NA | NA | NA | NA | NA | 1.51 | -0.70 | 4.71 | Moderate | NA | 1.51 | -0.70 | 4.71 | Very low | Severe imprecision |
| hydroxychloroquine + azithromycin | colchicine | NA | NA | NA | NA | NA | 0.82 | -1.88 | 4.23 | Moderate | NA | 0.82 | -1.88 | 4.23 | Very low | Severe imprecision |
| arbidol | colchicine | NA | NA | NA | NA | NA | 1.38 | -1.00 | 4.68 | Moderate | NA | 1.38 | -1.00 | 4.68 | Very low | Severe imprecision |
| favipiravir | colchicine | NA | NA | NA | NA | NA | 1.37 | -1.02 | 4.64 | Moderate | NA | 1.37 | -1.02 | 4.64 | Very low | Severe imprecision |

| | | Direct | | | | | Indirect | | | | | Network | | | | |
|---|---|---|---|---|---|---|---|---|---|---|---|---|---|---|---|---|
| Treatment 1 | Treatment 2 | EST* | LCrI* | UCrI* | Evidence | Reason | EST | LCrI | UCrI | Evidence | Reason | EST | LCrI | UCrI | Evidence | Reason |
| remdesivir | colchicine | NA | NA | NA | NA | NA | 1.40 | -0.81 | 4.60 | Very low | Intransitivity | 1.40 | -0.81 | 4.60 | Very low | Severe imprecision |
| lopinavir/ritonavir | colchicine | NA | NA | NA | NA | NA | 1.35 | -0.86 | 4.55 | Moderate | NA | 1.35 | -0.86 | 4.55 | Very low | Severe imprecision |
| convalescent plasma | colchicine | NA | NA | NA | NA | NA | 1.35 | -0.90 | 4.56 | Low | Intransitivity | 1.35 | -0.90 | 4.56 | Very low | Severe imprecision |
| methylprednisolone | colchicine | NA | NA | NA | NA | NA | 1.40 | -0.83 | 4.60 | Moderate | NA | 1.40 | -0.83 | 4.60 | Very low | Severe imprecision |
| dexamethasone | colchicine | NA | NA | NA | NA | NA | 1.32 | -0.89 | 4.52 | Low | Intransitivity | 1.32 | -0.89 | 4.52 | Very low | Severe imprecision |
| hydrocortisone | colchicine | NA | NA | NA | NA | NA | 1.22 | -1.03 | 4.43 | Low | Intransitivity | 1.22 | -1.03 | 4.43 | Very low | Severe imprecision |
| intravenous immunoglobulin | colchicine | NA | NA | NA | NA | NA | 0.97 | -1.35 | 4.22 | Low | Intransitivity | 0.97 | -1.35 | 4.22 | Very low | Severe imprecision |
| interferon beta | colchicine | NA | NA | NA | NA | NA | 1.46 | -0.75 | 4.66 | Very low | Intransitivity | 1.46 | -0.75 | 4.66 | Very low | Imprecision |
| recombinant human gcsf | colchicine | NA | NA | NA | NA | NA | 1.06 | -1.42 | 4.35 | Moderate | NA | 1.06 | -1.42 | 4.35 | Very low | Severe imprecision |
| tocilizumab | colchicine | NA | NA | NA | NA | NA | 1.60 | -0.65 | 4.81 | Moderate | NA | 1.60 | -0.65 | 4.81 | Very low | Severe imprecision |
| vitamin d3 | colchicine | NA | NA | NA | NA | NA | 1.81 | -0.68 | 5.17 | Low | Intransitivity | 1.81 | -0.68 | 5.17 | Very low | Severe imprecision |
| baricitinib + remdesivir | colchicine | NA | NA | NA | NA | NA | 0.94 | -1.33 | 4.18 | Low | | 0.94 | -1.33 | 4.18 | Very low | Severe imprecision |
| sulodexide | colchicine | NA | NA | NA | NA | NA | 0.49 | -2.24 | 3.92 | Moderate | | 0.49 | -2.24 | 3.92 | Very low | Severe imprecision |
| hydroxychloroquine + azithromycin | hydroxychloroquine | -0.68 | -2.26 | 0.56 | Moderate | RoB | NA | NA | NA | Moderate | NA | -0.68 | -2.26 | 0.56 | Very low | Severe imprecision |
| arbidol | hydroxychloroquine | NA | NA | NA | NA | NA | -0.14 | -1.03 | 0.74 | Moderate | NA | -0.14 | -1.03 | 0.74 | Very low | Severe imprecision |
| favipiravir | hydroxychloroquine | NA | NA | NA | NA | NA | -0.15 | -1.06 | 0.69 | Moderate | NA | -0.15 | -1.06 | 0.69 | Very low | Severe imprecision |
| remdesivir | hydroxychloroquine | -0.04 | -0.25 | 0.19 | NA | NA | -0.26 | -0.58 | 0.01 | Low | NA | -0.12 | -0.27 | 0.04 | Very low | Imprecision |
| lopinavir/ritonavir | hydroxychloroquine | -0.16 | -0.29 | -0.02 | Moderate | RoB | -0.20 | -0.78 | 0.27 | Moderate | NA | -0.16 | -0.30 | -0.03 | Moderate | NA |
| convalescent plasma | hydroxychloroquine | NA | NA | NA | NA | NA | -0.17 | -0.57 | 0.23 | Low | Intransitivity | -0.17 | -0.57 | 0.23 | Very low | Imprecision |

| | | Direct | | | | | Indirect | | | | | Network | | | | |
|---|---|---|---|---|---|---|---|---|---|---|---|---|---|---|---|---|
| **Treatment 1** | **Treatment 2** | **EST*** | **LCrI*** | **UCrI*** | **Evidence** | **Reason** | **EST** | **LCrI** | **UCrI** | **Evidence** | **Reason** | **EST** | **LCrI** | **UCrI** | **Evidence** | **Reason** |
| methylprednisolone | hydroxychloroquine | NA | NA | NA | NA | NA | -0.12 | -0.43 | 0.24 | Moderate | NA | -0.12 | -0.43 | 0.24 | Low | Imprecision |
| dexamethasone | hydroxychloroquine | -0.21 | -0.36 | -0.07 | Moderate | RoB | -0.18 | -0.54 | 0.19 | Low | Intransitivity | -0.19 | -0.33 | -0.06 | Moderate | NA |
| hydrocortisone | hydroxychloroquine | NA | NA | NA | NA | NA | -0.29 | -0.69 | 0.03 | Low | Intransitivity | -0.29 | -0.69 | 0.03 | Very low | Imprecision |
| intravenous immunoglobulin | hydroxychloroquine | NA | NA | NA | NA | NA | -0.55 | -1.23 | 0.12 | Low | Intransitivity | -0.55 | -1.23 | 0.12 | Very low | Imprecision |
| interferon beta | hydroxychloroquine | 0.03 | -0.21 | 0.28 | Moderate | RoB | -1.07 | -1.93 | -0.31 | Very low | Intransitivity | -0.05 | -0.23 | 0.12 | Very low | Imprecision |
| recombinant human gcsf | hydroxychloroquine | NA | NA | NA | NA | NA | -0.36 | -1.68 | 0.13 | Moderate | NA | -0.36 | -1.68 | 0.13 | Very low | Severe imprecision |
| tocilizumab | hydroxychloroquine | NA | NA | NA | NA | NA | 0.08 | -0.30 | 0.46 | Moderate | NA | 0.08 | -0.30 | 0.46 | Low | Imprecision |
| vitamin d3 | hydroxychloroquine | NA | NA | NA | NA | NA | 0.28 | -0.83 | 1.45 | Low | Intransitivity | 0.28 | -0.83 | 1.45 | Very low | Imprecision |
| baricitinib + remdesivir | hydroxychloroquine | NA | NA | NA | NA | NA | -0.57 | -1.14 | -0.03 | Low | NA | -0.57 | -1.14 | -0.03 | Low | NA |
| sulodexide | hydroxychloroquine | NA | NA | NA | NA | NA | -1.03 | -2.65 | 0.33 | Moderate | NA | -1.03 | -2.65 | 0.33 | Very low | Severe imprecision |
| arbidol | hydroxychloroquine + azithromycin | NA | NA | NA | NA | NA | 0.55 | -0.94 | 2.33 | Moderate | NA | 0.55 | -0.94 | 2.33 | Very low | Severe imprecision |
| favipiravir | hydroxychloroquine + azithromycin | NA | NA | NA | NA | NA | 0.54 | -0.97 | 2.31 | Moderate | NA | 0.54 | -0.97 | 2.31 | Very low | Severe imprecision |
| remdesivir | hydroxychloroquine + azithromycin | NA | NA | NA | NA | NA | 0.57 | -0.68 | 2.15 | Very low | Intransitivity | 0.57 | -0.68 | 2.15 | Very low | Severe imprecision |
| lopinavir/ritonavir | hydroxychloroquine + azithromycin | NA | NA | NA | NA | NA | 0.52 | -0.72 | 2.10 | Moderate | NA | 0.52 | -0.72 | 2.10 | Very low | Severe imprecision |
| convalescent plasma | hydroxychloroquine + azithromycin | NA | NA | NA | NA | NA | 0.52 | -0.78 | 2.13 | Low | Intransitivity | 0.52 | -0.78 | 2.13 | Very low | Severe imprecision |
| methylprednisolone | hydroxychloroquine + azithromycin | NA | NA | NA | NA | NA | 0.57 | -0.72 | 2.17 | Moderate | NA | 0.57 | -0.72 | 2.17 | Very low | Severe imprecision |
| dexamethasone | hydroxychloroquine + azithromycin | NA | NA | NA | NA | NA | 0.49 | -0.75 | 2.07 | Low | Intransitivity | 0.49 | -0.75 | 2.07 | Very low | Severe imprecision |
| hydrocortisone | hydroxychloroquine + azithromycin | NA | NA | NA | NA | NA | 0.38 | -0.91 | 1.99 | Low | Intransitivity | 0.38 | -0.91 | 1.99 | Very low | Severe imprecision |
| intravenous immunoglobulin | hydroxychloroquine + azithromycin | NA | NA | NA | NA | NA | 0.14 | -1.28 | 1.83 | Low | Intransitivity | 0.14 | -1.28 | 1.83 | Very low | Severe imprecision |
| interferon beta | hydroxychloroquine + azithromycin | NA | NA | NA | NA | NA | 0.63 | -0.62 | 2.21 | Very low | Intransitivity | 0.63 | -0.62 | 2.21 | Very low | Severe imprecision |

| | | Direct | | | | | Indirect | | | | | Network | | | | |
|---|---|---|---|---|---|---|---|---|---|---|---|---|---|---|---|---|
| **Treatment 1** | **Treatment 2** | **EST*** | **LCrI*** | **UCrI*** | **Evidence** | **Reason** | **EST** | **LCrI** | **UCrI** | **Evidence** | **Reason** | **EST** | **LCrI** | **UCrI** | **Evidence** | **Reason** |
| recombinant human gcsf | hydroxychloroquine + azithromycin | NA | NA | NA | NA | NA | 0.24 | -1.44 | 1.94 | Moderate | NA | 0.24 | -1.44 | 1.94 | Very low | Severe imprecision |
| tocilizumab | hydroxychloroquine + azithromycin | NA | NA | NA | NA | NA | 0.76 | -0.53 | 2.37 | Moderate | NA | 0.76 | -0.53 | 2.37 | Very low | Severe imprecision |
| vitamin d3 | hydroxychloroquine + azithromycin | NA | NA | NA | NA | NA | 0.98 | -0.70 | 2.90 | Low | Intransitivity | 0.98 | -0.70 | 2.90 | Very low | Severe imprecision |
| baricitinib + remdesivir | hydroxychloroquine + azithromycin | NA | NA | NA | NA | NA | 0.11 | -1.25 | 1.77 | Low | NA | 0.11 | -1.25 | 1.77 | Very low | Severe imprecision |
| sulodexide | hydroxychloroquine + azithromycin | NA | NA | NA | NA | NA | -0.34 | -2.36 | 1.70 | Moderate | NA | -0.34 | -2.36 | 1.70 | Very low | Severe imprecision |
| favipiravir | arbidol | 0.00 | -0.71 | 0.71 | Moderate | RoB | 0.00 | -1.00 | 1.01 | Moderate | NA | 0.00 | -1.05 | 0.98 | Low | Imprecision |
| remdesivir | arbidol | NA | NA | NA | NA | NA | 0.02 | -0.85 | 0.91 | Very low | Intransitivity | 0.02 | -0.85 | 0.91 | Very low | Severe imprecision |
| lopinavir/ritonavir | arbidol | 0.00 | -0.75 | 0.68 | Moderate | RoB | -0.02 | -1.19 | 1.09 | Moderate | NA | -0.02 | -0.91 | 0.86 | Very low | Severe imprecision |
| convalescent plasma | arbidol | NA | NA | NA | NA | NA | -0.03 | -0.97 | 0.91 | Low | Intransitivity | -0.03 | -0.97 | 0.91 | Very low | Severe imprecision |
| methylprednisolone | arbidol | NA | NA | NA | NA | NA | 0.01 | -0.85 | 1.00 | Moderate | NA | 0.01 | -0.85 | 1.00 | Very low | Severe imprecision |
| dexamethasone | arbidol | NA | NA | NA | NA | NA | -0.05 | -0.94 | 0.84 | Low | Intransitivity | -0.05 | -0.94 | 0.84 | Very low | Severe imprecision |
| hydrocortisone | arbidol | NA | NA | NA | NA | NA | -0.13 | -1.17 | 0.71 | Low | Intransitivity | -0.13 | -1.17 | 0.71 | Very low | Severe imprecision |
| intravenous immunoglobulin | arbidol | NA | NA | NA | NA | NA | -0.41 | -1.48 | 0.65 | Low | Intransitivity | -0.41 | -1.48 | 0.65 | Very low | Severe imprecision |
| interferon beta | arbidol | NA | NA | NA | NA | NA | 0.09 | -0.79 | 0.99 | Very low | Intransitivity | 0.09 | -0.79 | 0.99 | Very low | Severe imprecision |
| recombinant human gcsf | arbidol | NA | NA | NA | NA | NA | -0.19 | -2.00 | 0.49 | Moderate | NA | -0.19 | -2.00 | 0.49 | Very low | Severe imprecision |
| tocilizumab | arbidol | NA | NA | NA | NA | NA | 0.22 | -0.72 | 1.16 | Moderate | NA | 0.22 | -0.72 | 1.16 | Very low | Severe imprecision |
| vitamin d3 | arbidol | NA | NA | NA | NA | NA | 0.42 | -0.96 | 1.86 | Low | Intransitivity | 0.42 | -0.96 | 1.86 | Very low | Severe imprecision |

| | | Direct | | | | | Indirect | | | | | Network | | | | |
|---|---|---|---|---|---|---|---|---|---|---|---|---|---|---|---|---|
| Treatment 1 | Treatment 2 | EST* | LCrI* | UCrI* | Evidence | Reason | EST | LCrI | UCrI | Evidence | Reason | EST | LCrI | UCrI | Evidence | Reason |
| baricitinib + remdesivir | arbidol | NA | NA | NA | NA | NA | -0.43 | -1.43 | 0.57 | Low | NA | -0.43 | -1.43 | 0.57 | Very low | Severe imprecision |
| sulodexide | arbidol | NA | NA | NA | NA | NA | -0.89 | -2.71 | 0.71 | Moderate | NA | -0.89 | -2.71 | 0.71 | Very low | Severe imprecision |
| remdesivir | favipiravir | NA | NA | NA | NA | NA | 0.03 | -0.79 | 0.95 | Low | NA | 0.03 | -0.79 | 0.95 | Very low | Severe imprecision |
| lopinavir/ritonavir | favipiravir | NA | NA | NA | NA | NA | -0.01 | -0.85 | 0.89 | Moderate | NA | -0.01 | -0.85 | 0.89 | Very low | Severe imprecision |
| convalescent plasma | favipiravir | NA | NA | NA | NA | NA | -0.02 | -0.92 | 0.94 | Moderate | NA | -0.02 | -0.92 | 0.94 | Very low | Severe imprecision |
| methylprednisolone | favipiravir | NA | NA | NA | NA | NA | 0.01 | -0.81 | 1.04 | Moderate | NA | 0.01 | -0.81 | 1.04 | Very low | Severe imprecision |
| dexamethasone | favipiravir | NA | NA | NA | NA | NA | -0.05 | -0.88 | 0.87 | Moderate | NA | -0.05 | -0.88 | 0.87 | Very low | Severe imprecision |
| hydrocortisone | favipiravir | NA | NA | NA | NA | NA | -0.13 | -1.12 | 0.74 | Moderate | NA | -0.13 | -1.12 | 0.74 | Very low | Severe imprecision |
| intravenous immunoglobulin | favipiravir | NA | NA | NA | NA | NA | -0.40 | -1.44 | 0.68 | Moderate | NA | -0.40 | -1.44 | 0.68 | Very low | Severe imprecision |
| interferon beta | favipiravir | NA | NA | NA | NA | NA | 0.09 | -0.74 | 1.02 | Low | NA | 0.09 | -0.74 | 1.02 | Very low | Severe imprecision |
| recombinant human gcsf | favipiravir | NA | NA | NA | NA | NA | -0.18 | -1.91 | 0.50 | Moderate | NA | -0.18 | -1.91 | 0.50 | Very low | Severe imprecision |
| tocilizumab | favipiravir | NA | NA | NA | NA | NA | 0.23 | -0.66 | 1.19 | Moderate | NA | 0.23 | -0.66 | 1.19 | Very low | Severe imprecision |
| vitamin d3 | favipiravir | NA | NA | NA | NA | NA | 0.43 | -0.93 | 1.88 | Moderate | NA | 0.43 | -0.93 | 1.88 | Very low | Severe imprecision |
| baricitinib + remdesivir | favipiravir | NA | NA | NA | NA | NA | -0.43 | -1.38 | 0.60 | Low | NA | -0.43 | -1.38 | 0.60 | Very low | Imprecision |
| sulodexide | favipiravir | NA | NA | NA | NA | NA | -0.88 | -2.68 | 0.72 | Moderate | NA | -0.88 | -2.68 | 0.72 | Very low | Severe imprecision |
| lopinavir/ritonavir | remdesivir | -0.03 | -0.21 | 0.14 | Moderate | RoB | 0.08 | -0.19 | 0.38 | Low | NA | -0.05 | -0.20 | 0.10 | Very low | Imprecision |
| convalescent plasma | remdesivir | NA | NA | NA | NA | NA | -0.05 | -0.46 | 0.35 | Low | NA | -0.05 | -0.46 | 0.35 | Very low | Imprecision |
| methylprednisolone | remdesivir | NA | NA | NA | NA | NA | -0.01 | -0.33 | 0.36 | Low | NA | -0.01 | -0.33 | 0.36 | Very low | Imprecision |
| dexamethasone | remdesivir | NA | NA | NA | NA | NA | -0.08 | -0.24 | 0.09 | Low | NA | -0.08 | -0.24 | 0.09 | Very low | Imprecision |

| | | Direct | | | | | Indirect | | | | | Network | | | | |
|---|---|---|---|---|---|---|---|---|---|---|---|---|---|---|---|---|
| **Treatment 1** | **Treatment 2** | **EST*** | **LCrI*** | **UCrI*** | **Evidence** | **Reason** | **EST** | **LCrI** | **UCrI** | **Evidence** | **Reason** | **EST** | **LCrI** | **UCrI** | **Evidence** | **Reason** |
| hydrocortisone | remdesivir | NA | NA | NA | NA | NA | -0.18 | -0.58 | 0.15 | Low | NA | -0.18 | -0.58 | 0.15 | Very low | Imprecision |
| intravenous immunoglobulin | remdesivir | NA | NA | NA | NA | NA | -0.44 | -1.12 | 0.23 | Low | NA | -0.44 | -1.12 | 0.23 | Very low | Imprecision |
| interferon beta | remdesivir | 0.05 | -0.13 | 0.23 | Moderate | RoB | -0.66 | -1.56 | 0.05 | Low | NA | 0.06 | -0.11 | 0.24 | Very low | Severe imprecision |
| recombinant human gcsf | remdesivir | NA | NA | NA | NA | NA | -0.24 | -1.57 | 0.25 | Very low | Intransitivity | -0.24 | -1.57 | 0.25 | Very low | Severe imprecision |
| tocilizumab | remdesivir | NA | NA | NA | NA | NA | 0.19 | -0.19 | 0.59 | Very low | Intransitivity | 0.19 | -0.19 | 0.59 | Very low | Imprecision |
| vitamin d3 | remdesivir | NA | NA | NA | NA | NA | 0.39 | -0.72 | 1.57 | Low | NA | 0.39 | -0.72 | 1.57 | Very low | Severe imprecision |
| baricitinib + remdesivir | remdesivir | -0.46 | -1.00 | 0.06 | High | NA | NA | NA | NA | NA | NA | -0.46 | -1.00 | 0.06 | Moderate | Imprecision |
| sulodexide | remdesivir | NA | NA | NA | NA | NA | -0.91 | -2.54 | 0.45 | Low | NA | -0.91 | -2.54 | 0.45 | Very low | Severe imprecision |
| convalescent plasma | lopinavir/ritonavir | NA | NA | NA | NA | NA | -0.01 | -0.40 | 0.39 | Moderate | NA | -0.01 | -0.40 | 0.39 | Low | Imprecision |
| methylprednisolone | lopinavir/ritonavir | NA | NA | NA | NA | NA | 0.04 | -0.27 | 0.40 | Moderate | NA | 0.04 | -0.27 | 0.40 | Low | Imprecision |
| dexamethasone | lopinavir/ritonavir | -0.02 | -0.17 | 0.13 | Moderate | RoB | -0.07 | -0.45 | 0.28 | Moderate | NA | -0.03 | -0.17 | 0.11 | Low | Imprecision |
| hydrocortisone | lopinavir/ritonavir | NA | NA | NA | NA | NA | -0.13 | -0.52 | 0.19 | Moderate | NA | -0.13 | -0.52 | 0.19 | Low | Imprecision |
| intravenous immunoglobulin | lopinavir/ritonavir | NA | NA | NA | NA | NA | -0.39 | -1.07 | 0.28 | Moderate | NA | -0.39 | -1.07 | 0.28 | Low | Imprecision |
| interferon beta | lopinavir/ritonavir | 0.08 | -0.14 | 0.30 | Moderate | RoB | -0.82 | -1.69 | -0.07 | Moderate | NA | 0.11 | -0.06 | 0.28 | Very low | Severe imprecision |
| recombinant human gcsf | lopinavir/ritonavir | NA | NA | NA | NA | NA | -0.20 | -1.51 | 0.29 | Moderate | NA | -0.20 | -1.51 | 0.29 | Very low | Severe imprecision |
| tocilizumab | lopinavir/ritonavir | NA | NA | NA | NA | NA | 0.24 | -0.14 | 0.63 | Moderate | NA | 0.24 | -0.14 | 0.63 | Low | Imprecision |
| vitamin d3 | lopinavir/ritonavir | NA | NA | NA | NA | NA | 0.44 | -0.67 | 1.61 | Moderate | NA | 0.44 | -0.67 | 1.61 | Very low | Severe imprecision |
| baricitinib + remdesivir | lopinavir/ritonavir | NA | NA | NA | NA | NA | -0.41 | -0.97 | 0.13 | Low | NA | -0.41 | -0.97 | 0.13 | Very low | Imprecision |
| sulodexide | lopinavir/ritonavir | NA | NA | NA | NA | NA | -0.86 | -2.48 | 0.50 | Moderate | NA | -0.86 | -2.48 | 0.50 | Very low | Severe imprecision |
| methylprednisolone | convalescent plasma | NA | NA | NA | NA | NA | 0.05 | -0.43 | 0.56 | Moderate | NA | 0.05 | -0.43 | 0.56 | Low | Imprecision |
| dexamethasone | convalescent plasma | NA | NA | NA | NA | NA | -0.02 | -0.42 | 0.38 | Moderate | NA | -0.02 | -0.42 | 0.38 | Low | Imprecision |
| hydrocortisone | convalescent plasma | NA | NA | NA | NA | NA | -0.13 | -0.65 | 0.37 | Moderate | NA | -0.13 | -0.65 | 0.37 | Low | Imprecision |

| | | Direct | | | | | Indirect | | | | | Network | | | | |
|---|---|---|---|---|---|---|---|---|---|---|---|---|---|---|---|---|
| Treatment 1 | Treatment 2 | EST* | LCrI* | UCrI* | Evidence | Reason | EST | LCrI | UCrI | Evidence | Reason | EST | LCrI | UCrI | Evidence | Reason |
| intravenous immunoglobulin | convalescent plasma | NA | NA | NA | NA | NA | -0.38 | -1.16 | 0.38 | Moderate | NA | -0.38 | -1.16 | 0.38 | Low | Imprecision |
| interferon beta | convalescent plasma | NA | NA | NA | NA | NA | 0.12 | -0.29 | 0.53 | Moderate | NA | 0.12 | -0.29 | 0.53 | Very low | Severe imprecision |
| recombinant human gcsf | convalescent plasma | NA | NA | NA | NA | NA | -0.22 | -1.55 | 0.44 | Low | Intransitivity | -0.22 | -1.55 | 0.44 | Very low | Severe imprecision |
| tocilizumab | convalescent plasma | NA | NA | NA | NA | NA | 0.25 | -0.28 | 0.78 | Low | Intransitivity | 0.25 | -0.28 | 0.78 | Very low | Imprecision |
| vitamin d3 | convalescent plasma | NA | NA | NA | NA | NA | 0.45 | -0.72 | 1.68 | Moderate | NA | 0.45 | -0.72 | 1.68 | Very low | Severe imprecision |
| baricitinib + remdesivir | convalescent plasma | NA | NA | NA | NA | NA | -0.40 | -1.08 | 0.26 | Low | NA | -0.40 | -1.08 | 0.26 | Very low | Imprecision |
| sulodexide | convalescent plasma | NA | NA | NA | NA | NA | -0.86 | -2.51 | 0.55 | Low | Intransitivity | -0.86 | -2.51 | 0.55 | Very low | Severe imprecision |
| dexamethasone | methylprednisolone | NA | NA | NA | NA | NA | -0.07 | -0.43 | 0.23 | Moderate | NA | -0.07 | -0.43 | 0.23 | Low | Imprecision |
| hydrocortisone | methylprednisolone | NA | NA | NA | NA | NA | -0.16 | -0.70 | 0.20 | Moderate | NA | -0.16 | -0.70 | 0.20 | Low | Imprecision |
| intravenous immunoglobulin | methylprednisolone | NA | NA | NA | NA | NA | -0.43 | -1.18 | 0.30 | Moderate | NA | -0.43 | -1.18 | 0.30 | Low | Imprecision |
| interferon beta | methylprednisolone | NA | NA | NA | NA | NA | 0.07 | -0.30 | 0.40 | Moderate | NA | 0.07 | -0.30 | 0.40 | Very low | Severe imprecision |
| recombinant human gcsf | methylprednisolone | NA | NA | NA | NA | NA | -0.24 | -1.67 | 0.34 | Moderate | NA | -0.24 | -1.67 | 0.34 | Very low | Severe imprecision |
| tocilizumab | methylprednisolone | NA | NA | NA | NA | NA | 0.20 | -0.30 | 0.67 | Moderate | NA | 0.20 | -0.30 | 0.67 | Low | Imprecision |
| vitamin d3 | methylprednisolone | NA | NA | NA | NA | NA | 0.39 | -0.75 | 1.60 | Moderate | NA | 0.39 | -0.75 | 1.60 | Very low | Severe imprecision |
| baricitinib + remdesivir | methylprednisolone | NA | NA | NA | NA | NA | -0.45 | -1.10 | 0.16 | Low | NA | -0.45 | -1.10 | 0.16 | Very low | Imprecision |
| sulodexide | methylprednisolone | NA | NA | NA | NA | NA | -0.91 | -2.55 | 0.48 | Moderate | NA | -0.91 | -2.55 | 0.48 | Very low | Severe imprecision |
| hydrocortisone | dexamethasone | NA | NA | NA | NA | NA | -0.10 | -0.49 | 0.21 | Moderate | NA | -0.10 | -0.49 | 0.21 | Low | Imprecision |
| intravenous immunoglobulin | dexamethasone | NA | NA | NA | NA | NA | -0.36 | -1.04 | 0.31 | Moderate | NA | -0.36 | -1.04 | 0.31 | Low | Imprecision |
| interferon beta | dexamethasone | NA | NA | NA | NA | NA | 0.14 | -0.05 | 0.33 | Low | NA | 0.14 | -0.05 | 0.33 | Very low | Severe imprecision |

| | | Direct | | | | | Indirect | | | | | Network | | | | |
|---|---|---|---|---|---|---|---|---|---|---|---|---|---|---|---|---|
| **Treatment 1** | **Treatment 2** | **EST*** | **LCrI*** | **UCrI*** | **Evidence** | **Reason** | **EST** | **LCrI** | **UCrI** | **Evidence** | **Reason** | **EST** | **LCrI** | **UCrI** | **Evidence** | **Reason** |
| recombinant human gcsf | dexamethasone | NA | NA | NA | NA | NA | -0.17 | -1.49 | 0.33 | Low | Intransitivity | -0.17 | -1.49 | 0.33 | Very low | Severe imprecision |
| tocilizumab | dexamethasone | NA | NA | NA | NA | NA | 0.27 | -0.11 | 0.66 | Low | Intransitivity | 0.27 | -0.11 | 0.66 | Very low | Imprecision |
| vitamin d3 | dexamethasone | NA | NA | NA | NA | NA | 0.47 | -0.64 | 1.64 | Moderate | NA | 0.47 | -0.64 | 1.64 | Low | Imprecision |
| baricitinib + remdesivir | dexamethasone | NA | NA | NA | NA | NA | -0.38 | -0.95 | 0.17 | Low | NA | -0.38 | -0.95 | 0.17 | Very low | Imprecision |
| sulodexide | dexamethasone | NA | NA | NA | NA | NA | -0.83 | -2.45 | 0.53 | Low | Intransitivity | -0.83 | -2.45 | 0.53 | Very low | Severe imprecision |
| intravenous immunoglobulin | hydrocortisone | NA | NA | NA | NA | NA | -0.25 | -1.00 | 0.50 | Moderate | NA | -0.25 | -1.00 | 0.50 | Low | Imprecision |
| interferon beta | hydrocortisone | NA | NA | NA | NA | NA | 0.24 | -0.10 | 0.66 | Low | NA | 0.24 | -0.10 | 0.66 | Very low | Severe imprecision |
| recombinant human gcsf | hydrocortisone | NA | NA | NA | NA | NA | -0.08 | -1.34 | 0.48 | Low | Intransitivity | -0.08 | -1.34 | 0.48 | Very low | Severe imprecision |
| tocilizumab | hydrocortisone | NA | NA | NA | NA | NA | 0.38 | -0.11 | 0.90 | Moderate | Intransitivity | 0.38 | -0.11 | 0.90 | Low | Imprecision |
| vitamin d3 | hydrocortisone | NA | NA | NA | NA | NA | 0.58 | -0.57 | 1.80 | Moderate | NA | 0.58 | -0.57 | 1.80 | Very low | Severe imprecision |
| baricitinib + remdesivir | hydrocortisone | NA | NA | NA | NA | NA | -0.27 | -0.92 | 0.38 | Low | NA | -0.27 | -0.92 | 0.38 | Very low | Imprecision |
| sulodexide | hydrocortisone | NA | NA | NA | NA | NA | -0.73 | -2.37 | 0.68 | Low | Intransitivity | -0.73 | -2.37 | 0.68 | Very low | Severe imprecision |
| interferon beta | intravenous immunoglobulin | NA | NA | NA | NA | NA | 0.50 | -0.18 | 1.19 | Low | NA | 0.50 | -0.18 | 1.19 | Low | Imprecision |
| recombinant human gcsf | intravenous immunoglobulin | NA | NA | NA | NA | NA | 0.14 | -1.27 | 1.03 | Low | Intransitivity | 0.14 | -1.27 | 1.03 | Very low | Severe imprecision |
| tocilizumab | intravenous immunoglobulin | NA | NA | NA | NA | NA | 0.63 | -0.13 | 1.39 | Low | Intransitivity | 0.63 | -0.13 | 1.39 | Very low | Imprecision |
| vitamin d3 | intravenous immunoglobulin | NA | NA | NA | NA | NA | 0.83 | -0.45 | 2.17 | Moderate | NA | 0.83 | -0.45 | 2.17 | Very low | Severe imprecision |
| baricitinib + remdesivir | intravenous immunoglobulin | NA | NA | NA | NA | NA | -0.02 | -0.88 | 0.84 | Low | NA | -0.02 | -0.88 | 0.84 | Very low | Imprecision |
| sulodexide | intravenous immunoglobulin | NA | NA | NA | NA | NA | -0.48 | -2.22 | 1.04 | Low | Intransitivity | -0.48 | -2.22 | 1.04 | Very low | Severe imprecision |

| | | Direct | | | | | Indirect | | | | | Network | | | | |
|---|---|---|---|---|---|---|---|---|---|---|---|---|---|---|---|---|
| **Treatment 1** | **Treatment 2** | **EST*** | **LCrI*** | **UCrI*** | **Evidence** | **Reason** | **EST** | **LCrI** | **UCrI** | **Evidence** | **Reason** | **EST** | **LCrI** | **UCrI** | **Evidence** | **Reason** |
| recombinant human gcsf | interferon beta | NA | NA | NA | NA | NA | -0.30 | -1.64 | 0.15 | Very low | Intransitivity | -0.30 | -1.64 | 0.15 | Very low | Imprecision |
| tocilizumab | interferon beta | NA | NA | NA | NA | NA | 0.13 | -0.26 | 0.53 | Very low | Intransitivity | 0.13 | -0.26 | 0.53 | Very low | Imprecision |
| vitamin d3 | interferon beta | NA | NA | NA | NA | NA | 0.33 | -0.78 | 1.51 | Low | NA | 0.33 | -0.78 | 1.51 | Very low | Severe imprecision |
| baricitinib + remdesivir | interferon beta | NA | NA | NA | NA | NA | -0.52 | -1.09 | 0.03 | Low | NA | -0.52 | -1.09 | 0.03 | Very low | Imprecision |
| sulodexide | interferon beta | NA | NA | NA | NA | NA | -0.97 | -2.60 | 0.39 | Very low | Intransitivity | -0.97 | -2.60 | 0.39 | Very low | Severe imprecision |
| tocilizumab | recombinant human gcsf | NA | NA | NA | NA | NA | 0.46 | -0.18 | 1.80 | Moderate | NA | 0.46 | -0.18 | 1.80 | Very low | Severe imprecision |
| vitamin d3 | recombinant human gcsf | NA | NA | NA | NA | NA | 0.72 | -0.57 | 2.36 | Low | Intransitivity | 0.72 | -0.57 | 2.36 | Very low | Severe imprecision |
| baricitinib + remdesivir | recombinant human gcsf | NA | NA | NA | NA | NA | -0.17 | -0.96 | 1.20 | Low | NA | -0.17 | -0.96 | 1.20 | Very low | Imprecision |
| sulodexide | recombinant human gcsf | NA | NA | NA | NA | NA | -0.58 | -2.34 | 1.18 | Moderate | NA | -0.58 | -2.34 | 1.18 | Very low | Severe imprecision |
| vitamin d3 | tocilizumab | NA | NA | NA | NA | NA | 0.20 | -0.96 | 1.42 | Low | Intransitivity | 0.20 | -0.96 | 1.42 | Very low | Imprecision |
| baricitinib + remdesivir | tocilizumab | NA | NA | NA | NA | NA | -0.65 | -1.32 | 0.00 | Low | NA | -0.65 | -1.32 | 0.00 | Very low | Imprecision |
| sulodexide | tocilizumab | NA | NA | NA | NA | NA | -1.11 | -2.76 | 0.29 | Moderate | NA | -1.11 | -2.76 | 0.29 | Very low | Severe imprecision |
| baricitinib + remdesivir | vitamin d3 | NA | NA | NA | NA | NA | -0.85 | -2.14 | 0.37 | Low | NA | -0.85 | -2.14 | 0.37 | Very low | Severe imprecision |
| sulodexide | vitamin d3 | NA | NA | NA | NA | NA | -1.32 | -3.30 | 0.45 | Low | Intransitivity | -1.32 | -3.30 | 0.45 | Very low | Severe imprecision |
| sulodexide | baricitinib + remdesivir | NA | NA | NA | NA | NA | -0.46 | -2.15 | 1.02 | Low | NA | -0.46 | -2.15 | 1.02 | Very low | Severe imprecision |

*EST: estimate; LCrI: lower credible interval; UCrI: upper credible interval.

#### Table S10. Network meta-analysis results of the primary analysis (log odds ratio, log OR) and evaluation of certainty of evidence (mechanical ventilation).

| | | Direct | | | | | Indirect | | | | | Network | | | | |
|---|---|---|---|---|---|---|---|---|---|---|---|---|---|---|---|---|
| **Treatment 1** | **Treatment 2** | **EST*** | **LCrI*** | **UCrI*** | **Evidence** | **Reason** | **EST** | **LCrI** | **UCrI** | **Evidence** | **Reason** | **EST** | **LCrI** | **UCrI** | **Evidence** | **Reason** |
| azithromycin | soc | 0.02 | -0.16 | 0.19 | Very low | RoB, Indirectness, Inconsistency | -0.21 | -1.06 | 0.62 | Moderate | NA | 0.01 | -0.16 | 0.18 | Low | Imprecision |
| hydroxychloroquine | soc | 0.02 | -0.13 | 0.17 | Moderate | RoB | 0.22 | -0.60 | 1.06 | Moderate | NA | -0.01 | -0.15 | 0.14 | Low | Imprecision |
| hydroxychloroquine + azithromycin | soc | 0.46 | -0.19 | 1.09 | Moderate | RoB | NA | NA | NA | NA | NA | 0.46 | -0.19 | 1.09 | Low | Imprecision |
| remdesivir | soc | 0.06 | -0.07 | 0.19 | Very low | Inconsistency, Publication Bias, RoB, Indirectness | NA | NA | NA | NA | NA | 0.06 | -0.07 | 0.19 | Very low | Imprecision |
| lopinavir/ritonavir | soc | 0.02 | -0.12 | 0.16 | Moderate | RoB | NA | NA | NA | NA | NA | 0.02 | -0.12 | 0.16 | Low | Imprecision |
| convalescent plasma | soc | -0.04 | -0.37 | 0.29 | Moderate | RoB | NA | NA | NA | NA | NA | -0.04 | -0.37 | 0.29 | Low | Imprecision |
| methylprednisolone | soc | 0.08 | -0.53 | 0.71 | Moderate | RoB | NA | NA | NA | NA | NA | 0.08 | -0.53 | 0.71 | Low | Imprecision |
| dexamethasone | soc | -0.38 | -0.59 | -0.19 | Low | RoB, Indirectnessess | NA | NA | NA | NA | NA | -0.38 | -0.59 | -0.19 | Low | NA |
| hydrocortisone | soc | -0.08 | -0.76 | 0.62 | Moderate | Indirectnessess | NA | NA | NA | NA | NA | -0.08 | -0.76 | 0.62 | Low | Imprecision |
| intravenous immunoglobulin | soc | -0.08 | -0.88 | 0.72 | Low | RoB,Inconsistency | NA | NA | NA | NA | NA | -0.08 | -0.88 | 0.72 | Very low | Imprecision |
| interferon beta | soc | 0.00 | -0.17 | 0.16 | Moderate | RoB | NA | NA | NA | NA | NA | 0.00 | -0.17 | 0.16 | Low | Imprecision |
| recombinant human gcsf | soc | -1.58 | -2.27 | -0.93 | Moderate | RoB | NA | NA | NA | NA | NA | -1.58 | -2.27 | -0.93 | Moderate | NA |

| | | Direct | | | | | Indirect | | | | | Network | | | | |
|---|---|---|---|---|---|---|---|---|---|---|---|---|---|---|---|---|
| Treatment 1 | Treatment 2 | EST* | LCrI* | UCrI* | Evidence | Reason | EST | LCrI | UCrI | Evidence | Reason | EST | LCrI | UCrI | Evidence | Reason |
| tocilizumab | soc | -0.41 | -0.87 | 0.07 | High | NA | NA | NA | NA | NA | NA | -0.41 | -0.87 | 0.07 | Moderate | Imprecision |
| vitamin d3 | soc | -0.86 | -1.81 | 0.00 | High | NA | NA | NA | NA | NA | NA | -0.86 | -1.81 | 0.00 | Moderate | Imprecision |
| baricitinib + remdesivir | soc | NA | NA | NA | NA | NA | -0.43 | -0.85 | -0.01 | Very low | NA | -0.43 | -0.85 | -0.01 | Very low | Imprecision |
| sulodexide | soc | -0.82 | -2.44 | 0.57 | Moderate | RoB | NA | NA | NA | NA | NA | -0.82 | -2.44 | 0.57 | Very low | Severe Imprecision |
| hydroxychloroquine | azithromycin | 0.19 | -0.62 | 1.04 | Moderate | RoB | -0.03 | -0.26 | 0.19 | Very low | Intransitivity | -0.02 | -0.24 | 0.20 | Very low | Imprecision |
| hydroxychloroquine + azithromycin | azithromycin | NA | NA | NA | NA | NA | 0.44 | -0.23 | 1.10 | Very low | Intransitivity | 0.44 | -0.23 | 1.10 | Very low | Imprecision |
| remdesivir | azithromycin | NA | NA | NA | NA | NA | 0.04 | -0.17 | 0.26 | Very low | NA | 0.04 | -0.17 | 0.26 | Very low | Imprecision |
| lopinavir/ritonavir | azithromycin | NA | NA | NA | NA | NA | 0.01 | -0.22 | 0.23 | Very low | NA | 0.01 | -0.22 | 0.23 | Very low | Imprecision |
| convalescent plasma | azithromycin | NA | NA | NA | NA | NA | -0.05 | -0.42 | 0.32 | Very low | NA | -0.05 | -0.42 | 0.32 | Very low | Imprecision |
| methylprednisolone | azithromycin | NA | NA | NA | NA | NA | 0.06 | -0.57 | 0.72 | Very low | NA | 0.06 | -0.57 | 0.72 | Very low | Imprecision |
| dexamethasone | azithromycin | NA | NA | NA | NA | NA | -0.40 | -0.66 | -0.14 | Very low | NA | -0.40 | -0.66 | -0.14 | Low | NA |
| hydrocortisone | azithromycin | NA | NA | NA | NA | NA | -0.09 | -0.80 | 0.63 | Very low | NA | -0.09 | -0.80 | 0.63 | Very low | Imprecision |
| intravenous immunoglobulin | azithromycin | NA | NA | NA | NA | NA | -0.10 | -0.91 | 0.72 | Very low | NA | -0.10 | -0.91 | 0.72 | Very low | Imprecision |
| interferon beta | azithromycin | NA | NA | NA | NA | NA | -0.01 | -0.25 | 0.22 | Very low | NA | -0.01 | -0.25 | 0.22 | Very low | Imprecision |
| recombinant human gcsf | azithromycin | NA | NA | NA | NA | NA | -1.60 | -2.30 | -0.92 | Very low | Intransitivity | -1.60 | -2.30 | -0.92 | Very low | NA |
| tocilizumab | azithromycin | NA | NA | NA | NA | NA | -0.42 | -0.92 | 0.08 | Very low | NA | -0.42 | -0.92 | 0.08 | Very low | Imprecision |

| | | Direct | | | | | Indirect | | | | | Network | | | | |
|---|---|---|---|---|---|---|---|---|---|---|---|---|---|---|---|---|
| **Treatment 1** | **Treatment 2** | **EST*** | **LCrI*** | **UCrI*** | **Evidence** | **Reason** | **EST** | **LCrI** | **UCrI** | **Evidence** | **Reason** | **EST** | **LCrI** | **UCrI** | **Evidence** | **Reason** |
| vitamin d3 | azithromycin | NA | NA | NA | NA | NA | -0.88 | -1.84 | 0.01 | Very low | NA | -0.88 | -1.84 | 0.01 | Very low | Severe Imprecision |
| baricitinib + remdesivir | azithromycin | NA | NA | NA | NA | NA | -0.44 | -0.90 | 0.01 | Very low | NA | -0.44 | -0.90 | 0.01 | Very low | Imprecision |
| sulodexide | azithromycin | NA | NA | NA | NA | NA | -0.83 | -2.47 | 0.57 | Very low | Intransitivity | -0.83 | -2.47 | 0.57 | Very low | Severe Imprecision |
| hydroxychloroquine + azithromycin | hydroxychloroquine | 0.47 | -0.18 | 1.10 | Moderate | RoB | NA | NA | NA | Moderate | NA | 0.47 | -0.18 | 1.10 | Low | Imprecision |
| remdesivir | hydroxychloroquine | 0.33 | 0.08 | 0.60 | MA | NA | -0.36 | -0.66 | -0.06 | Very low | NA | 0.07 | -0.11 | 0.25 | Very low | Imprecision |
| lopinavir/ritonavir | hydroxychloroquine | 0.04 | -0.15 | 0.23 | Moderate | RoB | -0.16 | -0.84 | 0.50 | Low | Intransitivity | 0.03 | -0.15 | 0.21 | Low | Imprecision |
| convalescent plasma | hydroxychloroquine | NA | NA | NA | NA | NA | -0.03 | -0.39 | 0.33 | Moderate | NA | -0.03 | -0.39 | 0.33 | Low | Imprecision |
| methylprednisolone | hydroxychloroquine | NA | NA | NA | NA | NA | 0.09 | -0.54 | 0.74 | Moderate | NA | 0.09 | -0.54 | 0.74 | Low | Imprecision |
| dexamethasone | hydroxychloroquine | -0.57 | -0.84 | -0.30 | Moderate | RoB | 0.04 | -0.44 | 0.52 | Very low | Intransitivity | -0.38 | -0.61 | -0.15 | Moderate | NA |
| hydrocortisone | hydroxychloroquine | NA | NA | NA | NA | NA | -0.07 | -0.77 | 0.64 | Low | Intransitivity | -0.07 | -0.77 | 0.64 | Very low | Imprecision |
| intravenous immunoglobulin | hydroxychloroquine | NA | NA | NA | NA | NA | -0.08 | -0.88 | 0.74 | Very low | Intransitivity | -0.08 | -0.88 | 0.74 | Very low | Imprecision |
| interferon beta | hydroxychloroquine | 0.22 | -0.06 | 0.50 | Moderate | RoB | -0.75 | -1.37 | -0.15 | Moderate | NA | 0.01 | -0.20 | 0.21 | Low | Imprecision |
| recombinant human gcsf | hydroxychloroquine | NA | NA | NA | NA | NA | -1.58 | -2.27 | -0.90 | Moderate | NA | -1.58 | -2.27 | -0.90 | Moderate | NA |
| tocilizumab | hydroxychloroquine | NA | NA | NA | NA | NA | -0.40 | -0.89 | 0.10 | Moderate | NA | -0.40 | -0.89 | 0.10 | Low | Imprecision |
| vitamin d3 | hydroxychloroquine | NA | NA | NA | NA | NA | -0.86 | -1.81 | 0.02 | Low | Intransitivity | -0.86 | -1.81 | 0.02 | Low | NA |
| baricitinib + remdesivir | hydroxychloroquine | NA | NA | NA | NA | NA | -0.42 | -0.86 | 0.02 | Very low | NA | -0.42 | -0.86 | 0.02 | Very low | Imprecision |

| | | Direct | | | | | Indirect | | | | | Network | | | | |
|---|---|---|---|---|---|---|---|---|---|---|---|---|---|---|---|---|
| **Treatment 1** | **Treatment 2** | **EST*** | **LCrI*** | **UCrI*** | **Evidence** | **Reason** | **EST** | **LCrI** | **UCrI** | **Evidence** | **Reason** | **EST** | **LCrI** | **UCrI** | **Evidence** | **Reason** |
| sulodexide | hydroxychloroquine | NA | NA | NA | NA | NA | -0.81 | -2.44 | 0.59 | Moderate | NA | -0.81 | -2.44 | 0.59 | Very low | Severe Imprecision |
| remdesivir | hydroxychloroquine + azithromycin | NA | NA | NA | NA | NA | -0.40 | -1.05 | 0.26 | Very low | Intransitivity | -0.40 | -1.05 | 0.26 | Very low | Imprecision |
| lopinavir/ritonavir | hydroxychloroquine + azithromycin | NA | NA | NA | NA | NA | -0.44 | -1.09 | 0.22 | Low | Intransitivity | -0.44 | -1.09 | 0.22 | Very low | Imprecision |
| convalescent plasma | hydroxychloroquine + azithromycin | NA | NA | NA | NA | NA | -0.50 | -1.22 | 0.23 | Low | Intransitivity | -0.50 | -1.22 | 0.23 | Very low | Imprecision |
| methylprednisolone | hydroxychloroquine + azithromycin | NA | NA | NA | NA | NA | -0.38 | -1.26 | 0.52 | Moderate | NA | -0.38 | -1.26 | 0.52 | Low | Imprecision |
| dexamethasone | hydroxychloroquine + azithromycin | NA | NA | NA | NA | NA | -0.84 | -1.50 | -0.17 | Very low | Intransitivity | -0.84 | -1.50 | -0.17 | Very low | NA |
| hydrocortisone | hydroxychloroquine + azithromycin | NA | NA | NA | NA | NA | -0.53 | -1.47 | 0.41 | Low | Intransitivity | -0.53 | -1.47 | 0.41 | Very low | Imprecision |
| intravenous immunoglobulin | hydroxychloroquine + azithromycin | NA | NA | NA | NA | NA | -0.54 | -1.56 | 0.49 | Very low | Intransitivity | -0.54 | -1.56 | 0.49 | Very low | Imprecision |
| interferon beta | hydroxychloroquine + azithromycin | NA | NA | NA | NA | NA | -0.46 | -1.11 | 0.21 | Moderate | NA | -0.46 | -1.11 | 0.21 | Low | Imprecision |
| recombinant human gcsf | hydroxychloroquine + azithromycin | NA | NA | NA | NA | NA | -2.04 | -2.98 | -1.12 | Moderate | NA | -2.04 | -2.98 | -1.12 | Moderate | NA |
| tocilizumab | hydroxychloroquine + azithromycin | NA | NA | NA | NA | NA | -0.86 | -1.65 | -0.06 | Moderate | NA | -0.86 | -1.65 | -0.06 | Moderate | NA |

| | | Direct | | | | | Indirect | | | | | Network | | | | |
|---|---|---|---|---|---|---|---|---|---|---|---|---|---|---|---|---|
| **Treatment 1** | **Treatment 2** | **EST*** | **LCrI*** | **UCrI*** | **Evidence** | **Reason** | **EST** | **LCrI** | **UCrI** | **Evidence** | **Reason** | **EST** | **LCrI** | **UCrI** | **Evidence** | **Reason** |
| vitamin d3 | hydroxychloroquine + azithromycin | NA | NA | NA | NA | NA | -1.32 | -2.46 | -0.24 | Low | Intransitivity | -1.32 | -2.46 | -0.24 | Low | NA |
| baricitinib + remdesivir | hydroxychloroquine + azithromycin | NA | NA | NA | NA | NA | -0.88 | -1.65 | -0.11 | Very low | NA | -0.88 | -1.65 | -0.11 | Very low | NA |
| sulodexide | hydroxychloroquine + azithromycin | NA | NA | NA | NA | NA | -1.28 | -3.03 | 0.26 | Moderate | NA | -1.28 | -3.03 | 0.26 | Very low | Severe Imprecision |
| lopinavir/ritonavir | remdesivir | -0.22 | -0.44 | -0.01 | NA | NA | 0.39 | 0.08 | 0.70 | Very low | NA | -0.04 | -0.21 | 0.13 | Very low | Imprecision |
| convalescent plasma | remdesivir | NA | NA | NA | NA | NA | -0.10 | -0.45 | 0.26 | Very low | NA | -0.10 | -0.45 | 0.26 | Very low | Imprecision |
| methylprednisolone | remdesivir | NA | NA | NA | NA | NA | 0.02 | -0.60 | 0.67 | Very low | NA | 0.02 | -0.60 | 0.67 | Very low | Imprecision |
| dexamethasone | remdesivir | NA | NA | NA | NA | NA | -0.44 | -0.68 | -0.21 | Very low | NA | -0.44 | -0.68 | -0.21 | Very low | NA |
| hydrocortisone | remdesivir | NA | NA | NA | NA | NA | -0.14 | -0.83 | 0.57 | Very low | NA | -0.14 | -0.83 | 0.57 | Very low | Imprecision |
| intravenous immunoglobulin | remdesivir | NA | NA | NA | NA | NA | -0.14 | -0.95 | 0.67 | Very low | NA | -0.14 | -0.95 | 0.67 | Very low | Imprecision |
| interferon beta | remdesivir | -0.11 | -0.30 | 0.07 | NA | NA | -0.43 | -1.04 | 0.18 | Very low | NA | -0.06 | -0.24 | 0.12 | Very low | Imprecision |
| recombinant human gcsf | remdesivir | NA | NA | NA | NA | NA | -1.64 | -2.34 | -0.97 | Very low | Intransitivity | -1.64 | -2.34 | -0.97 | Very low | NA |
| tocilizumab | remdesivir | NA | NA | NA | NA | NA | -0.46 | -0.95 | 0.03 | Very low | NA | -0.46 | -0.95 | 0.03 | Very low | Imprecision |
| vitamin d3 | remdesivir | NA | NA | NA | NA | NA | -0.92 | -1.88 | -0.04 | Very low | NA | -0.92 | -1.88 | -0.04 | Very low | Severe Imprecision |
| baricitinib + remdesivir | remdesivir | -0.48 | -0.89 | -0.09 | Moderate | RoB | NA | NA | NA | NA | NA | -0.48 | -0.89 | -0.09 | Moderate | NA |
| sulodexide | remdesivir | NA | NA | NA | NA | NA | -0.88 | -2.51 | 0.52 | Very low | Intransitivity | -0.88 | -2.51 | 0.52 | Very low | Severe Imprecision |

| | | Direct | | | | | Indirect | | | | | Network | | | | |
|---|---|---|---|---|---|---|---|---|---|---|---|---|---|---|---|---|
| **Treatment 1** | **Treatment 2** | **EST*** | **LCrI*** | **UCrI*** | **Evidence** | **Reason** | **EST** | **LCrI** | **UCrI** | **Evidence** | **Reason** | **EST** | **LCrI** | **UCrI** | **Evidence** | **Reason** |
| convalescent plasma | lopinavir/ritonavir | NA | NA | NA | NA | NA | -0.06 | -0.42 | 0.30 | Moderate | NA | -0.06 | -0.42 | 0.30 | Low | Imprecision |
| methylprednisolone | lopinavir/ritonavir | NA | NA | NA | NA | NA | 0.06 | -0.56 | 0.71 | Moderate | NA | 0.06 | -0.56 | 0.71 | Low | Imprecision |
| dexamethasone | lopinavir/ritonavir | -0.56 | -0.82 | -0.30 | Moderate | RoB | 0.00 | -0.48 | 0.48 | Low | NA | -0.40 | -0.63 | -0.18 | Moderate | NA |
| hydrocortisone | lopinavir/ritonavir | NA | NA | NA | NA | NA | -0.10 | -0.80 | 0.61 | Moderate | NA | -0.10 | -0.80 | 0.61 | Low | Imprecision |
| intravenous immunoglobulin | lopinavir/ritonavir | NA | NA | NA | NA | NA | -0.10 | -0.91 | 0.71 | Low | NA | -0.10 | -0.91 | 0.71 | Very low | Imprecision |
| interferon beta | lopinavir/ritonavir | 0.11 | -0.12 | 0.35 | NA | NA | -0.78 | -1.40 | -0.18 | Moderate | NA | -0.02 | -0.22 | 0.18 | Low | Imprecision |
| recombinant human gcsf | lopinavir/ritonavir | NA | NA | NA | NA | NA | -1.60 | -2.30 | -0.93 | Low | Intransitivity | -1.60 | -2.30 | -0.93 | Low | NA |
| tocilizumab | lopinavir/ritonavir | NA | NA | NA | NA | NA | -0.42 | -0.91 | 0.07 | Moderate | NA | -0.42 | -0.91 | 0.07 | Low | Imprecision |
| vitamin d3 | lopinavir/ritonavir | NA | NA | NA | NA | NA | -0.88 | -1.84 | 0.00 | Moderate | NA | -0.88 | -1.84 | 0.00 | Moderate | NA |
| baricitinib + remdesivir | lopinavir/ritonavir | NA | NA | NA | NA | NA | -0.44 | -0.88 | -0.01 | Very low | NA | -0.44 | -0.88 | -0.01 | Very low | Imprecision |
| sulodexide | lopinavir/ritonavir | NA | NA | NA | NA | NA | -0.84 | -2.46 | 0.56 | Moderate | NA | -0.84 | -2.46 | 0.56 | Very low | Severe Imprecision |
| methylprednisolone | convalescent plasma | NA | NA | NA | NA | NA | 0.12 | -0.58 | 0.83 | Moderate | NA | 0.12 | -0.58 | 0.83 | Low | Imprecision |
| dexamethasone | convalescent plasma | NA | NA | NA | NA | NA | -0.34 | -0.73 | 0.04 | Low | NA | -0.34 | -0.73 | 0.04 | Very low | Imprecision |
| hydrocortisone | convalescent plasma | NA | NA | NA | NA | NA | -0.04 | -0.80 | 0.73 | Moderate | NA | -0.04 | -0.80 | 0.73 | Low | Imprecision |
| intravenous immunoglobulin | convalescent plasma | NA | NA | NA | NA | NA | -0.04 | -0.91 | 0.82 | Low | NA | -0.04 | -0.91 | 0.82 | Very low | Imprecision |
| interferon beta | convalescent plasma | NA | NA | NA | NA | NA | 0.04 | -0.33 | 0.41 | Moderate | NA | 0.04 | -0.33 | 0.41 | Low | Imprecision |
| recombinant human gcsf | convalescent plasma | NA | NA | NA | NA | NA | -1.55 | -2.30 | -0.81 | Low | Intransitivity | -1.55 | -2.30 | -0.81 | Low | NA |

| | | Direct | | | | | Indirect | | | | | Network | | | | |
|---|---|---|---|---|---|---|---|---|---|---|---|---|---|---|---|---|
| **Treatment 1** | **Treatment 2** | **EST*** | **LCrI*** | **UCrI*** | **Evidence** | **Reason** | **EST** | **LCrI** | **UCrI** | **Evidence** | **Reason** | **EST** | **LCrI** | **UCrI** | **Evidence** | **Reason** |
| tocilizumab | convalescent plasma | NA | NA | NA | NA | NA | -0.36 | -0.94 | 0.21 | Moderate | NA | -0.36 | -0.94 | 0.21 | Low | Imprecision |
| vitamin d3 | convalescent plasma | NA | NA | NA | NA | NA | -0.82 | -1.82 | 0.11 | Moderate | NA | -0.82 | -1.82 | 0.11 | Very low | Severe Imprecision |
| baricitinib + remdesivir | convalescent plasma | NA | NA | NA | NA | NA | -0.39 | -0.92 | 0.15 | Very low | NA | -0.39 | -0.92 | 0.15 | Very low | Imprecision |
| sulodexide | convalescent plasma | NA | NA | NA | NA | NA | -0.78 | -2.43 | 0.65 | Low | Intransitivity | -0.78 | -2.43 | 0.65 | Very low | Severe Imprecision |
| dexamethason e | methylpredni solone | NA | NA | NA | NA | NA | -0.46 | -1.13 | 0.17 | Low | NA | -0.46 | -1.13 | 0.17 | Very low | Imprecision |
| hydrocortisone | methylpredni solone | NA | NA | NA | NA | NA | -0.15 | -1.06 | 0.72 | Moderate | NA | -0.15 | -1.06 | 0.72 | Low | Imprecision |
| intravenous immunoglobul in | methylpredni solone | NA | NA | NA | NA | NA | -0.16 | -1.18 | 0.84 | Low | NA | -0.16 | -1.18 | 0.84 | Very low | Imprecision |
| interferon beta | methylpredni solone | NA | NA | NA | NA | NA | -0.08 | -0.73 | 0.55 | Moderate | NA | -0.08 | -0.73 | 0.55 | Low | Imprecision |
| recombinant human gcsf | methylpredni solone | NA | NA | NA | NA | NA | -1.66 | -2.62 | -0.74 | Moderate | NA | -1.66 | -2.62 | -0.74 | Moderate | NA |
| tocilizumab | methylpredni solone | NA | NA | NA | NA | NA | -0.48 | -1.27 | 0.29 | Moderate | NA | -0.48 | -1.27 | 0.29 | Low | Imprecision |
| vitamin d3 | methylpredni solone | NA | NA | NA | NA | NA | -0.95 | -2.08 | 0.12 | Moderate | NA | -0.95 | -2.08 | 0.12 | Very low | Severe Imprecision |
| baricitinib + remdesivir | methylpredni solone | NA | NA | NA | NA | NA | -0.50 | -1.27 | 0.23 | Very low | NA | -0.50 | -1.27 | 0.23 | Very low | Imprecision |
| sulodexide | methylpredni solone | NA | NA | NA | NA | NA | -0.91 | -2.63 | 0.62 | Moderate | NA | -0.91 | -2.63 | 0.62 | Very low | Severe Imprecision |
| hydrocortisone | dexamethaso ne | NA | NA | NA | NA | NA | 0.31 | -0.40 | 1.03 | Low | NA | 0.31 | -0.40 | 1.03 | Very low | Imprecision |
| intravenous immunoglobul in | dexamethaso ne | NA | NA | NA | NA | NA | 0.30 | -0.52 | 1.13 | Low | NA | 0.30 | -0.52 | 1.13 | Very low | Imprecision |

| | | Direct | | | | | Indirect | | | | | Network | | | | |
|---|---|---|---|---|---|---|---|---|---|---|---|---|---|---|---|---|
| Treatment 1 | Treatment 2 | EST* | LCrI* | UCrI* | Evidence | Reason | EST | LCrI | UCrI | Evidence | Reason | EST | LCrI | UCrI | Evidence | Reason |
| interferon beta | dexamethasone | NA | NA | NA | NA | NA | 0.38 | 0.13 | 0.64 | Low | NA | 0.38 | 0.13 | 0.64 | Very low | Imprecision |
| recombinant human gcsf | dexamethasone | NA | NA | NA | NA | NA | -1.20 | -1.91 | -0.52 | Very low | Intransitivity | -1.20 | -1.91 | -0.52 | Very low | NA |
| tocilizumab | dexamethasone | NA | NA | NA | NA | NA | -0.02 | -0.53 | 0.49 | Low | NA | -0.02 | -0.53 | 0.49 | Very low | Imprecision |
| vitamin d3 | dexamethasone | NA | NA | NA | NA | NA | -0.48 | -1.44 | 0.41 | Low | NA | -0.48 | -1.44 | 0.41 | Very low | Severe Imprecision |
| baricitinib + remdesivir | dexamethasone | NA | NA | NA | NA | NA | -0.04 | -0.51 | 0.42 | Very low | NA | -0.04 | -0.51 | 0.42 | Very low | Imprecision |
| sulodexide | dexamethasone | NA | NA | NA | NA | NA | -0.44 | -2.07 | 0.97 | Very low | Intransitivity | -0.44 | -2.07 | 0.97 | Very low | Severe Imprecision |
| intravenous immunoglobulin | hydrocortisone | NA | NA | NA | NA | NA | -0.01 | -1.06 | 1.05 | Low | NA | -0.01 | -1.06 | 1.05 | Very low | Imprecision |
| interferon beta | hydrocortisone | NA | NA | NA | NA | NA | 0.08 | -0.64 | 0.78 | Moderate | NA | 0.08 | -0.64 | 0.78 | Low | Imprecision |
| recombinant human gcsf | hydrocortisone | NA | NA | NA | NA | NA | -1.50 | -2.51 | -0.56 | Low | Intransitivity | -1.50 | -2.51 | -0.56 | Low | NA |
| tocilizumab | hydrocortisone | NA | NA | NA | NA | NA | -0.33 | -1.17 | 0.50 | Moderate | NA | -0.33 | -1.17 | 0.50 | Low | Imprecision |
| vitamin d3 | hydrocortisone | NA | NA | NA | NA | NA | -0.79 | -1.95 | 0.32 | Moderate | NA | -0.79 | -1.95 | 0.32 | Very low | Severe Imprecision |
| baricitinib + remdesivir | hydrocortisone | NA | NA | NA | NA | NA | -0.35 | -1.16 | 0.45 | Very low | NA | -0.35 | -1.16 | 0.45 | Very low | Imprecision |
| sulodexide | hydrocortisone | NA | NA | NA | NA | NA | -0.75 | -2.51 | 0.81 | Low | Intransitivity | -0.75 | -2.51 | 0.81 | Very low | Severe Imprecision |
| interferon beta | intravenous immunoglobulin | NA | NA | NA | NA | NA | 0.08 | -0.73 | 0.89 | Low | NA | 0.08 | -0.73 | 0.89 | Very low | Imprecision |

| | | Direct | | | | | Indirect | | | | | Network | | | | |
|---|---|---|---|---|---|---|---|---|---|---|---|---|---|---|---|---|
| Treatment 1 | Treatment 2 | EST* | LCrI* | UCrI* | Evidence | Reason | EST | LCrI | UCrI | Evidence | Reason | EST | LCrI | UCrI | Evidence | Reason |
| recombinant human gcsf | intravenous immunoglobulin | NA | NA | NA | NA | NA | -1.50 | -2.55 | -0.47 | Very low | Intransitivity | -1.50 | -2.55 | -0.47 | Very low | NA |
| tocilizumab | intravenous immunoglobulin | NA | NA | NA | NA | NA | -0.32 | -1.25 | 0.60 | Low | NA | -0.32 | -1.25 | 0.60 | Very low | Imprecision |
| vitamin d3 | intravenous immunoglobulin | NA | NA | NA | NA | NA | -0.78 | -2.01 | 0.39 | Low | NA | -0.78 | -2.01 | 0.39 | Very low | Severe Imprecision |
| baricitinib + remdesivir | intravenous immunoglobulin | NA | NA | NA | NA | NA | -0.34 | -1.25 | 0.55 | Very low | NA | -0.34 | -1.25 | 0.55 | Very low | Imprecision |
| sulodexide | intravenous immunoglobulin | NA | NA | NA | NA | NA | -0.74 | -2.54 | 0.87 | Very low | Intransitivity | -0.74 | -2.54 | 0.87 | Very low | Severe Imprecision |
| recombinant human gcsf | interferon beta | NA | NA | NA | NA | NA | -1.58 | -2.29 | -0.90 | Moderate | NA | -1.58 | -2.29 | -0.90 | Moderate | NA |
| tocilizumab | interferon beta | NA | NA | NA | NA | NA | -0.40 | -0.90 | 0.10 | Moderate | NA | -0.40 | -0.90 | 0.10 | Low | Imprecision |
| vitamin d3 | interferon beta | NA | NA | NA | NA | NA | -0.86 | -1.82 | 0.02 | Moderate | NA | -0.86 | -1.82 | 0.02 | Very low | Severe Imprecision |
| baricitinib + remdesivir | interferon beta | NA | NA | NA | NA | NA | -0.42 | -0.87 | 0.01 | Very low | NA | -0.42 | -0.87 | 0.01 | Very low | Imprecision |
| sulodexide | interferon beta | NA | NA | NA | NA | NA | -0.82 | -2.45 | 0.58 | Low | Intransitivity | -0.82 | -2.45 | 0.58 | Very low | Severe Imprecision |
| tocilizumab | recombinant human gcsf | NA | NA | NA | NA | NA | 1.18 | 0.37 | 2.01 | Moderate | NA | 1.18 | 0.37 | 2.01 | Moderate | NA |
| vitamin d3 | recombinant human gcsf | NA | NA | NA | NA | NA | 0.72 | -0.43 | 1.82 | Low | Intransitivity | 0.72 | -0.43 | 1.82 | Very low | Imprecision |
| baricitinib + remdesivir | recombinant human gcsf | NA | NA | NA | NA | NA | 1.16 | 0.37 | 1.96 | Very low | NA | 1.16 | 0.37 | 1.96 | Very low | |

| | | Direct | | | | | Indirect | | | | | Network | | | | |
|---|---|---|---|---|---|---|---|---|---|---|---|---|---|---|---|---|
| **Treatment 1** | **Treatment 2** | **EST*** | **LCrI*** | **UCrI*** | **Evidence** | **Reason** | **EST** | **LCrI** | **UCrI** | **Evidence** | **Reason** | **EST** | **LCrI** | **UCrI** | **Evidence** | **Reason** |
| sulodexide | recombinant human gcsf | NA | NA | NA | NA | NA | 0.76 | -0.98 | 2.31 | Moderate | NA | 0.76 | -0.98 | 2.31 | Very low | Severe Imprecision |
| vitamin d3 | tocilizumab | NA | NA | NA | NA | NA | -0.46 | -1.51 | 0.53 | High | NA | -0.46 | -1.51 | 0.53 | Low | Severe Imprecision |
| baricitinib + remdesivir | tocilizumab | NA | NA | NA | NA | NA | -0.02 | -0.65 | 0.61 | Very low | NA | -0.02 | -0.65 | 0.61 | Very low | Imprecision |
| sulodexide | tocilizumab | NA | NA | NA | NA | NA | -0.42 | -2.11 | 1.05 | Moderate | NA | -0.42 | -2.11 | 1.05 | Very low | Severe Imprecision |
| baricitinib + remdesivir | vitamin d3 | NA | NA | NA | NA | NA | 0.44 | -0.53 | 1.47 | Very low | NA | 0.44 | -0.53 | 1.47 | Very low | Imprecision |
| sulodexide | vitamin d3 | NA | NA | NA | NA | NA | 0.04 | -1.79 | 1.73 | Low | Intransitivity | 0.04 | -1.79 | 1.73 | Very low | Severe Imprecision |
| sulodexide | baricitinib + remdesivir | NA | NA | NA | NA | NA | -0.39 | -2.07 | 1.06 | Very low | NA | -0.39 | -2.07 | 1.06 | Very low | Severe Imprecision |

*EST: estimate; LCrI: lower credible interval; UCrI: upper credible interval.

#### Table S11. Network meta-analysis results of the primary analysis (log odds ratio, log OR) and evaluation of certainty of evidence (discharge).

| | | Direct | | | | | Indirect | | | | | Network | | | | |
|---|---|---|---|---|---|---|---|---|---|---|---|---|---|---|---|---|
| **Treatment 1** | **Treatment 2** | **EST*** | **LCrI*** | **UCrI*** | **Evidence** | **Reason** | **EST** | **LCrI** | **UCrI** | **Evidence** | **Reason** | **EST** | **LCrI** | **UCrI** | **Evidence** | **Reason** |
| azithromycin | soc | 0.03 | -0.07 | 0.12 | Moderate | RoB | -0.01 | -0.63 | 0.54 | Moderate | NA | 0.03 | -0.07 | 0.12 | Low | Imprecision |
| hydroxychloroquine | soc | -0.15 | -0.25 | -0.04 | Moderate | RoB | -0.09 | -0.72 | 0.61 | Moderate | NA | -0.14 | -0.24 | -0.03 | Moderate | NA |
| hydroxychloroquine + azithromycin | soc | -0.16 | -0.64 | 0.33 | Moderate | RoB | NA | NA | NA | NA | NA | -0.16 | -0.64 | 0.33 | Low | Imprecision |
| favipiravir | soc | 0.18 | -0.74 | 0.63 | Moderate | RoB | NA | NA | NA | NA | NA | 0.18 | -0.74 | 0.63 | Low | Imprecision |
| remdesivir | soc | 0.32 | 0.14 | 0.50 | Moderate | Publication bias | NA | NA | NA | NA | NA | 0.32 | 0.14 | 0.50 | Moderate | NA |
| lopinavir/ritonavir | soc | 0.27 | 0.15 | 0.38 | Low | RoB, Inconsistency | NA | NA | NA | NA | NA | 0.27 | 0.15 | 0.38 | Low | NA |
| convalescent plasma | soc | 0.09 | -0.29 | 0.47 | Moderate | RoB | NA | NA | NA | NA | NA | 0.09 | -0.29 | 0.47 | Low | Imprecision |
| dexamethasone | soc | 0.18 | 0.07 | 0.29 | Low | RoB, Inconsistency | NA | NA | NA | NA | NA | 0.18 | 0.07 | 0.29 | Low | NA |
| interferon beta | soc | 0.77 | 0.23 | 1.32 | Moderate | RoB | NA | NA | NA | NA | NA | 0.77 | 0.23 | 1.32 | Moderate | NA |
| tocilizumab | soc | 0.34 | 0.05 | 0.64 | Moderate | RoB | NA | NA | NA | NA | NA | 0.34 | 0.05 | 0.64 | Moderate | NA |
| baricitinib + remdesivir | soc | NA | NA | NA | NA | NA | 0.56 | 0.24 | 0.87 | Moderate | NA | 0.56 | 0.24 | 0.87 | Moderate | NA |
| hydroxychloroquine | azithromycin | -0.13 | -0.67 | 0.48 | Moderate | RoB | -0.17 | -0.31 | -0.03 | Low | Intransitivity | -0.17 | -0.31 | -0.02 | Low | Imprecision |
| hydroxychloroquine + azithromycin | azithromycin | NA | NA | NA | NA | NA | -0.19 | -0.68 | 0.31 | Low | Intransitivity | -0.19 | -0.68 | 0.31 | Very low | Imprecision |
| favipiravir | azithromycin | NA | NA | NA | NA | NA | 0.15 | -0.77 | 0.61 | Low | Intransitivity | 0.15 | -0.77 | 0.61 | Very low | Imprecision |
| remdesivir | azithromycin | NA | NA | NA | NA | NA | 0.29 | 0.09 | 0.49 | Moderate | NA | 0.29 | 0.09 | 0.49 | Moderate | NA |

| | | Direct | | | | | Indirect | | | | | Network | | | | |
|---|---|---|---|---|---|---|---|---|---|---|---|---|---|---|---|---|
| **Treatment 1** | **Treatment 2** | **EST*** | **LCrI*** | **UCrI*** | **Evidence** | **Reason** | **EST** | **LCrI** | **UCrI** | **Evidence** | **Reason** | **EST** | **LCrI** | **UCrI** | **Evidence** | **Reason** |
| lopinavir/ritonavir | azithromycin | NA | NA | NA | NA | NA | 0.24 | 0.09 | 0.39 | Low | NA | 0.24 | 0.09 | 0.39 | Low | NA |
| convalescent plasma | azithromycin | NA | NA | NA | NA | NA | 0.06 | -0.33 | 0.45 | Moderate | NA | 0.06 | -0.33 | 0.45 | Low | Imprecision |
| dexamethasone | azithromycin | NA | NA | NA | NA | NA | 0.15 | 0.01 | 0.30 | Low | NA | 0.15 | 0.01 | 0.30 | Very low | Imprecision |
| interferon beta | azithromycin | NA | NA | NA | NA | NA | 0.74 | 0.19 | 1.30 | Moderate | NA | 0.74 | 0.19 | 1.30 | Moderate | NA |
| tocilizumab | azithromycin | NA | NA | NA | NA | NA | 0.31 | 0.00 | 0.63 | Low | Intransitivity | 0.31 | 0.00 | 0.63 | Low | NA |
| baricitinib + remdesivir | azithromycin | NA | NA | NA | NA | NA | 0.53 | 0.20 | 0.86 | Moderate | NA | 0.53 | 0.20 | 0.86 | Moderate | NA |
| hydroxychloroquine + azithromycin | hydroxychloroquine | -0.03 | -0.50 | 0.47 | Moderate | RoB | NA | NA | NA | Moderate | NA | -0.03 | -0.50 | 0.47 | Low | Imprecision |
| favipiravir | hydroxychloroquine | NA | NA | NA | NA | NA | 0.31 | -0.60 | 0.78 | Moderate | NA | 0.31 | -0.60 | 0.78 | Low | Imprecision |
| remdesivir | hydroxychloroquine | NA | NA | NA | NA | NA | 0.45 | 0.25 | 0.66 | Moderate | NA | 0.45 | 0.25 | 0.66 | Moderate | NA |
| lopinavir/ritonavir | hydroxychloroquine | 0.40 | 0.26 | 0.55 | Moderate | RoB | 0.51 | 0.11 | 1.05 | Low | NA | 0.40 | 0.27 | 0.54 | Low | NA |
| convalescent plasma | hydroxychloroquine | NA | NA | NA | NA | NA | 0.23 | -0.17 | 0.62 | Low | Intransitivity | 0.23 | -0.17 | 0.62 | Very low | Imprecision |
| dexamethasone | hydroxychloroquine | 0.33 | 0.20 | 0.47 | Moderate | RoB | 0.51 | 0.02 | 1.30 | Very low | Intransitivity | 0.32 | 0.19 | 0.45 | Moderate | NA |
| interferon beta | hydroxychloroquine | NA | NA | NA | NA | NA | 0.91 | 0.36 | 1.47 | Low | Intransitivity | 0.91 | 0.36 | 1.47 | Low | NA |
| tocilizumab | hydroxychloroquine | NA | NA | NA | NA | NA | 0.48 | 0.17 | 0.80 | Moderate | NA | 0.48 | 0.17 | 0.80 | Moderate | NA |
| baricitinib + remdesivir | hydroxychloroquine | NA | NA | NA | NA | NA | 0.70 | 0.37 | 1.03 | Moderate | NA | 0.70 | 0.37 | 1.03 | Moderate | NA |
| favipiravir | hydroxychloroquine + azithromycin | NA | NA | NA | NA | NA | 0.31 | -0.67 | 0.99 | Moderate | NA | 0.31 | -0.67 | 0.99 | Low | Imprecision |

| | | Direct | | | | | Indirect | | | | | Network | | | | |
|---|---|---|---|---|---|---|---|---|---|---|---|---|---|---|---|---|
| Treatment 1 | Treatment 2 | EST* | LCrI* | UCrI* | Evidence | Reason | EST | LCrI | UCrI | Evidence | Reason | EST | LCrI | UCrI | Evidence | Reason |
| remdesivir | hydroxychloroquine + azithromycin | NA | NA | NA | NA | NA | 0.48 | -0.04 | 0.99 | Low | Intransitivity | 0.48 | -0.04 | 0.99 | Very low | Imprecision |
| lopinavir/ritonavir | hydroxychloroquine + azithromycin | NA | NA | NA | NA | NA | 0.43 | -0.07 | 0.92 | Low | NA | 0.43 | -0.07 | 0.92 | Very low | Imprecision |
| convalescent plasma | hydroxychloroquine + azithromycin | NA | NA | NA | NA | NA | 0.25 | -0.37 | 0.86 | Low | Intransitivity | 0.25 | -0.37 | 0.86 | Very low | Imprecision |
| dexamethasone | hydroxychloroquine + azithromycin | NA | NA | NA | NA | NA | 0.35 | -0.16 | 0.83 | Very low | Intransitivity | 0.35 | -0.16 | 0.83 | Very low | Imprecision |
| interferon beta | hydroxychloroquine + azithromycin | NA | NA | NA | NA | NA | 0.93 | 0.20 | 1.66 | Low | Intransitivity | 0.93 | 0.20 | 1.66 | Very low | Imprecision |
| tocilizumab | hydroxychloroquine + azithromycin | NA | NA | NA | NA | NA | 0.50 | -0.07 | 1.06 | Moderate | NA | 0.50 | -0.07 | 1.06 | Low | Imprecision |
| baricitinib + remdesivir | hydroxychloroquine + azithromycin | NA | NA | NA | NA | NA | 0.72 | 0.14 | 1.30 | Moderate | NA | 0.72 | 0.14 | 1.30 | Moderate | NA |
| remdesivir | favipiravir | NA | NA | NA | NA | NA | 0.13 | -0.32 | 1.07 | Low | Intransitivity | 0.13 | -0.32 | 1.07 | Very low | Imprecision |
| lopinavir/ritonavir | favipiravir | NA | NA | NA | NA | NA | 0.09 | -0.37 | 1.00 | Low | NA | 0.09 | -0.37 | 1.00 | Very low | Imprecision |
| convalescent plasma | favipiravir | NA | NA | NA | NA | NA | -0.07 | -0.68 | 0.88 | Low | Intransitivity | -0.07 | -0.68 | 0.88 | Very low | Imprecision |
| dexamethasone | favipiravir | NA | NA | NA | NA | NA | 0.01 | -0.46 | 0.92 | Very low | Intransitivity | 0.01 | -0.46 | 0.92 | Very low | Imprecision |
| interferon beta | favipiravir | NA | NA | NA | NA | NA | 0.62 | -0.11 | 1.63 | Low | Intransitivity | 0.62 | -0.11 | 1.63 | Very low | Severe Imprecision |
| tocilizumab | favipiravir | NA | NA | NA | NA | NA | 0.18 | -0.37 | 1.12 | Moderate | NA | 0.18 | -0.37 | 1.12 | Low | Imprecision |
| baricitinib + remdesivir | favipiravir | NA | NA | NA | NA | NA | 0.39 | -0.14 | 1.34 | Moderate | NA | 0.39 | -0.14 | 1.34 | Low | Imprecision |

| | | Direct | | | | | Indirect | | | | | Network | | | | |
|---|---|---|---|---|---|---|---|---|---|---|---|---|---|---|---|---|
| **Treatment 1** | **Treatment 2** | **EST*** | **LCrI*** | **UCrI*** | **Evidence** | **Reason** | **EST** | **LCrI** | **UCrI** | **Evidence** | **Reason** | **EST** | **LCrI** | **UCrI** | **Evidence** | **Reason** |
| lopinavir/ritonavir | remdesivir | NA | NA | NA | NA | NA | -0.05 | -0.26 | 0.15 | Low | NA | -0.05 | -0.26 | 0.15 | Very low | Imprecision |
| convalescent plasma | remdesivir | NA | NA | NA | NA | NA | -0.23 | -0.65 | 0.19 | Moderate | NA | -0.23 | -0.65 | 0.19 | Low | Imprecision |
| dexamethasone | remdesivir | NA | NA | NA | NA | NA | -0.13 | -0.34 | 0.07 | Low | NA | -0.13 | -0.34 | 0.07 | Very low | Imprecision |
| interferon beta | remdesivir | NA | NA | NA | NA | NA | 0.45 | -0.12 | 1.03 | Moderate | NA | 0.45 | -0.12 | 1.03 | Low | Imprecision |
| tocilizumab | remdesivir | NA | NA | NA | NA | NA | 0.02 | -0.32 | 0.37 | Moderate | NA | 0.02 | -0.32 | 0.37 | Low | Imprecision |
| baricitinib + remdesivir | remdesivir | 0.24 | -0.02 | 0.50 | Moderate | RoB | NA | NA | NA | NA | NA | 0.24 | -0.02 | 0.50 | Low | Imprecision |
| convalescent plasma | lopinavir/ritonavir | NA | NA | NA | NA | NA | -0.18 | -0.58 | 0.22 | Low | NA | -0.18 | -0.58 | 0.22 | Very low | Imprecision |
| dexamethasone | lopinavir/ritonavir | -0.07 | -0.21 | 0.07 | Moderate | RoB | -0.01 | -0.68 | 0.81 | Low | NA | -0.09 | -0.22 | 0.05 | Very low | Imprecision |
| interferon beta | lopinavir/ritonavir | NA | NA | NA | NA | NA | 0.50 | -0.05 | 1.06 | Low | NA | 0.50 | -0.05 | 1.06 | Very low | Imprecision |
| tocilizumab | lopinavir/ritonavir | NA | NA | NA | NA | NA | 0.07 | -0.24 | 0.39 | Low | NA | 0.07 | -0.24 | 0.39 | Very low | Imprecision |
| baricitinib + remdesivir | lopinavir/ritonavir | NA | NA | NA | NA | NA | 0.29 | -0.04 | 0.62 | Low | NA | 0.29 | -0.04 | 0.62 | Very low | Imprecision |
| dexamethasone | convalescent plasma | NA | NA | NA | NA | NA | 0.09 | -0.30 | 0.49 | Low | NA | 0.09 | -0.30 | 0.49 | Very low | Imprecision |
| interferon beta | convalescent plasma | NA | NA | NA | NA | NA | 0.68 | 0.02 | 1.35 | Moderate | NA | 0.68 | 0.02 | 1.35 | Low | Imprecision |
| tocilizumab | convalescent plasma | NA | NA | NA | NA | NA | 0.25 | -0.23 | 0.74 | Low | Intransitivity | 0.25 | -0.23 | 0.74 | Very low | Imprecision |
| baricitinib + remdesivir | convalescent plasma | NA | NA | NA | NA | NA | 0.47 | -0.02 | 0.96 | Moderate | NA | 0.47 | -0.02 | 0.96 | Very low | Imprecision |
| interferon beta | dexamethasone | NA | NA | NA | NA | NA | 0.59 | 0.04 | 1.15 | Low | NA | 0.59 | 0.04 | 1.15 | Very low | Imprecision |
| tocilizumab | dexamethasone | NA | NA | NA | NA | NA | 0.16 | -0.15 | 0.48 | Very low | Intransitivity | 0.16 | -0.15 | 0.48 | Very low | Imprecision |
| baricitinib + remdesivir | dexamethasone | NA | NA | NA | NA | NA | 0.38 | 0.05 | 0.71 | Low | NA | 0.38 | 0.05 | 0.71 | Low | NA |

| | | Direct | | | | | Indirect | | | | | Network | | | | |
|---|---|---|---|---|---|---|---|---|---|---|---|---|---|---|---|---|
| **Treatment 1** | **Treatment 2** | **EST*** | **LCrI*** | **UCrI*** | **Evidence** | **Reason** | **EST** | **LCrI** | **UCrI** | **Evidence** | **Reason** | **EST** | **LCrI** | **UCrI** | **Evidence** | **Reason** |
| tocilizumab | interferon beta | NA | NA | NA | NA | NA | -0.43 | -1.05 | 0.19 | Low | Intransitivity | -0.43 | -1.05 | 0.19 | Very low | Imprecision |
| baricitinib + remdesivir | interferon beta | NA | NA | NA | NA | NA | -0.21 | -0.84 | 0.42 | Moderate | NA | -0.21 | -0.84 | 0.42 | Low | Imprecision |
| baricitinib + remdesivir | tocilizumab | NA | NA | NA | NA | NA | 0.22 | -0.22 | 0.65 | Moderate | NA | 0.22 | -0.22 | 0.65 | Low | Imprecision |

*EST: estimate; LCrI: lower credible interval; UCrI: upper credible interval.

#### Table S12. Network meta-analysis results of the primary analysis (log odds ratio, log OR) and evaluation of certainty of evidence (viral clearance).

| | | Direct | | | | | Indirect | | | | | Network | | | | |
|---|---|---|---|---|---|---|---|---|---|---|---|---|---|---|---|---|
| **Treatment 1** | **Treatment 2** | **EST*** | **LCrI*** | **UCrI*** | **Evidence** | **Reason** | **EST** | **LCrI** | **UCrI** | **Evidence** | **Reason** | **EST** | **LCrI** | **UCrI** | **Evidence** | **Reason** |
| hydroxychloroquine | soc | 0.20 | -0.05 | 0.46 | Low | RoB, Inconsistency | NA | NA | NA | NA | NA | 0.20 | -0.05 | 0.46 | Very low | Imprecision |
| nitazoxanide | soc | 0.54 | 0.13 | 1.03 | High | NA | NA | NA | NA | NA | NA | 0.54 | 0.13 | 1.03 | Low | NA |
| hydroxychloroquine + azithromycin | soc | -0.10 | -0.76 | 0.53 | High | NA | NA | NA | NA | NA | NA | -0.10 | -0.76 | 0.53 | Moderate | Imprecision |
| favipiravir | soc | 0.58 | -0.15 | 1.54 | Low | RoB, Inconsistency | NA | NA | NA | NA | NA | 0.58 | -0.15 | 1.54 | Very low | Imprecision |
| remdesivir | soc | 0.18 | -0.40 | 0.74 | Moderate | Publication bias | NA | NA | NA | NA | NA | 0.18 | -0.40 | 0.74 | Low | Imprecision |
| convalescent plasma | soc | 0.83 | 0.45 | 1.21 | Low | RoB, Inconsistency | NA | NA | NA | NA | NA | 0.83 | 0.45 | 1.21 | Low | NA |
| methylprednisolone | soc | 0.02 | -0.52 | 0.56 | High | NA | NA | NA | NA | NA | NA | 0.02 | -0.52 | 0.56 | Moderate | Imprecision |
| nitazoxanide | hydroxychloroquine | NA | NA | NA | NA | NA | 0.33 | -0.11 | 0.89 | Low | NA | 0.33 | -0.11 | 0.89 | Very low | Imprecision |
| hydroxychloroquine + azithromycin | hydroxychloroquine | -0.30 | -0.96 | 0.32 | High | NA | NA | NA | NA | NA | NA | -0.30 | -0.96 | 0.32 | Moderate | Imprecision |
| favipiravir | hydroxychloroquine | NA | NA | NA | NA | NA | 0.38 | -0.41 | 1.39 | Low | NA | 0.38 | -0.41 | 1.39 | Very low | Severe imprecision |
| remdesivir | hydroxychloroquine | NA | NA | NA | NA | NA | -0.03 | -0.65 | 0.58 | Very low | Intransitivity | -0.03 | -0.65 | 0.58 | Very low | Imprecision |
| convalescent plasma | hydroxychloroquine | NA | NA | NA | NA | NA | 0.62 | 0.17 | 1.08 | Low | NA | 0.62 | 0.17 | 1.08 | Low | NA |
| methylprednisolone | hydroxychloroquine | NA | NA | NA | NA | NA | -0.18 | -0.78 | 0.42 | Low | NA | -0.18 | -0.78 | 0.42 | Very low | Imprecision |
| hydroxychloroquine + azithromycin | nitazoxanide | NA | NA | NA | NA | NA | -0.65 | -1.46 | 0.10 | High | NA | -0.65 | -1.46 | 0.10 | Low | Severe imprecision |
| favipiravir | nitazoxanide | NA | NA | NA | NA | NA | 0.04 | -0.80 | 1.01 | Low | NA | 0.04 | -0.80 | 1.01 | Very low | Imprecision |
| remdesivir | nitazoxanide | NA | NA | NA | NA | NA | -0.37 | -1.16 | 0.36 | Low | Intransitivity | -0.37 | -1.16 | 0.36 | Very low | Imprecision |
| convalescent plasma | nitazoxanide | NA | NA | NA | NA | NA | 0.28 | -0.33 | 0.85 | Low | NA | 0.28 | -0.33 | 0.85 | Very low | Imprecision |
| methylprednisolone | nitazoxanide | NA | NA | NA | NA | NA | -0.53 | -1.25 | 0.17 | High | NA | -0.53 | -1.25 | 0.17 | Moderate | Imprecision |
| favipiravir | hydroxychloroquine + azithromycin | NA | NA | NA | NA | NA | 0.70 | -0.29 | 1.84 | Low | NA | 0.70 | -0.29 | 1.84 | Very low | Severe imprecision |
| remdesivir | hydroxychloroquine + azithromycin | NA | NA | NA | NA | NA | 0.28 | -0.57 | 1.14 | Low | Intransitivity | 0.28 | -0.57 | 1.14 | Very low | Imprecision |
| convalescent plasma | hydroxychloroquine + azithromycin | NA | NA | NA | NA | NA | 0.92 | 0.19 | 1.69 | Low | NA | 0.92 | 0.19 | 1.69 | Very low | Severe imprecision |
| methylprednisolone | hydroxychloroquine + azithromycin | NA | NA | NA | NA | NA | 0.12 | -0.71 | 0.97 | High | NA | 0.12 | -0.71 | 0.97 | Moderate | Imprecision |

| | | Direct | | | | | Indirect | | | | | Network | | | | |
|---|---|---|---|---|---|---|---|---|---|---|---|---|---|---|---|---|
| **Treatment 1** | **Treatment 2** | **EST*** | **LCrI*** | **UCrI*** | **Evidence** | **Reason** | **EST** | **LCrI** | **UCrI** | **Evidence** | **Reason** | **EST** | **LCrI** | **UCrI** | **Evidence** | **Reason** |
| remdesivir | favipiravir | NA | NA | NA | NA | NA | -0.36 | -1.53 | 0.31 | Very low | Intransit ivity | -0.36 | -1.53 | 0.31 | Very low | Severe imprecision |
| convalescent plasma | favipiravir | NA | NA | NA | NA | NA | 0.24 | -0.78 | 1.07 | Low | NA | 0.24 | -0.78 | 1.07 | Very low | Imprecision |
| methylprednisolone | favipiravir | NA | NA | NA | NA | NA | -0.57 | -1.65 | 0.35 | Low | NA | -0.57 | -1.65 | 0.35 | Very low | Severe imprecision |
| convalescent plasma | remdesivir | NA | NA | NA | NA | NA | 0.65 | -0.03 | 1.34 | Very low | Intransit ivity | 0.65 | -0.03 | 1.34 | Very low | Imprecision |
| methylprednisolone | remdesivir | NA | NA | NA | NA | NA | -0.16 | -0.94 | 0.64 | Low | Intransit ivity | -0.16 | -0.94 | 0.64 | Very low | Imprecision |
| methylprednisolone | convalescent plasma | NA | NA | NA | NA | NA | -0.81 | -1.47 | -0.14 | Low | NA | -0.81 | -1.47 | -0.14 | Low | NA |

*EST: estimate; LCrI: lower credible interval; UCrI: upper credible interval.

#### Table S13. Sensitivity analysis: fixed-effects model versus fixed-effects model which treated RECOVERY and SOLIDARITY as multiple two-arm trials versus random-effects model (mortality).

| | Fixed-effects | | | Fixed-effects (two-arm) | | | Random-effects | | |
|---|---|---|---|---|---|---|---|---|---|
| **Treatment** | **OR*** | **LCrI*** | **UCrI*** | **OR** | **LCrI** | **UCrI** | **OR** | **LCrI** | **UCrI** |
| Antibacterials for systemic use | 1.00 | 0.48 | 2.06 | 1.00 | 0.46 | 2.18 | 0.98 | 0.48 | 1.97 |
| Azithromycin | 1.00 | 0.89 | 1.12 | 1.00 | 0.89 | 1.12 | 0.98 | 0.71 | 1.28 |
| Antigout preparations | 0.23 | 0.01 | 2.31 | 0.22 | 0.01 | 2.33 | 0.24 | 0.01 | 2.46 |
| Colchicine | 0.23 | 0.01 | 2.07 | 0.22 | 0.01 | 2.06 | 0.24 | 0.01 | 2.23 |
| Antiprotozoals | 1.03 | 0.50 | 2.13 | 1.11 | 0.51 | 2.42 | 0.97 | 0.49 | 1.93 |
| Hydroxychloroquine | 1.03 | 0.93 | 1.15 | 1.11 | 0.99 | 1.25 | 0.98 | 0.77 | 1.19 |
| Antiprotozoals + Antibacterials for systemic use | 0.52 | 0.09 | 2.14 | 0.54 | 0.10 | 2.29 | 0.51 | 0.09 | 2.17 |
| Hydroxychloroquine + Azithromycin | 0.52 | 0.11 | 1.80 | 0.55 | 0.11 | 1.86 | 0.51 | 0.10 | 1.87 |
| Antivirals for systemic use | 0.89 | 0.53 | 1.48 | 0.97 | 0.54 | 1.63 | 0.84 | 0.51 | 1.38 |
| Arbidol | 0.89 | 0.37 | 2.16 | 0.97 | 0.36 | 2.43 | 0.84 | 0.38 | 1.99 |
| Favipiravir | 0.89 | 0.36 | 2.05 | 0.96 | 0.35 | 2.31 | 0.84 | 0.36 | 1.81 |
| Remdesivir | 0.92 | 0.81 | 1.05 | 0.93 | 0.80 | 1.07 | 0.85 | 0.65 | 1.06 |
| Lopinavir/Ritonavir | 0.88 | 0.79 | 0.97 | 1.01 | 0.90 | 1.14 | 0.84 | 0.65 | 1.02 |
| Blood substitutes and perfusion solutions | 0.87 | 0.39 | 1.93 | 0.87 | 0.37 | 2.02 | 0.84 | 0.39 | 1.80 |
| Convalescent plasma | 0.87 | 0.59 | 1.27 | 0.87 | 0.60 | 1.27 | 0.84 | 0.54 | 1.28 |
| Corticosteroids, dermatological preparations | 0.84 | 0.54 | 1.31 | 0.84 | 0.52 | 1.35 | 0.82 | 0.53 | 1.27 |
| Methylprednisolone | 0.91 | 0.68 | 1.29 | 0.91 | 0.68 | 1.30 | 0.89 | 0.62 | 1.34 |
| Dexamethasone | 0.85 | 0.76 | 0.95 | 0.85 | 0.76 | 0.96 | 0.82 | 0.63 | 1.04 |
| Hydrocortisone | 0.77 | 0.53 | 1.04 | 0.77 | 0.52 | 1.05 | 0.76 | 0.50 | 1.08 |
| Immune sera and immunoglobulins | 0.59 | 0.23 | 1.54 | 0.59 | 0.22 | 1.59 | 0.58 | 0.23 | 1.48 |
| Intravenous immunoglobulin | 0.59 | 0.30 | 1.15 | 0.59 | 0.31 | 1.15 | 0.58 | 0.29 | 1.17 |
| Immunostimulants | 0.85 | 0.33 | 1.27 | 0.86 | 0.32 | 1.34 | 0.72 | 0.31 | 1.17 |
| Interferon beta | 0.98 | 0.84 | 1.14 | 1.02 | 0.84 | 1.23 | 0.83 | 0.53 | 1.09 |

| | Fixed-effects | | | Fixed-effects (two-arm) | | | Random-effects | | |
|---|---|---|---|---|---|---|---|---|---|
| **Treatment** | **OR*** | **LCrI*** | **UCrI*** | **OR** | **LCrI** | **UCrI** | **OR** | **LCrI** | **UCrI** |
| Recombinant human GCSF | 0.72 | 0.19 | 1.16 | 0.71 | 0.18 | 1.20 | 0.63 | 0.20 | 1.08 |
| Immunosuppressants | 1.11 | 0.51 | 2.45 | 1.11 | 0.48 | 2.58 | 1.13 | 0.53 | 2.42 |
| Tocilizumab | 1.11 | 0.78 | 1.62 | 1.11 | 0.78 | 1.62 | 1.13 | 0.75 | 1.72 |
| Vitamins | 1.36 | 0.37 | 5.27 | 1.37 | 0.36 | 5.38 | 1.37 | 0.37 | 5.35 |
| Vitamin D3 | 1.36 | 0.45 | 4.37 | 1.37 | 0.45 | 4.34 | 1.37 | 0.44 | 4.60 |
| Immunosuppressants + Antivirals for systemic use | 0.57 | 0.21 | 1.53 | 0.61 | 0.21 | 1.72 | 0.53 | 0.19 | 1.44 |
| Baricitinib + Remdesivir | 0.58 | 0.33 | 1.00 | 0.59 | 0.34 | 1.02 | 0.54 | 0.27 | 1.03 |
| Antithrombotic agents | 0.37 | 0.06 | 1.69 | 0.37 | 0.06 | 1.70 | 0.37 | 0.07 | 1.71 |
| Sulodexide | 0.37 | 0.07 | 1.43 | 0.37 | 0.07 | 1.40 | 0.37 | 0.07 | 1.47 |

*OR: odds ratio; LCrI: lower credible interval; UCrI: upper credible interval.

#### Table S14. Sensitivity analysis: fixed-effects model versus fixed-effects model which treated RECOVERY and SOLIDARITY as multiple two-arm trials versus random-effects model (mechanical ventilation).

| | Fixed-effects | | | Fixed-effects (two-arm) | | | Random-effects | | |
|---|---|---|---|---|---|---|---|---|---|
| **Treatment** | **OR*** | **LCrI*** | **UCrI*** | **OR** | **LCrI** | **UCrI** | **OR** | **LCrI** | **UCrI** |
| Antibacterials for systemic use | 1.01 | 0.18 | 5.57 | 1.02 | 0.20 | 5.13 | 1.05 | 0.26 | 4.26 |
| Azithromycin | 1.01 | 0.85 | 1.20 | 1.02 | 0.85 | 1.21 | 1.05 | 0.68 | 1.61 |
| Antiprotozoals | 0.99 | 0.18 | 5.40 | 1.09 | 0.22 | 5.43 | 0.93 | 0.23 | 3.77 |
| Hydroxychloroquine | 0.99 | 0.86 | 1.15 | 1.08 | 0.92 | 1.28 | 0.94 | 0.68 | 1.25 |
| Antiprotozoals + Antibacterials for systemic use | 1.57 | 0.26 | 9.40 | 1.65 | 0.29 | 9.09 | 1.54 | 0.33 | 7.28 |
| Hydroxychloroquine + Azithromycin | 1.58 | 0.83 | 2.98 | 1.65 | 0.86 | 3.12 | 1.55 | 0.67 | 3.51 |
| Antivirals for systemic use | 1.04 | 0.31 | 3.48 | 1.01 | 0.32 | 3.15 | 0.92 | 0.35 | 2.51 |
| Remdesivir | 1.06 | 0.93 | 1.21 | 0.94 | 0.82 | 1.09 | 0.92 | 0.64 | 1.26 |
| Lopinavir/Ritonavir | 1.02 | 0.88 | 1.17 | 1.08 | 0.92 | 1.27 | 0.93 | 0.65 | 1.29 |
| Blood substitutes and perfusion solutions | 0.96 | 0.17 | 5.39 | 0.96 | 0.19 | 4.95 | 0.96 | 0.23 | 4.00 |
| Convalescent plasma | 0.96 | 0.69 | 1.34 | 0.96 | 0.69 | 1.34 | 0.96 | 0.58 | 1.59 |
| Corticosteroids, dermatological preparations | 0.88 | 0.32 | 2.50 | 0.91 | 0.35 | 2.47 | 0.85 | 0.37 | 2.11 |
| Methylprednisolone | 1.08 | 0.59 | 2.04 | 1.08 | 0.59 | 2.03 | 0.99 | 0.52 | 2.04 |
| Dexamethasone | 0.68 | 0.56 | 0.83 | 0.75 | 0.61 | 0.92 | 0.70 | 0.47 | 1.08 |
| Hydrocortisone | 0.92 | 0.47 | 1.86 | 0.93 | 0.47 | 1.86 | 0.88 | 0.43 | 1.92 |
| Immune sera and immunoglobulins | 0.92 | 0.14 | 5.85 | 0.92 | 0.16 | 5.36 | 0.85 | 0.17 | 4.21 |
| Intravenous immunoglobulin | 0.92 | 0.42 | 2.04 | 0.92 | 0.42 | 2.03 | 0.85 | 0.34 | 2.12 |
| Immunostimulants | 0.46 | 0.12 | 1.49 | 0.45 | 0.13 | 1.35 | 0.47 | 0.14 | 1.17 |
| Interferon beta | 1.00 | 0.85 | 1.18 | 0.92 | 0.76 | 1.12 | 0.78 | 0.45 | 1.15 |
| Recombinant human GCSF | 0.20 | 0.10 | 0.40 | 0.21 | 0.10 | 0.41 | 0.27 | 0.11 | 0.67 |
| Immunosuppressants | 0.67 | 0.12 | 3.88 | 0.67 | 0.13 | 3.53 | 0.67 | 0.15 | 2.91 |
| Tocilizumab | 0.67 | 0.42 | 1.07 | 0.67 | 0.42 | 1.07 | 0.67 | 0.35 | 1.28 |
| Vitamins | 0.42 | 0.06 | 2.80 | 0.42 | 0.07 | 2.57 | 0.42 | 0.08 | 2.23 |

| | Fixed-effects | | | Fixed-effects (two-arm) | | | Random-effects | | |
|---|---|---|---|---|---|---|---|---|---|
| **Treatment** | **OR*** | **LCrI*** | **UCrI*** | **OR** | **LCrI** | **UCrI** | **OR** | **LCrI** | **UCrI** |
| Vitamin D3 | 0.42 | 0.16 | 1.00 | 0.42 | 0.16 | 1.00 | 0.42 | 0.14 | 1.20 |
| Immunosuppressants + Antivirals for systemic use | 0.64 | 0.08 | 5.22 | 0.62 | 0.09 | 4.60 | 0.57 | 0.10 | 3.45 |
| Baricitinib + Remdesivir | 0.65 | 0.43 | 0.99 | 0.58 | 0.38 | 0.88 | 0.57 | 0.25 | 1.22 |
| Antithrombotic agents | 0.44 | 0.04 | 3.83 | 0.43 | 0.05 | 3.59 | 0.44 | 0.05 | 3.23 |
| Sulodexide | 0.44 | 0.09 | 1.77 | 0.44 | 0.09 | 1.77 | 0.44 | 0.08 | 2.00 |

*OR: odds ratio; LCrI: lower credible interval; UCrI: upper credible interval.

#### Table S15. Sensitivity analysis: fixed-effects model versus fixed-effects model which treated RECOVERY and SOLIDARITY as multiple two-arm trials versus random-effects model (discharge).

| | Fixed-effects | | | Fixed-effects (two-arm) | | | Random-effects | | |
|---|---|---|---|---|---|---|---|---|---|
| **Treatment** | **OR*** | **LCrI*** | **UCrI*** | **OR** | **LCrI** | **UCrI** | **OR** | **LCrI** | **UCrI** |
| Antibacterials for systemic use | 1.03 | 0.42 | 2.53 | 1.03 | 0.33 | 3.18 | 0.97 | 0.36 | 2.60 |
| Azithromycin | 1.03 | 0.94 | 1.13 | 1.03 | 0.94 | 1.13 | 0.98 | 0.73 | 1.25 |
| Antiprotozoals | 0.87 | 0.36 | 2.12 | 0.89 | 0.29 | 2.74 | 0.91 | 0.34 | 2.45 |
| Hydroxychloroquine | 0.87 | 0.78 | 0.97 | 0.89 | 0.79 | 0.99 | 0.91 | 0.74 | 1.13 |
| Antiprotozoals + Antibacterials for systemic use | 0.85 | 0.32 | 2.28 | 0.86 | 0.26 | 2.86 | 0.87 | 0.29 | 2.58 |
| Hydroxychloroquine + Azithromycin | 0.85 | 0.53 | 1.39 | 0.86 | 0.53 | 1.41 | 0.87 | 0.50 | 1.53 |
| Antivirals for systemic use | 1.29 | 0.66 | 2.05 | 1.12 | 0.52 | 2.09 | 1.32 | 0.64 | 2.22 |
| Favipiravir | 1.19 | 0.48 | 1.88 | 1.01 | 0.41 | 1.80 | 1.20 | 0.46 | 1.97 |
| Remdesivir | 1.37 | 1.15 | 1.64 | 1.35 | 1.12 | 1.64 | 1.36 | 1.06 | 1.75 |
| Lopinavir/Ritonavir | 1.30 | 1.16 | 1.47 | 1.01 | 0.89 | 1.14 | 1.38 | 1.09 | 1.82 |
| Blood substitutes and perfusion solutions | 1.09 | 0.42 | 2.81 | 1.09 | 0.34 | 3.48 | 1.12 | 0.40 | 3.14 |
| Convalescent plasma | 1.09 | 0.75 | 1.60 | 1.09 | 0.75 | 1.60 | 1.12 | 0.73 | 1.73 |
| Corticosteroids, dermatological preparations | 1.20 | 0.49 | 2.92 | 1.19 | 0.38 | 3.63 | 1.28 | 0.48 | 3.53 |
| Dexamethasone | 1.20 | 1.08 | 1.34 | 1.19 | 1.06 | 1.32 | 1.28 | 0.99 | 1.77 |
| Immunostimulants | 2.15 | 0.79 | 5.87 | 2.16 | 0.63 | 7.24 | 2.17 | 0.73 | 6.39 |
| Interferon beta | 2.16 | 1.26 | 3.74 | 2.16 | 1.26 | 3.74 | 2.17 | 1.23 | 3.88 |
| Immunosuppressants | 1.41 | 0.56 | 3.54 | 1.41 | 0.44 | 4.45 | 1.39 | 0.50 | 3.79 |
| Tocilizumab | 1.40 | 1.05 | 1.89 | 1.40 | 1.04 | 1.89 | 1.39 | 0.98 | 1.95 |
| Immunosuppressants + Antivirals for systemic use | 1.64 | 0.50 | 4.30 | 1.42 | 0.35 | 5.01 | 1.68 | 0.46 | 5.06 |
| Baricitinib + Remdesivir | 1.75 | 1.28 | 2.39 | 1.72 | 1.25 | 2.38 | 1.74 | 1.08 | 2.78 |

*OR: odds ratio; LCrI: lower credible interval; UCrI: upper credible interval.

#### Table S16. Sensitivity analysis: fixed-effects model versus random-effects model (viral clearance).

| | Fixed-effects | | | Random-effects | | |
|---|---|---|---|---|---|---|
| **Treatment** | **OR*** | **LCrI*** | **UCrI*** | **OR** | **LCrI** | **UCrI** |
| Antiprotozoals | 1.44 | 0.49 | 4.56 | 1.17 | 0.36 | 4.52 |
| Hydroxychloroquine | 1.23 | 0.95 | 1.58 | 1.04 | 0.57 | 1.82 |
| Nitazoxanide | 1.72 | 1.13 | 2.80 | 1.34 | 0.53 | 3.97 |
| Antiprotozoals + Antibacterials for systemic use | 0.91 | 0.18 | 4.47 | 0.84 | 0.11 | 6.12 |
| Hydroxychloroquine + Azithromycin | 0.91 | 0.47 | 1.69 | 0.84 | 0.22 | 3.12 |
| Antivirals for systemic use | 1.46 | 0.49 | 5.14 | 1.53 | 0.36 | 6.57 |
| Favipiravir | 1.79 | 0.86 | 4.65 | 1.72 | 0.57 | 5.55 |
| Remdesivir | 1.20 | 0.67 | 2.10 | 1.36 | 0.43 | 4.19 |
| Blood substitutes and perfusion solutions | 2.28 | 0.48 | 10.56 | 2.87 | 0.50 | 16.79 |
| Convalescent plasma | 2.28 | 1.57 | 3.34 | 2.87 | 1.23 | 7.31 |
| Corticosteroids, dermatological preparations | 1.02 | 0.21 | 5.07 | 1.02 | 0.13 | 8.14 |
| Methylprednisolone | 1.02 | 0.59 | 1.76 | 1.02 | 0.25 | 4.23 |

*OR: odds ratio; LCrI: lower credible interval; UCrI: upper credible interval.

#### Supplementary Figures

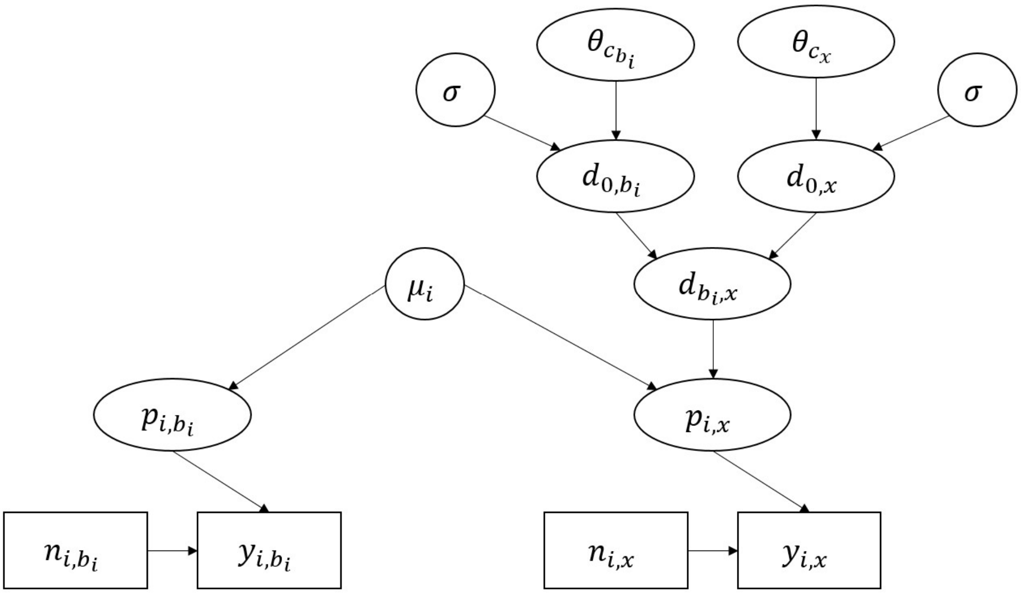

**Figure S1. Bayesian hierarchical framework for trial $i$ in the network meta-analysis, where $(n_{i,x}, y_{i,x})$ are the sample size and number of events in for treatment $x$, $c_y$ is the class of treatment $y$.**

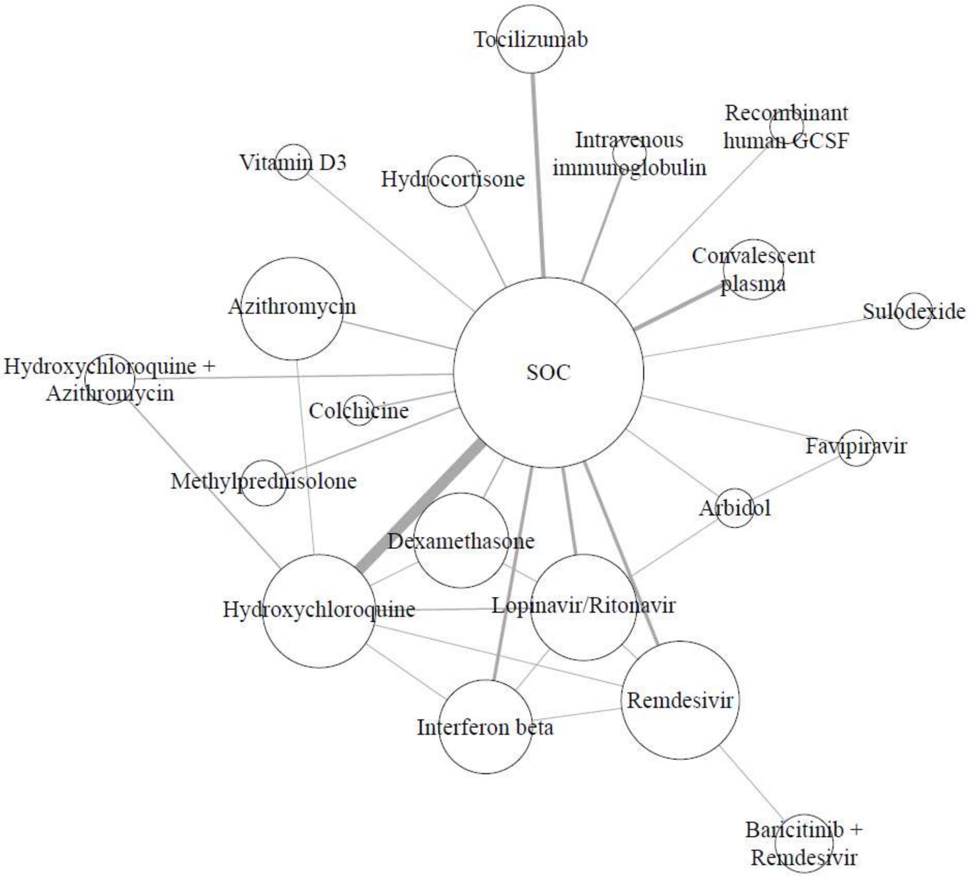

**Figure S2. Network plots for mortality.** The width of the lines is proportional to the number of direct comparisons and the size of the node is proportional to the patients included.

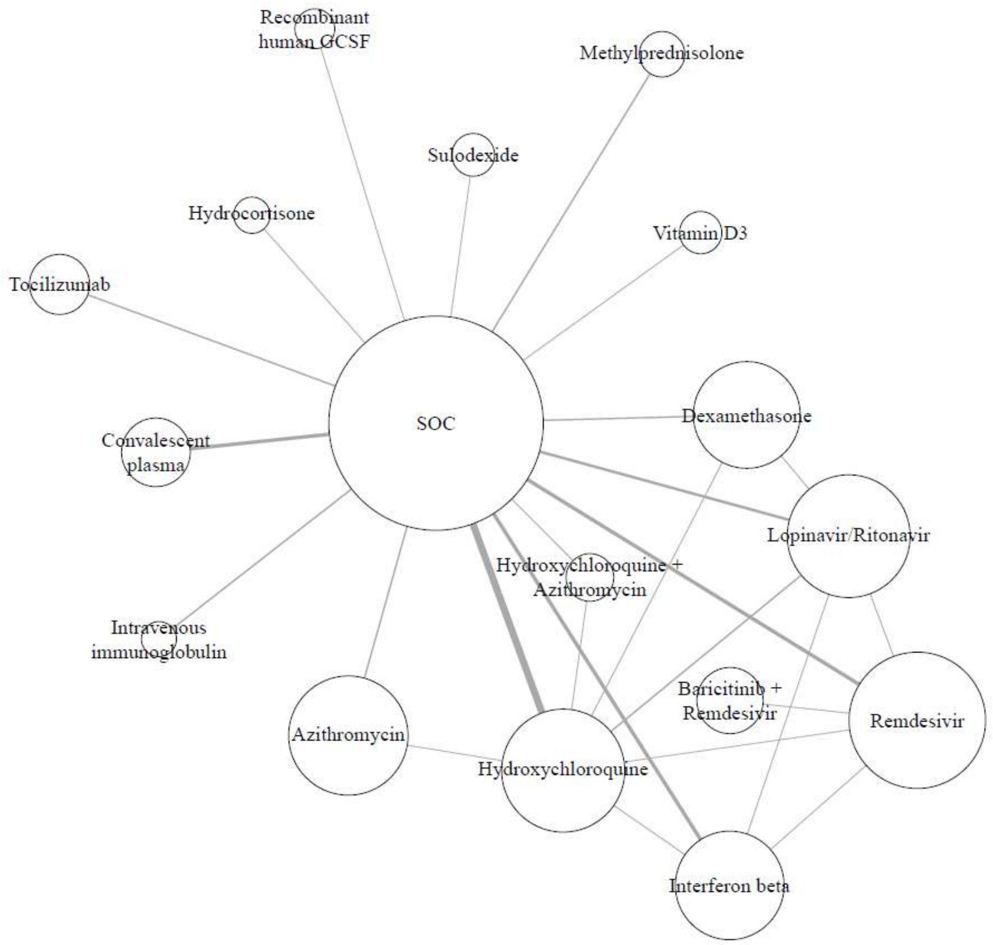

**Figure S3. Network plots for mechanical ventilation.** The width of the lines is proportional to the number of direct comparisons and the size of the node is proportional to the patients included.

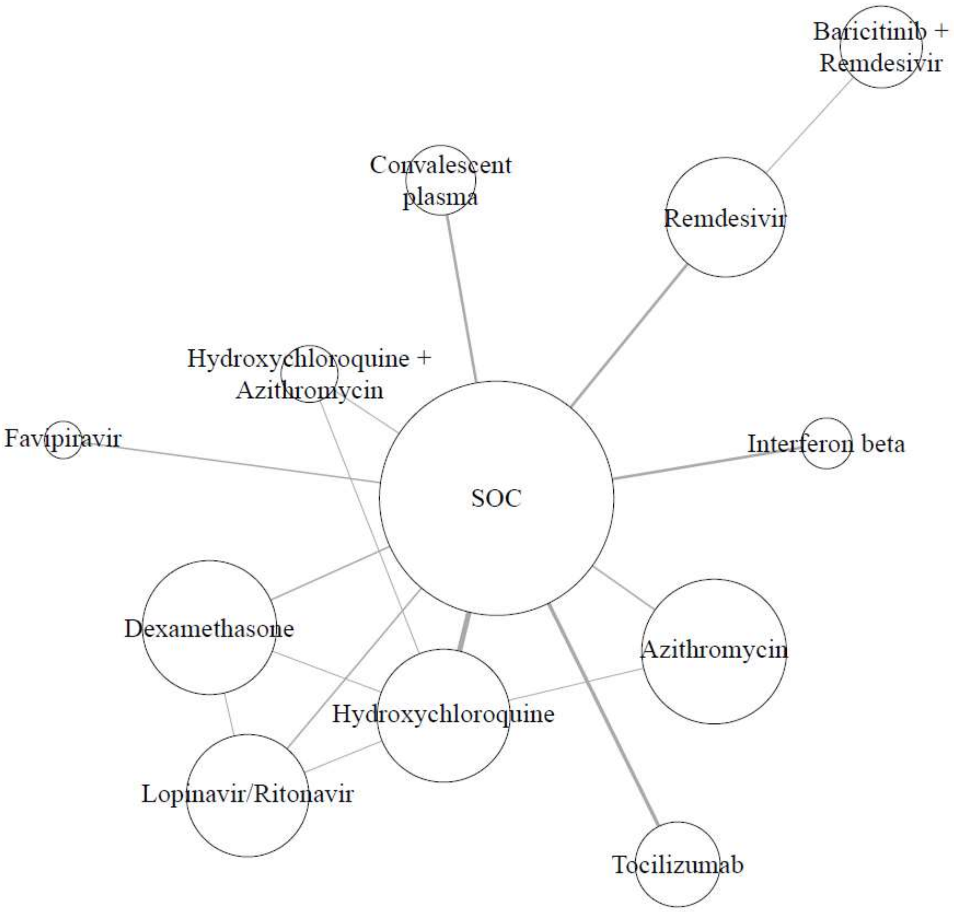

**Figure S4. Network plots for discharge.** The width of the lines is proportional to the number of direct comparisons and the size of the node is proportional to the patients included.

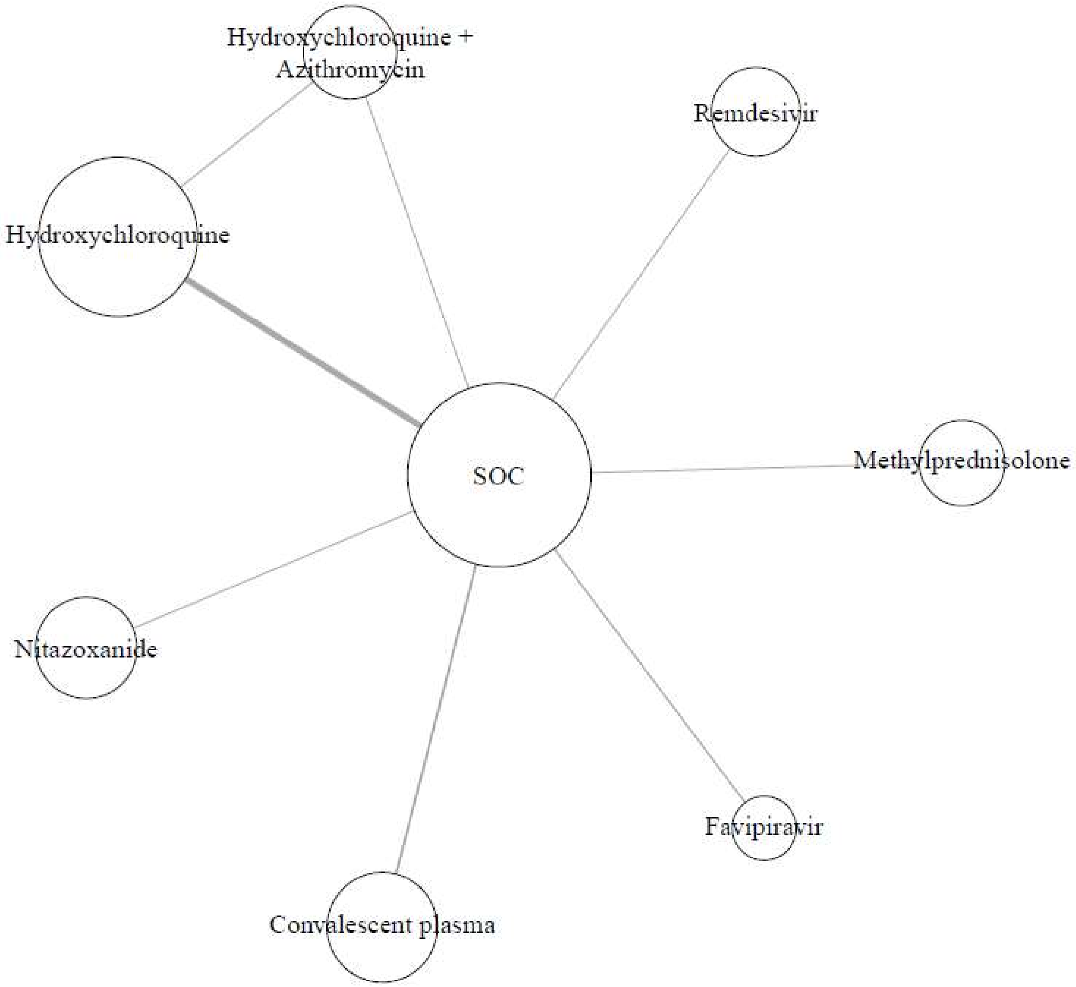

**Figure S5. Network plots for viral clearance.** The width of the lines is proportional to the number of direct comparisons and the size of the node is proportional to the patients included.

| Class<br>Treatment | OR (95% CrI)<br>(Compared to SOC) | Posterior Probability<br>Favouring Treatment |
|---|---|---|
| Antibacterials for systemic use | 0.99 [0.45,2.16] | 0.516 |
| Azithromycin | 0.99 [0.67,1.46] | 0.526 |
| Antiprotozoals | 1.04 [0.51,2.11] | 0.419 |
| Hydroxychloroquine | 1.04 [0.93,1.16] | 0.239 |
| Antiprotozoals + Antibacterials for systemic use | 0.51 [0.09,2.09] | 0.825 |
| Hydroxychloroquine + Azithromycin | 0.51 [0.10,1.76] | 0.848 |
| Antivirals for systemic use | 0.90 [0.55,1.49] | 0.764 |
| Remdesivir | 0.92 [0.81,1.05] | 0.897 |
| Lopinavir/Ritonavir | 0.88 [0.79,0.98] | 0.992 |
| Blood substitutes and perfusion solutions | 0.95 [0.43,2.07] | 0.573 |
| Convalescent plasma | 0.95 [0.64,1.40] | 0.608 |
| Corticosteroids, dermatological preparations | 0.84 [0.53,1.28] | 0.872 |
| Methylprednisolone | 0.89 [0.65,1.26] | 0.771 |
| Dexamethasone | 0.85 [0.76,0.95] | 0.998 |
| Hydrocortisone | 0.77 [0.53,1.04] | 0.956 |
| Immune sera and immunoglobulins | 0.63 [0.24,1.65] | 0.849 |
| Intravenous immunoglobulin | 0.63 [0.32,1.26] | 0.902 |
| Immunostimulants | 0.85 [0.34,1.27] | 0.797 |
| Interferon beta | 0.98 [0.84,1.14] | 0.603 |
| Recombinant human GCSF | 0.73 [0.20,1.17] | 0.885 |
| Immunosuppressants | 1.22 [0.44,3.45] | 0.341 |
| Tocilizumab | 1.22 [0.56,2.73] | 0.312 |
| Immunosuppressants + Antivirals for systemic use | 0.57 [0.21,1.53] | 0.906 |
| Baricitinib + Remdesivir | 0.58 [0.33,1.00] | 0.974 |

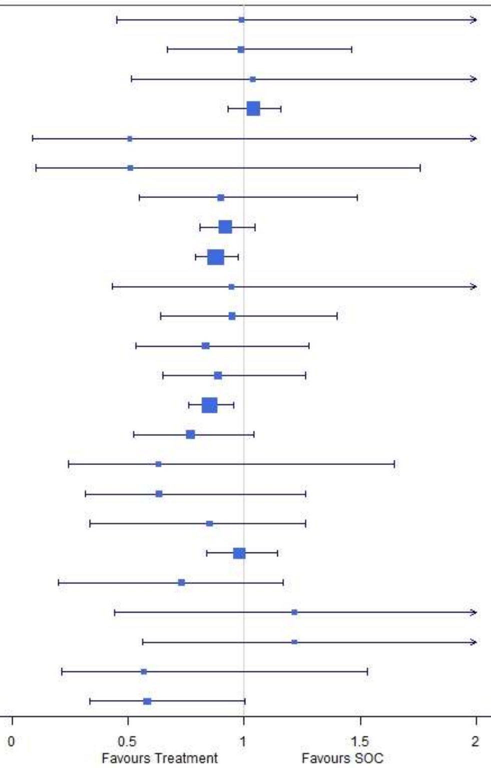

**Figure S6. Subgroup analysis for published studies: mortality under treatments compared with the standard of care (SOC).** OR is the odds ratio and CrI represents credible interval.

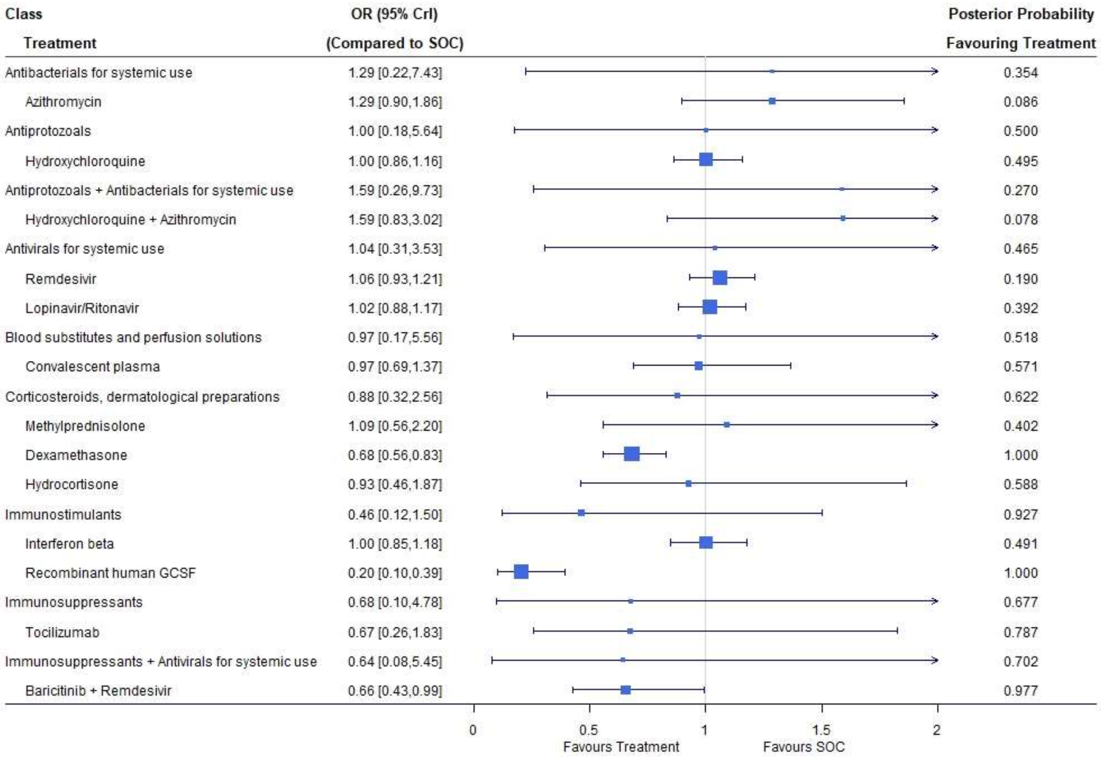

**Figure S7. Subgroup analysis for published studies: mechanical ventilation under treatments compared with the standard of care (SOC).** OR is the odds ratio and CrI represents credible interval.

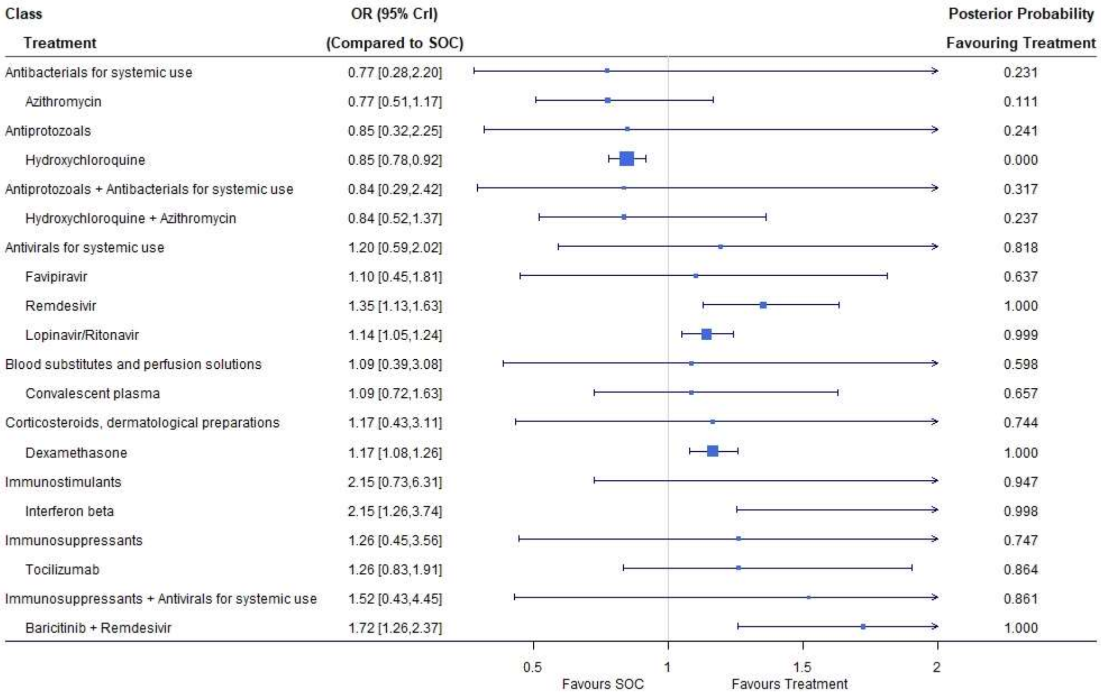

**Figure S8. Subgroup analysis for published studies: discharge under treatments compared with the standard of care (SOC).** OR is the odds ratio and CrI represents credible interval.

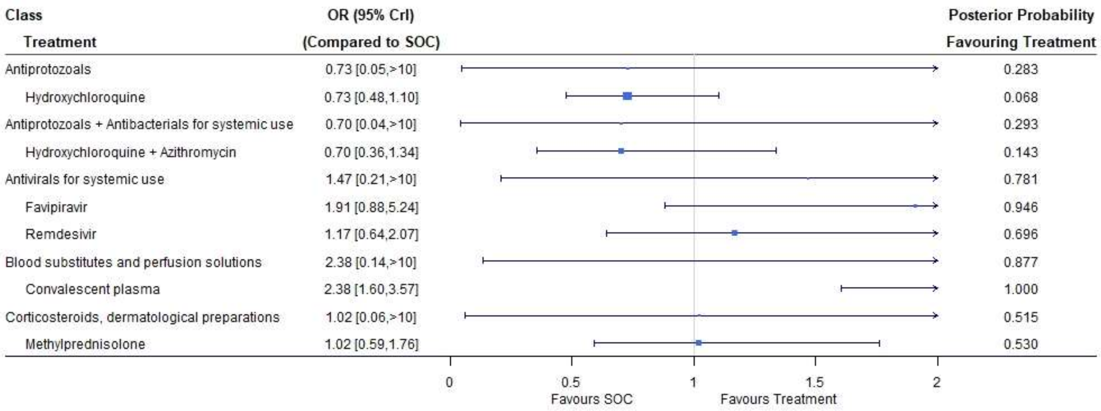

**Figure S9. Subgroup analysis for published studies: viral clearance under treatments compared with the standard of care (SOC).** OR is the odds ratio and CrI represents credible interval.

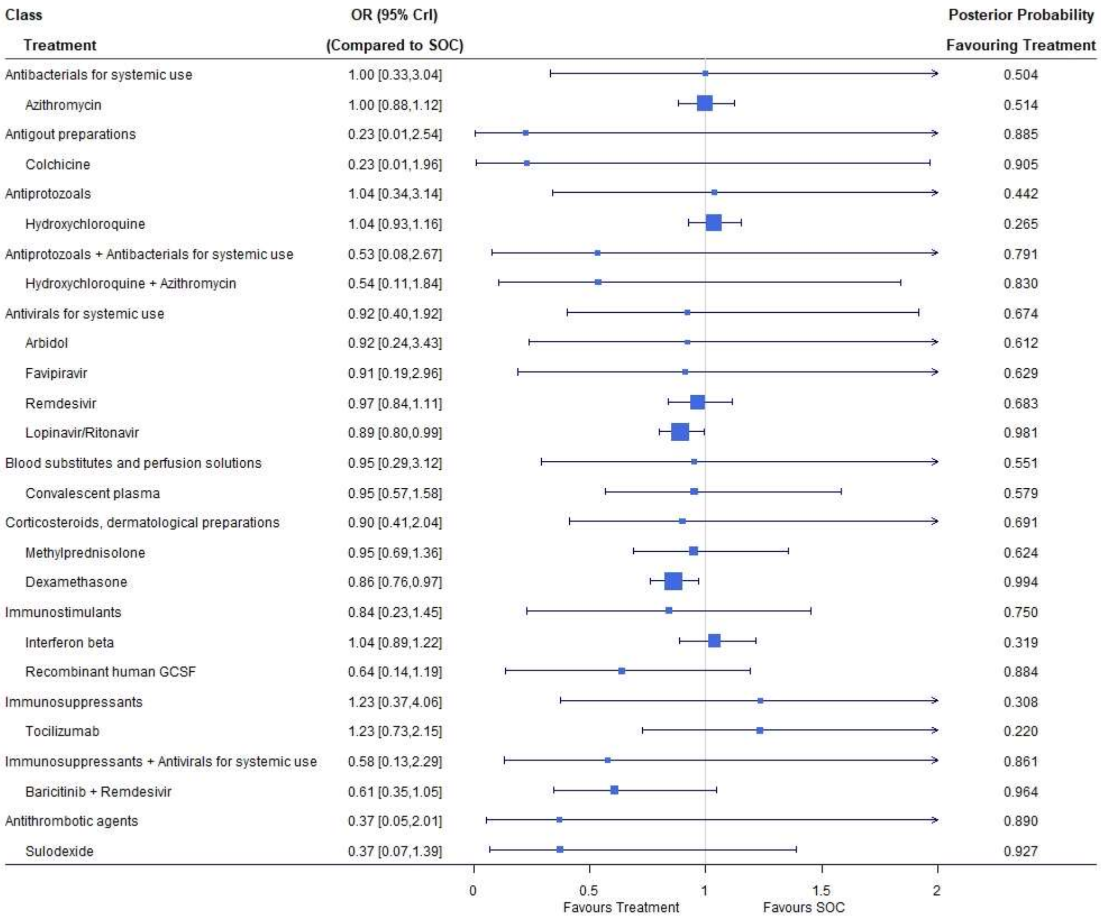

**Figure S10. Subgroup analysis for mild/moderate COVID-19 patients: mortality under treatments compared with the standard of care (SOC).** OR is the odds ratio and CrI represents credible interval.

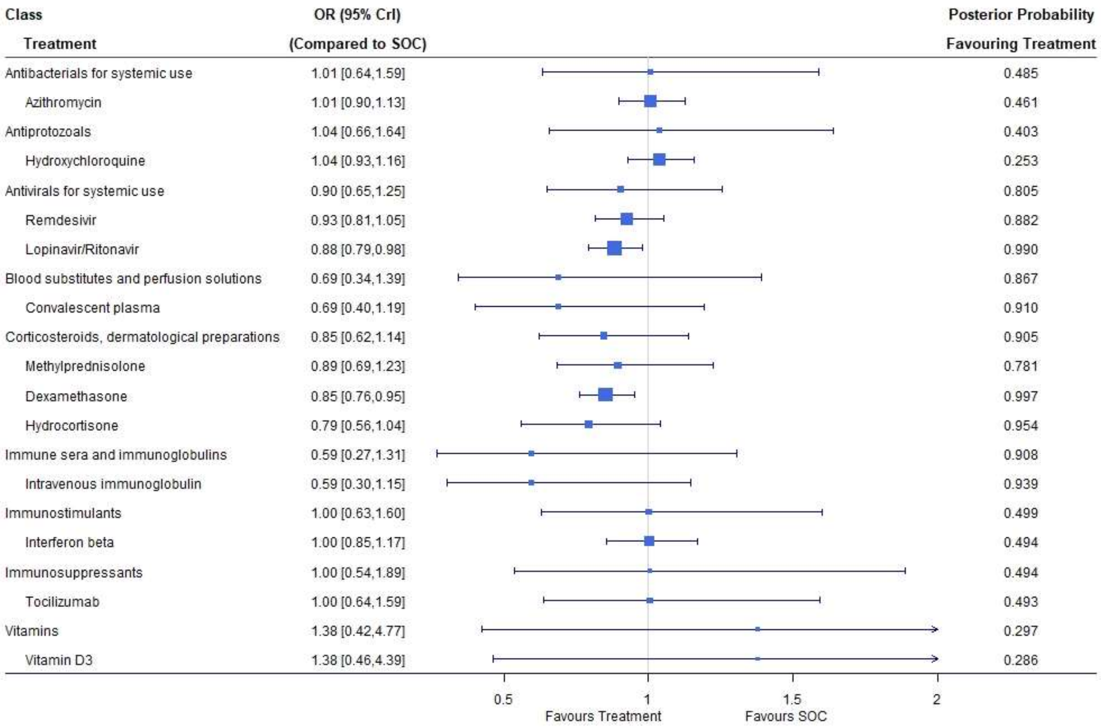

**Figure S11. Subgroup analysis for severe COVID-19 patients: mortality under treatments compared with the standard of care (SOC).** OR is the odds ratio and CrI represents credible interval.

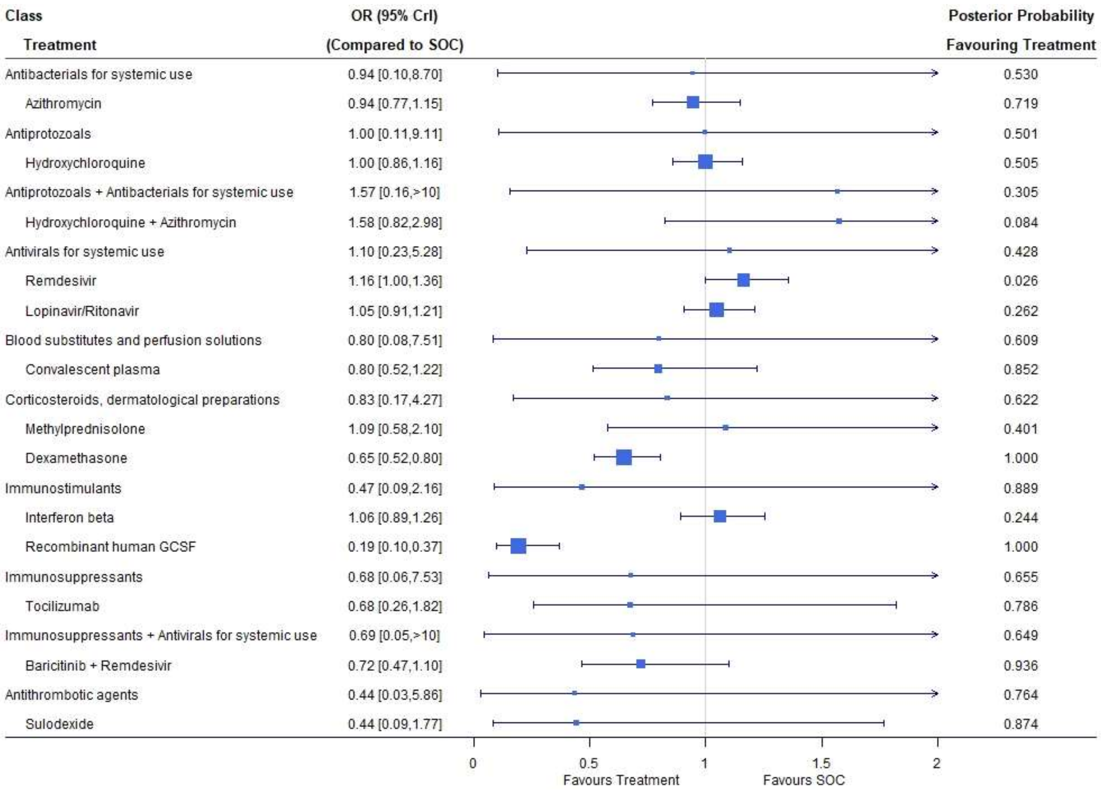

**Figure S12. Subgroup analysis for mild/moderate COVID-19 patients: mechanical ventilation under treatments compared with the standard of care (SOC).** OR is the odds ratio and CrI represents credible interval.

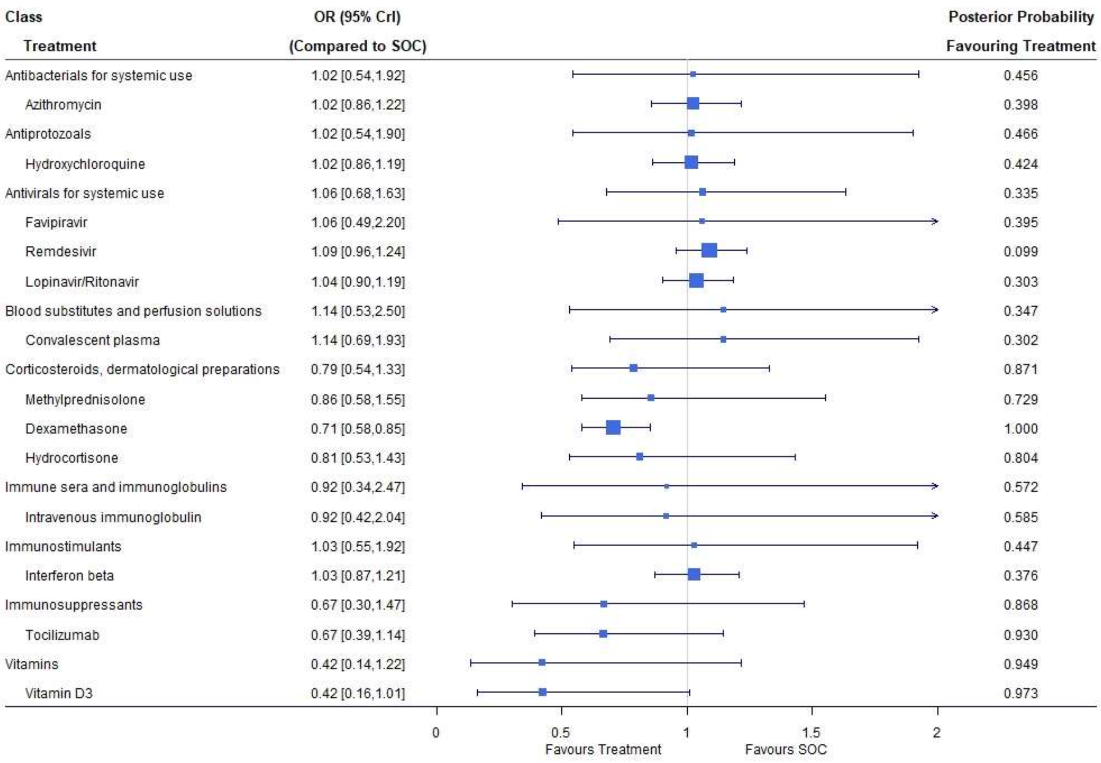

**Figure S13. Subgroup analysis for severe COVID-19 patients: mechanical ventilation under treatments compared with the standard of care (SOC).** OR is the odds ratio and CrI represents credible interval.

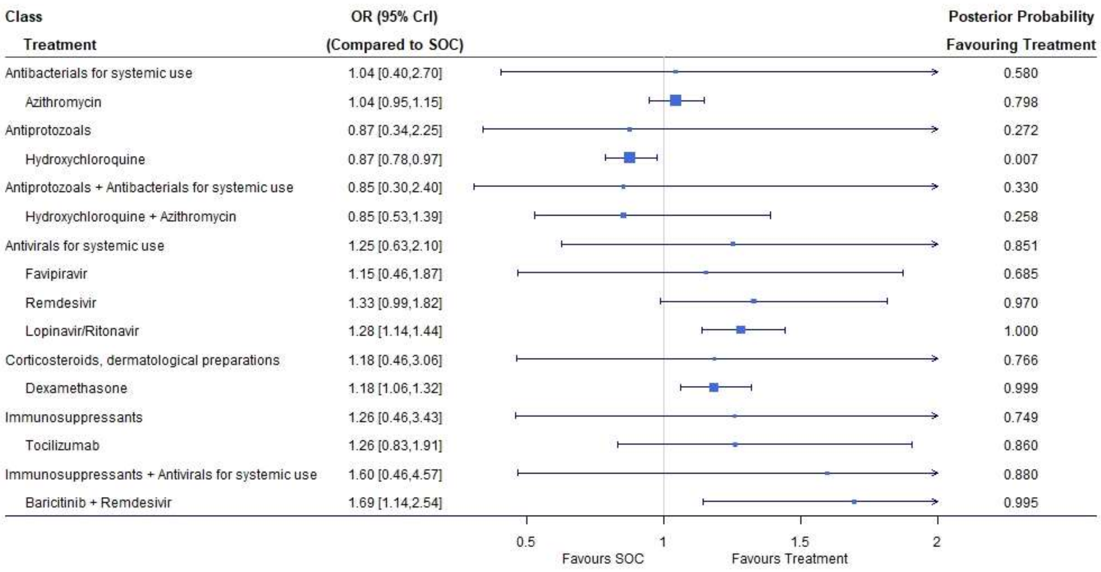

**Figure S14. Subgroup analysis for mild/moderate COVID-19 patients: discharge under treatments compared with the standard of care (SOC).** OR is the odds ratio and CrI represents credible interval.

| Class<br>Treatment | OR (95% CrI)<br>(Compared to SOC) | Posterior Probability<br>Favouring Treatment |
|---|---|---|
| Antibacterials for systemic use | 1.03 [0.27,3.82] | 0.554 |
| Azithromycin | 1.03 [0.94,1.13] | 0.723 |
| Antiprotozoals | 0.85 [0.23,3.15] | 0.246 |
| Hydroxychloroquine | 0.85 [0.75,0.95] | 0.003 |
| Antivirals for systemic use | 1.34 [0.52,3.47] | 0.890 |
| Remdesivir | 1.38 [1.12,1.71] | 0.999 |
| Lopinavir/Ritonavir | 1.30 [1.15,1.46] | 1.000 |
| Blood substitutes and perfusion solutions | 1.09 [0.29,4.26] | 0.609 |
| Convalescent plasma | 1.09 [0.75,1.60] | 0.681 |
| Corticosteroids, dermatological preparations | 1.19 [0.31,4.41] | 0.762 |
| Dexamethasone | 1.19 [1.07,1.33] | 0.999 |
| Immunostimulants | 2.16 [0.53,8.46] | 0.935 |
| Interferon beta | 2.16 [1.26,3.74] | 0.997 |
| Immunosuppressants | 1.53 [0.40,5.91] | 0.873 |
| Tocilizumab | 1.53 [1.07,2.22] | 0.989 |

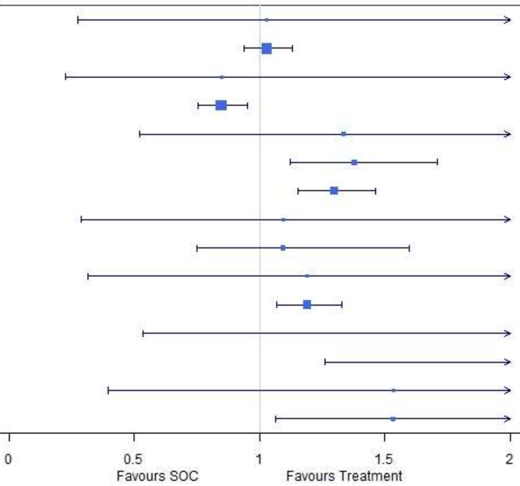

**Figure S15. Subgroup analysis for severe COVID-19 patients: discharge under treatments compared with the standard of care (SOC).** OR is the odds ratio and CrI represents credible interval.

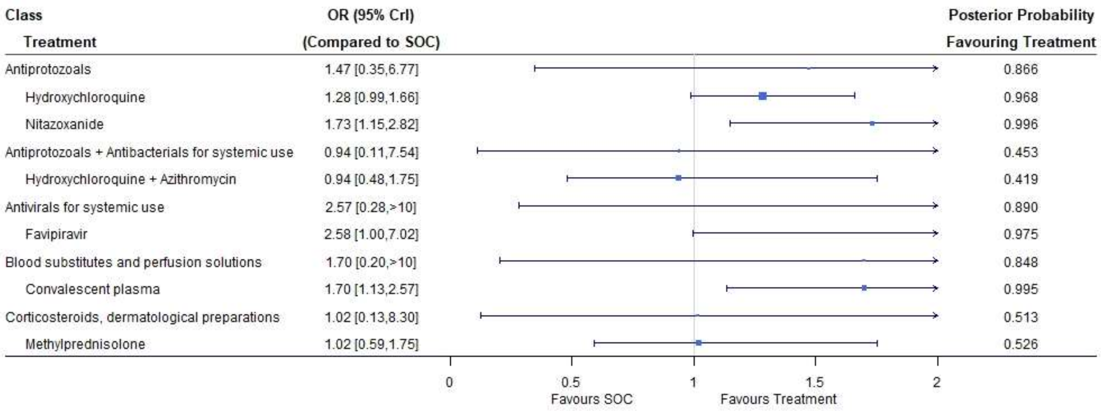

**Figure S16. Subgroup analysis for mild/moderate COVID-19 patients: viral clearance under treatments compared with the standard of care (SOC).** OR is the odds ratio and CrI represents credible interval.

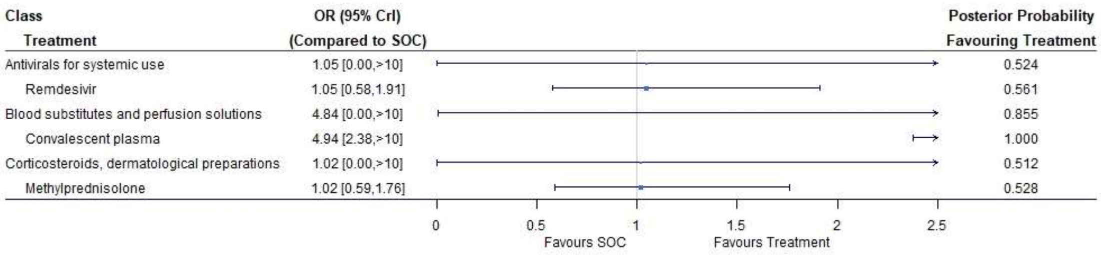

**Figure S17. Subgroup analysis for severe COVID-19 patients: viral clearance under treatments compared with the standard of care (SOC).** OR is the odds ratio and CrI represents credible interval.
